# Type-1 interferon-driven innate and *GZMK*+ CD8 T cell activation precedes subclinical joint inflammation when rheumatoid arthritis is imminent

**DOI:** 10.64898/2026.03.27.26349561

**Authors:** Fareeha Tariq, Paul Martin, Kerem Abacar, Weiyu Ye, Shawn Sun, Sophie MacKay, Dylan Muldoon, Sana Sharrack, Madhvi Menon, Hussein Al-Mossawi, Maya H Buch, Paul Emery, Darren Newton, Benjamin P Fairfax, Kulveer Mankia

**Affiliations:** Leeds Institute of Rheumatic and Musculoskeletal Medicine, University of Leeds, Leeds, UK; NIHR Leeds Biomedical Research Centre, Chapel Allerton Hospital, Leeds Teaching Hospitals NHS Trust, Leeds, UK; Centre for Genetics and Genomics, Centre for Musculoskeletal Research, The University of Manchester, Manchester, UK; NIHR Manchester Biomedical Research Centre, Manchester University NHS foundation Trust, Manchester Academic Health Science Centre, Manchester, UK; Department of Dermatology, Oxford University Hospitals NHS Foundation Trust, Oxford, UK; Department of Oncology, University of Oxford, Oxford, UK; Division of Immunology, University of Manchester, Manchester, UK; Centre for Musculoskeletal Research, University of Manchester, Manchester, UK; Leeds Institute of Medical Research, University of Leeds, Leeds, UK; Department of Rheumatology, King’s College Hospital, London, UK

## Abstract

Rheumatoid arthritis is a prototypical autoimmune disease, characterised by prolonged systemic autoimmunity prior to organ-specific tissue inflammation. To achieve the contemporary goal of autoimmune disease prevention, a nuanced understanding of the transition from systemic autoimmunity to tissue-specific inflammation is critical. Here, we sought to identify immune signatures associated with the transition to subclinical joint inflammation detected by multi-joint ultrasound in anti-citrullinated protein antibodies (ACPA+)-positive individuals who imminently progress to RA. To achieve this, we performed single-cell transcriptomic and proteomic profiling on prospectively collected blood samples from high-risk ACPA+ imminent progressors, who were further stratified by the presence or absence of ultrasound (US)-detectable subclinical synovitis and compared them with ACPA+ non-progressors. We found type-1 interferon (IFN-I) activation in circulating CD14+ classical monocyte and *GZMK*+ CD8+ T cells preceding subclinical joint inflammation in ultrasound-negative (USneg) future progressors. In contrast, US-positive (USpos) future progressors exhibited a phenotypic shift in CD14+ classical monocytes towards IL1ß+ expression and clonal expansion of *GZMB*+ cytotoxic CD8+ T cells at the onset of subclinical synovitis. Plasma proteomics also revealed a shift from Toll-like receptor–associated innate pathways in USneg future progressors toward effector and tissue-remodeling signatures in USpos future progressors. These findings suggest IFN-I-driven immune priming in specific immune subsets precedes the onset of subclinical joint inflammation, whereas tissue-directed inflammatory and cytotoxic programmes emerge at the onset of joint inflammation when clinical RA is imminent.

## Introduction

Rheumatoid arthritis (RA) is a common, prototypical autoimmune disease, which usually develops after a preclinical phase characterised by systemic autoimmunity and the presence of circulating autoantibodies such as anti-citrullinated protein antibodies (ACPA) (*1*). Individuals who are seropositive for ACPA but do not have clinically detectable arthritis are considered ‘at-risk’, representing a potential window of opportunity for arthritis prevention, prior to established tissue-specific inflammation and damage. The ‘at risk’ phase of RA encompasses a broad range of clinical and pathobiological phenotypes (*2*). Asymptomatic at-risk individuals often have a low short-term risk of developing arthritis and may be reluctant to receive immune modulating treatment. In contrast, ACPA-positive (ACPA+) individuals with arthralgia have more advanced pathobiology (*3, 4*), are at higher risk, and often seek healthcare for their symptoms. Consequently, these individuals are more motivated to receive early interventions, and therapeutic trials are more feasible to power and successfully execute (*5*). Two recent clinical trials have demonstrated that short-term treatment with the T cell co-stimulation modulator, abatacept can alleviate symptoms and produce modest preventive benefit when used in such high-risk individuals (*6, 7*). To advance progress towards preventative therapies in those at high risk of developing RA, a comprehensive and nuanced understanding of the immune mechanisms that underpin the transition from autoimmunity to tissue-centric inflammation (i.e. arthritis) is required. Recent integrated multi-omics analyses have identified a broad systemic immune activation in ACPA-positive at-risk individuals without clinical arthritis (*8–13*). Furthermore, longitudinal studies suggest the expansion of inflammatory innate and adaptive immune cells occurs when at risk individuals progress to clinically detectable arthritis. However, insights into the specific mechanisms that accompany the late transition from systemic autoimmunity to joint-centric inflammation – the so called ‘second hit’ of RA pathogenesis – remains elusive. This late transition occurs when the clinical onset of RA is imminent and represents a stage in the disease continuum that is particularly challenging to capture and investigate.

We aimed to address this important question by performing blood single-cell transcriptomic and proteomic profiling in ACPA+ individuals at high risk of RA (high ACPA titres, HLA-DR shared epitope+, arthralgia), who were prospectively identified and sampled. Individuals were further stratified at time of sampling using whole-body multi-joint ultrasound assessment to identify the presence or absence of early subclinical joint inflammation. In individuals who imminently developed clinical RA, we identified a transcriptional signature of type-1 interferon (IFN-I)-driven innate and adaptive immune activation preceding any detectable synovial joint involvement, and a shift towards inflammatory, cytotoxic phenotypes coinciding with the transition to joint-specific inflammation.

### 1. Peripheral immune cell phenotypes define functional heterogeneity in at-risk individuals

To comprehensively characterize peripheral blood immune cell heterogeneity in ACPA+ individuals with arthralgia at high risk of developing RA, including subgroups who went on to develop RA imminently, we performed single cell RNA sequencing (scRNAseq) on peripheral blood immune cells (PBMCs) from 40 ACPA+ at risk individuals (Figure 1a) (*1*). Among these, 22 individuals were selected because they had gone on to progress to RA within 6 months of identification and PBMC sampling (hereafter termed ‘imminent future progressors’). Importantly, imminent future progressors were further stratified according to whether there was evidence of subclinical synovial tissue inflammation, based on ultrasound assessment of 38 joints and 10 tendons. As such, imminent future progressors where further divided into ‘ultrasound (US)-negative’ (USneg, n = 11) and ‘US-positive’ (USpos, n = 11) groups. US positivity was determined by the presence of subclinical joint and/or tendon inflammation, detected by a power Doppler (PD) score ≥1 and a Grayscale (GS) score ≥1 in at least one joint and/or tendon. The remaining 18 at-risk individuals, although high risk, did not develop RA during the 24-month follow-up period and were US-negative at baseline (non-progressors, NP). Baseline demographic and clinical characteristics are summarized in Supplementary Table S1.

**Figure 1.**
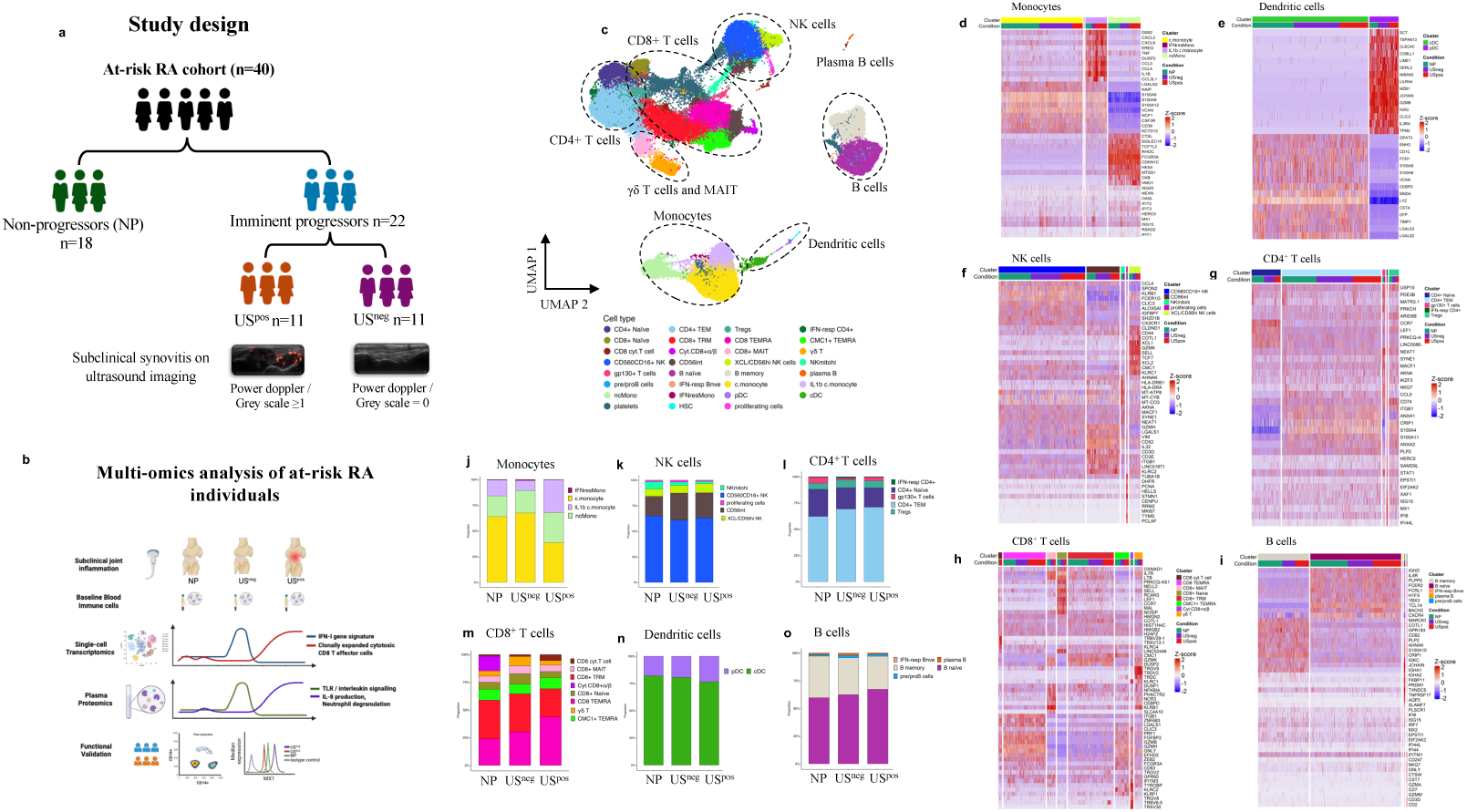
Single cell transcriptomic profiling of individuals at high risk of RA. a) ACPA+ individuals with arthralgia at high risk of developing RA (n = 40) were stratified into non-progressors at 24 months (NP; n = 18) and imminent progressors who went on to develop arthritis within 6 months (n = 22). Imminent future progressors were further divided into those with subclinical joint and/or tendon inflammation denoted by power Doppler/greyscale ≥ 1 on multi-joint ultrasound assessment (USpos n = 11) and those without detectable joint and/or tendon inflammation (USneg n = 11). b) Overview of the integrated workflow including clinical imaging assessments, baseline blood collection, single-cell transcriptomics, plasma proteomics and functional validation. c) UMAP projection of peripheral immune cells coloured by major cell type, highlighting monocytes, NK cells, CD4+ T cells, CD8+ and γδ T cells/MAIT, B cells, dendritic cells and plasma B cells. Cells were first clustered using a shared nearest-neighbour graph built on the top 30 principal components, followed by Leiden community detection (resolution = 1.0); UMAP was computed on the same 30-PC space to visualise the resulting clusters. d-i) Heatmaps showing the top 10 cluster-defining genes for each indicated immune population. Marker genes were identified with FindAllMarkers using the MAST test (test.use = MAST) on the 30-PC expression space, with thresholds of log₂ fold change ≥ 1, minimum detection fraction (min.pct) ≥ 0.40, and return.thresh = 0.05; genes with adjusted P value (Benjamini-Hochberg) < 0.01 were retained, and the top 10 markers per cluster were visualised. j-o) Proportions of cells per subset within each major lineage (monocytes (j), NK cells (k), CD4+ T cells (l), CD8+ T cells (m), dendritic cells (n) and B cells (o) in the three clinical groups (NP, USpos and USneg), shown as stacked bars.

The experimental workflow (Fig. 1b) included single-cell RNA sequencing (scRNA-seq) of cryopreserved PBMCs collected at baseline, Olink proteomic profiling (Explore HT) of matched plasma samples, and flow cytometric analysis to validate transcriptomic findings. For scRNA-seq, approximately 5,000 cells per sample were profiled using 5′ gene expression and variable, diversity, joining (V(D)J) libraries, yielding a mean depth of ∼20,000 reads per cell for transcriptome data and ∼3,000 reads per cell for paired (V(D)J) sequencing. Using the Leiden algorithm for unsupervised clustering, we identified 31 transcriptionally distinct cell clusters. Cell cluster identities were assigned based on canonical markers defined by the COVID-19 Multi-omics Blood Atlas Consortium (*14*) and further validated using the FindAllMarkers function in Seurat v5 (*15*) (Supplementary Fig. S1a-e, S2). We identified 31 transcriptionally distinct clusters encompassing all major immune lineages including, monocytes, natural killer (NK) cells, dendritic cells (DCs), T cells and B cells (Fig. 1c).

We first examined the broad transcriptomic features defining each cell subset to establish a comprehensive atlas of immune cell states in ACPA+ individuals at high risk of developing RA. Subset-defining transcripts identified using the MAST test (log2 fold change ≥ 1 and adjusted p < 0.01 BH) highlighted discrete transcriptional programs across immune lineages (Fig. 1d-i). Among monocytes, four transcriptionally distinct subsets were identified: CD14+/CD16− classical, CD14−CD16+ non-classical, CD14+IL1β+ inflammatory monocytes, and a small subset of CD14+ cells with high expression of type-1 interferon-stimulated genes (ISGs), which clustered separately from other monocytes. We termed this interferon-high cluster as IFNresMono. CD14+ classical monocytes were characterised by upregulation of *S100A8*, *S100A9*, and *S100A12*, together with *CD36* and *VCAN*, defining an inflammatory population primed for endothelial adhesion and tissue migration (*16*). The IL1β+ classical monocytes showed elevated expression of chemokines (*CXCL2, CXCL8, CCL3, CCL4*) and cytokines (*TNF, IL1β)* along with G0S2 and *DUSP* genes, indicating a strong pro-inflammatory profile and synovial priming (*17*). CD16+ non-classical monocytes exhibited upregulation of *SIGLEC10, TCF7L2, CDKNIC*, and *RHOC*, and a concomitant downregulation of inflammatory S100A family genes, indicating a divergent functional program between these subsets. Upregulation of *SIGLEC10*, a negative regulator of DAMP-driven inflammation, together with *CDKN1C* (a cell-cycle inhibitor) and *RHOC* (a Rho GTPase involved in cytoskeletal mobility), suggests that CD16+ monocytes are primed for intravascular patrolling and endothelial surveillance, reflecting a shift away from inflammation toward a regulatory, homeostatic state (*18, 19*).

The IFNresMono formed a distinct minor cluster (Fig. S3) with markedly increased expression of at least nine ISGs (*IFIT1, IFIT2, IFIT3, ISG15, MX1, RSAD2, ISG20, OASL*), which were not expressed at comparable levels in other monocyte subsets (Fig. 1d). This pattern is consistent with an acute IFN-I-driven transcriptional burst, sufficient to override subset-defining transcriptional programs. A similar, lower-intensity ISG signature was, however, detectable across the remaining monocyte subsets, suggesting a broad IFN exposure, which did not affect their core cellular identities.

Dendritic cells were subdivided into plasmacytoid DCs (pDCs) and conventional DCs (cDCs). pDCs had high expression of *CLEC4C, LILRA4, IRF7, MZB1, GZMB*, and *NDRG2*. The *IRF7* and *MZB1* co-expression indicates a transcriptionally poised state for IFN-I secretion, while *GZMB* and *NDRG2* suggest activation-associated stress adaptation characteristic of pDCs in autoimmune inflammation (*20*). cDCs expressed *CD1C, S100A8, S100A9*, and *LYZ* indicative of an antigen-presenting phenotype (Fig. 1e).

The NK cell compartment formed four distinct clusters: CD56dimCD16hi, CD56int, NK mitohi, and *XCL1*+CD56hi (Fig. 1e). The CD56dimCD16hi displayed increased expression of tissue-homing genes including *KLRB1, CX3CR1, SPON2*, and *CCL4* and reduced expression of *GZMK, HLA-DRB1* and *HLA-DRA* genes (*21*). CD56int population exhibited an activation-primed NK phenotype, with features of enhanced cytotoxic readiness and inflammatory responsiveness (*KLRC2, ITGB1, IL32*), alongside reduced expression of adaptive NK cell-associated markers (*KLRB1, FCER1G*). The NK mitohi is a metabolically active CD56dim-like effector subset with an enrichment of mitochondrial/OXPHOS transcripts (*MT-CYB, MT-CO3, MT-ATP*) and elevated *HLA-DRA/B1* with reduced *GZMK, CCL4*, and *KLRB1* expression. Finally, the *XCL1*+ CD56hi NK subset was characterised by elevated *XCL1/2*, *GZMK*, *TCF7*, *COTL1* and reduced expression of *CX3CR1* consistent with a tissue-associated, less differentiated population involved in local immune orchestration.

The T cell compartment, encompassing CD4+ and CD8+ T cells, exhibited pronounced transcriptional heterogeneity with five distinct CD4+ and eight CD8+ subsets (Fig. 1g-h). Among CD4+ T cells, the naïve population was marked by reduced expression of inflammatory S100A family genes, however, the T effector memory (CD4+ TEM) cells had elevated expression of *S100A8/9*, *STAT1* and sparse upregulation of IFN-I-stimulated genes. We additionally found two smaller CD4+ T cell subsets, one gp130 (IL6ST)-expressing, and the other only enriched interferon-responsive genes (IFN-resp CD4+, Fig. 1g, S3). The gp130+ subset showed increased expression of *NEAT1*, *CD74*, and *STAT1* and downregulation of S100A family genes. Gp130 is a signal-transducing subunit for IL-6 family cytokines acting through JAK/STAT pathway (*22*). Although IL-6 signalling is known to promote Th17 differentiation, canonical Th17 markers such as *RORC* and *IL23R* were not detected, indicating this subset represents cytokine-responsive CD4+ T cells rather than committed Th17 cells. Additionally, a Treg subset, identified by expression of *FOXP3/IL2RA*, displayed *CD74, ITGB1*, and *S100A* upregulation with *CCL5* downregulation, consistent with an activated, tissue-engaged regulatory program.

The CD8+ T cell compartment comprised naïve (CCR7+), activated effector-memory (TRM) (*GZMK+DUSP2+GZMB*–*IL7R+*), *CMC1+* TEMRA, CD8+ TEMRA with cytotoxic effector phenotype (*CCR7*–*GZMB*+*GZMK*–*PRF1+*), MAIT (*KLRB1+*), gamma delta (γδ) T cells (TRD/TRG) and two smaller subsets, CD8 cyt. T cells and cyt.CD8α/β, retained cytotoxic features but were distinguishable by differences in T-cell receptor α and β chain usage (Figure 1h). The CD8 TEMRA subset was characterised by increased expression of *ITGB1*, *ZNF683*, and *IFITM3*, consistent with terminal cytotoxic population displaying tissue-resident-associated and interferon-responsive features. *ITGB1* has been reported as a marker of large cytotoxic CD8+ T cell clones, suggesting a potential clonal expansion within this subset (*23, 24*). Both MAIT and γδ were enriched for *GZMK*, *KLRB1*, and *DUSP2*. However, MAIT cells uniquely showed upregulation of *SLC4A10* and *DUSP1*, indicating an early activation and mucosal identity.

The B cell compartment consisted of transcriptionally distinct naïve, memory, and plasmablast-like subsets, with a few small pre/pro B and interferon-responsive populations (Fig 1i). Naïve B cells expressed *FCRL1*, *CXCR4*, and *IL4R*, consistent with resting and cytokine-responsive phenotypes, while memory B cells were enriched for *S100A10*, *CD82*, and *IFITM1*, reflecting antigen experience and enhanced activation potential. IFITM1 expression across both naïve and memory clusters indicates broad type-1 interferon exposure within the circulating B cell pool. The plasma cell subset expressed high levels of *IGHG*, *IGHA*, and *PRDM1*, defining a terminally differentiated, antibody-secreting population.

### 2. CD14+ monocytes and CD8+ TEMRA proportional shifts across subclinical synovitis

We next examined whether the transcriptionally distinct immune cell subsets identified above differed in proportional abundance between at-risk groups (NP, USneg, USpos). Statistically significant differences were observed in selected monocyte and CD8+ T cell subsets, whereas additional populations exhibited consistent, albeit non-significant, directional trends across the at-risk continuum. Among monocytes, the CD14+ classical subset was more abundant in USneg future progressors compared with USpos future progressors (Fig. 1j, S4d) suggesting potential migration from the periphery into inflamed joints and/or tendons at the onset of subclinical inflammation before clinical arthritis develops. The IFNres-Mono subset was significantly enriched in USneg future progressors compared to USpos future progressors (p = 0.012, Wilcoxon rank-sum test; Fig. S4d), suggesting marked IFN-1-driven monocyte activation prior to the onset of synovial tissue inflammation when progression to arthritis is imminent. In contrast, IL1β+ monocytes were enriched in USpos future progressors, indicating an expansion of this inflammatory myeloid subset coinciding with early subclinical joint inflammation in the pre-RA phase (Fig. Ij, S4d). In the NK cell compartment, the overall distribution of subsets was comparable across groups, apart from a higher proportion of mitochondrial-high NK cells (NKmitohi) in NP and increased frequencies of CD56int NK cells in both USneg and USpos future progressors, the latter consistent with the expansion of adaptive-like NK populations in individuals closer to arthritis onset (Fig. 1k, S4c).

Within the adaptive lymphoid compartment, the CD4+ T cell population displayed progressive compositional shifts, with naïve and gp130+ subsets enriched in NP, effector memory (TEM) cells most abundant in USpos future progressors, and Tregs relatively increased in USneg future progressors (Fig 1l). The CD8+ T cell compartment demonstrated the most striking differences. The CD8+ TEMRA and *CMC1*+ TEMRA cells were significantly elevated in USpos future progressors (p = 0.03 and p = 0.04, respectively, Fig. S4b), suggesting the expansion of these cells is associated with the onset of subclinical synovial tissue inflammation even when individuals are still in the pre-RA phase. In contrast, naïve CD8+ T cells were relatively enriched in USneg future progressors and CD8+ TEM proportions were reduced in USpos future progressors compared to both USneg and NP. The γδ T cell subset was also expanded in USneg future progressors, suggesting preferential activation or retention of innate-like T cells before the development of synovial inflammation (Fig. 1m).

The B cell compartment showed broadly similar distributions across groups, with naïve and memory B cells comprising the majority. Memory B cells were modestly higher in NP compared with USneg future progressors (p = 0.049, Fig. S4f), whereas plasma, pre/pro B, and IFN-responsive B-cell subset (IFNres B) remained infrequent across all groups. Together, these data suggest shifts in immune cell profiles as high-risk individuals with arthralgia approach the onset of tissue inflammation, when clinically apparent arthritis is imminent (Fig. 1o).

### 3. IFN-I activation in innate subsets precedes subclinical synovitis in ACPA+ imminent progressors

To assess whether alterations in immune cell composition among the three at-risk groups were associated with differential transcriptional activity, we performed DESeq2-based differential gene expression analysis across the 31 immune cell subsets. Comparisons were made between (i) USneg future progressors vs. NP (ii) USpos future progressors vs. NP, and (iii) USneg future progressors vs. USpos future progressors. Across innate immune lineages (monocyte and NK cell subsets), IFN-I-related transcriptional activity was preferentially enriched across USneg future progressors (Fig. 2a), indicating that IFN-I pathway activation emerges before US-detectable synovial inflammation becomes evident in imminent progressors. In CD14+ classical monocytes, USneg future progressors exhibited a robust upregulation of IFN-I-stimulated genes (ISGs), including *MX1*, *IFI6*, *ISG15*, and *IFIT3* (Fig. 2b). A similar ISG pattern was observed in IL1ß+ inflammatory monocytes when comparing USneg with USpos future progressors, despite a lower overall frequency of this subset in USneg group, suggesting that IFN-I activation in monocytes precede overt inflammatory expansion of immune cells. In CD16+ non-classical monocytes, CD56dimCD16hi NK, and mitochondrial-high NK (NK mitohi) subsets, IFN-I activation was primarily restricted to *MX1*, which was selectively upregulated in USneg future progressors compared to NP (Fig. 2c). Protein-level validation further confirmed increased *MX1* and *ISG15* expression in CD14+ classical monocytes and CD56dimCD16hi NK cells from USneg future progressors compared to NP and USpos future progressors, supporting concordant transcriptional and translational IFN-1 pathway activation in these subsets (Fig 2c, S5-7).

**Figure 2.**
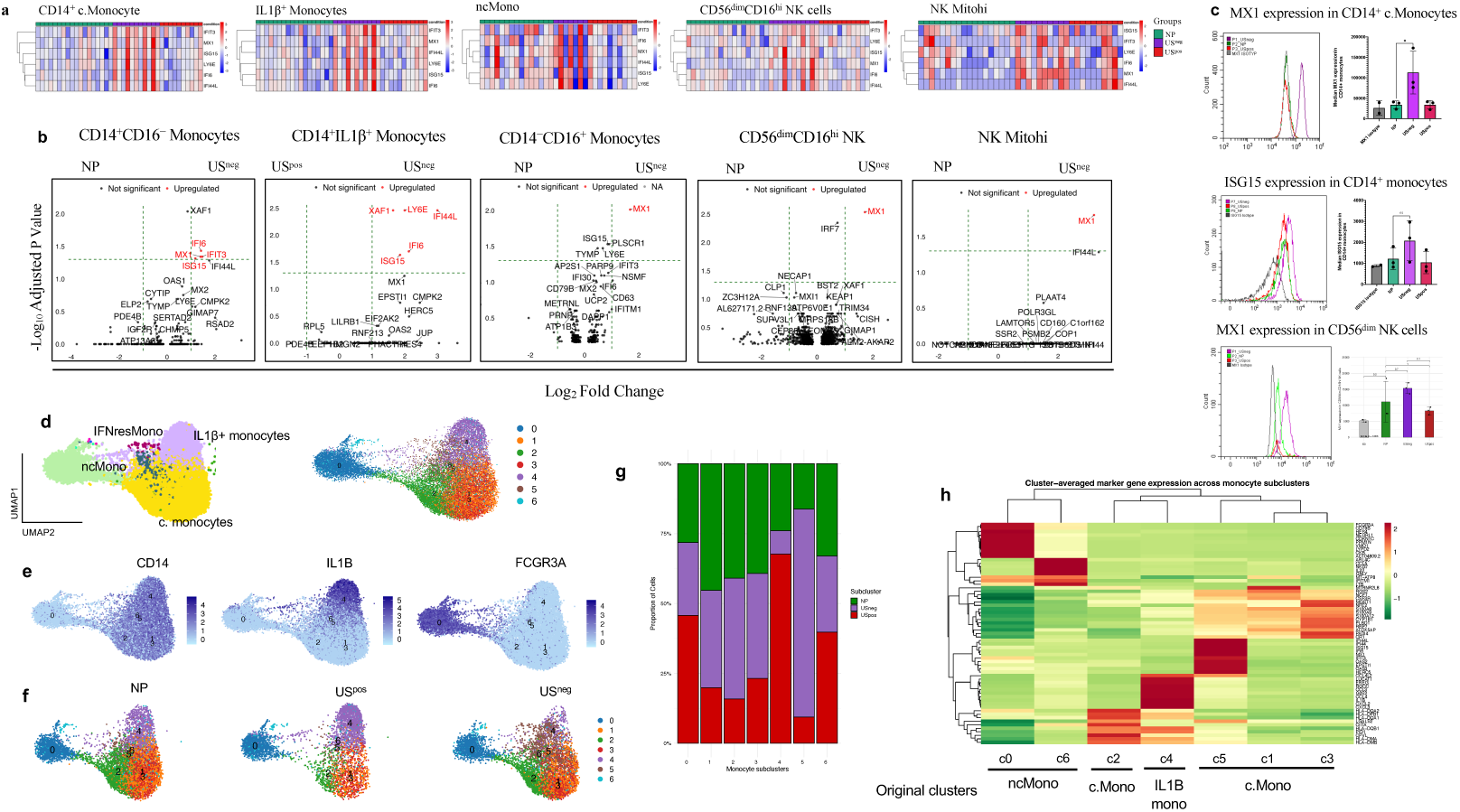
Upregulation of type-1 interferon (IFN-I) genes in innate immune subsets in ACPA+ high-risk individuals with imminent RA but no detectable synovial inflammation (USneg group). a) Heatmap showing per-individual expression of type I interferon (IFN-I)-stimulated genes across innate immune subsets, demonstrating enrichment of ISG expression in USneg future progressors compared with NP and USpos future progressors. b) Volcano plots displaying significantly upregulated ISGs (red; DESeq2, padj < 0.05, log₂FC > 1) in USneg progressors relative to NP and USpos groups within CD14+CD16-classical monocytes, IL1β+ monocytes, CD16+CD14– non-classical monocytes, CD56dimCD16hi NK, and NK mitohi subsets. c) Flow cytometry validation showing higher MX1 and ISG15 protein expression in CD14+CD16-monocytes and CD56dimCD16hi NK cells in USneg future progressors compared with both NP and USpos future progressors. d) Secondary Leiden clustering (resolution = 0.4) of all monocyte subsets identified seven transcriptionally distinct subclusters. e) UMAP feature plots of CD14, IL1β, and FCGR3A expression delineating major classical, IL1β+, and non-classical monocyte compartments in subclusters. f-g) UMAP projection and proportional distribution of subclusters across NP, USneg, and USpos groups showing expansion of ISG-enriched clusters in USneg future progressors and IL1β+ cluster in USpos future progressors. h) Heatmap of the top 10 marker genes per subcluster (identified using FindAllMarkers, log₂FC ≥ 1, min.pct ≥ 0.25, adjusted p < 0.05). The dendrogram illustrates transcriptional relationships among related subclusters.

We next examined the biological context of these transcriptional changes by performing pathway-level enrichment analysis of DESeq2 results. Across all innate cell subsets, there was a convergent antiviral and IFN-I-associated transcriptional program. Notably, RIG-I-like receptor signalling was enriched exclusively in CD14+ classical and IL1β+ inflammatory monocytes, the two subsets showing the highest number of IFN-I upregulated genes (Supplementary Fig S8a-b). Within these populations, genes associated with cytosolic RNA sensing and downstream IFN-I signalling were markedly upregulated, including IFI6 (Fig. 2a, b), consistent with intrinsic activation of the RIG-ITBK1-IRF3/7 axis (*25*). In contrast, *MX1*-expressing subsets such as CD16+ non-classical monocytes and NK cell subsets showed enrichment primarily for viral response pathways (e.g., coronavirus disease, human papillomavirus, and measles) without a detectable RIG-I module (Fig. S7c-e).

To further investigate the transcriptional heterogeneity within monocyte subsets, we performed sub-clustering analysis on all four monocyte subsets using Leiden clustering (resolution = 0.4). This revealed seven transcriptionally distinct subclusters (Fig. 2d, S9). The *FCGR3A*/CD16+ non-classical monocytes (cluster c0) and IL1ß+ monocytes (cluster c4) did not separate into further subclusters (Fig. 2d-e). Proportions of both these clusters were dominant in USpos future progressors (Fig. 2g). The CD14+CD16– classical monocyte formed 4 subclusters (clusters c1, c3, c2, c5) based on differences in transcriptomic profiles (Fig 2d-e). All four of these subclusters showed varied proportions across the three at-risk groups, with subcluster c5 predominant in USneg future progressors compared to both USpos and NP, and subclusters c1, c2, and c3 showed similar proportions in USneg and NP but reduced in USpos (Fig 2g). Cluster c6 was the smallest subcluster that grouped together with FCGR3A-expressing cluster c0.

We then examined transcriptional heterogeneity among these seven subclusters by identifying differentially expressed genes using FindAllMarkers() function in Seurat (padj <0.05, Log2FC.thresh = 0.25) and visualised top 10 marker genes per subcluster in heatmap (Fig. 2h). Subclusters c0 and c4 retained the transcriptional profiles of parent clusters (ncMono and IL1ß+ monocyte) with upregulation of *CDKNIC*, *VMO1*, and *IL1ß* alongside chemokines (*CXCL2/3* and *CCL3/4*), respectively. The c.Mono subclusters (subclusters c1, c2, c3, c5) showed distinct transcriptional heterogeneity. Clusters c1, and c3 were defined by upregulation of inflammatory S100A family genes (*S100A8, S100A9, S100A12*), and other genes including *PAD14*, *CR1*, and *PLBD1*, however, these genes were more enriched in c3 (Fig. 2h). Cluster c5 showed a strong enrichment of ISGs including *IFI44L*, *IFI44*, *ISG15*, *IFI6*, *MX1*, *OAS2*, and *OAS3*. In initial clustering analysis, the ISG signature was distributed among CD14+ classical monocyte and a small IFN-responsive monocyte subset (section 1, Fig. 1d), however, subclustering resolved them into a distinct ISG-high subset. Cluster c2 segregated from other subclusters due to upregulation of HLA genes (*HLA-DMA/DMB/DQA2/DQA1/DPB1/DQB1*), which encode components of the MHC-II antigen-presentation pathway (Fig. 2h) (*26*). The HLA family genes were also slightly upregulated in IL1ß+ cluster c4. This suggests that a subset of classical monocytes acquires an antigen-presenting phenotype capable of engaging and activating antigen-specific cells.

### 4. Expansion of IFN-I activated *GZMK*+ CD8+ T cells and cytotoxic *GZMB*^+^ CD8^+^ T cell clones occur at different stages in imminent RA

Given the differential abundance of CD8+ T cells between patient groups (Fig. 1m), we undertook a more focused analysis of transcriptional states within the CD8+ T cell compartment. CD8+ T cell subsets identified in section 1 were re-clustered after removing TCR (V(D)J) genes from the expression matrix to prevent TCR-driven clustering (see Methods). This analysis resolved CD8+ cytotoxic, CD8+ TEMRA, CD8+ TRM, and MAIT/γδ T cells into well-defined transcriptional states on the lineage-aware UMAP, forming four stable islands that were preserved across donors (Fig. 3a). The heatmap of cluster-defining transcripts revealed four major cell states, a *GZMB*- and *PRF1*-expressing TEMRA-like cytotoxic state, a naïve/central memory-like population, a *GZMK*+ effector-like, and a MAIT or γδ TRM-like state (Fig. 3c). These four states were defined by cytotoxic genes (*PRF1, GZMB, GNLY*, and *NKG7*), memory-associated genes (*TCF7, CCR7, IL7R*), chemokine-associated effectors (*GZMK, CCL3, CCL4*) and residency markers (*KLRB1, RORC, ZNF683*, and *ITGAE*), respectively.

**Figure 3.**
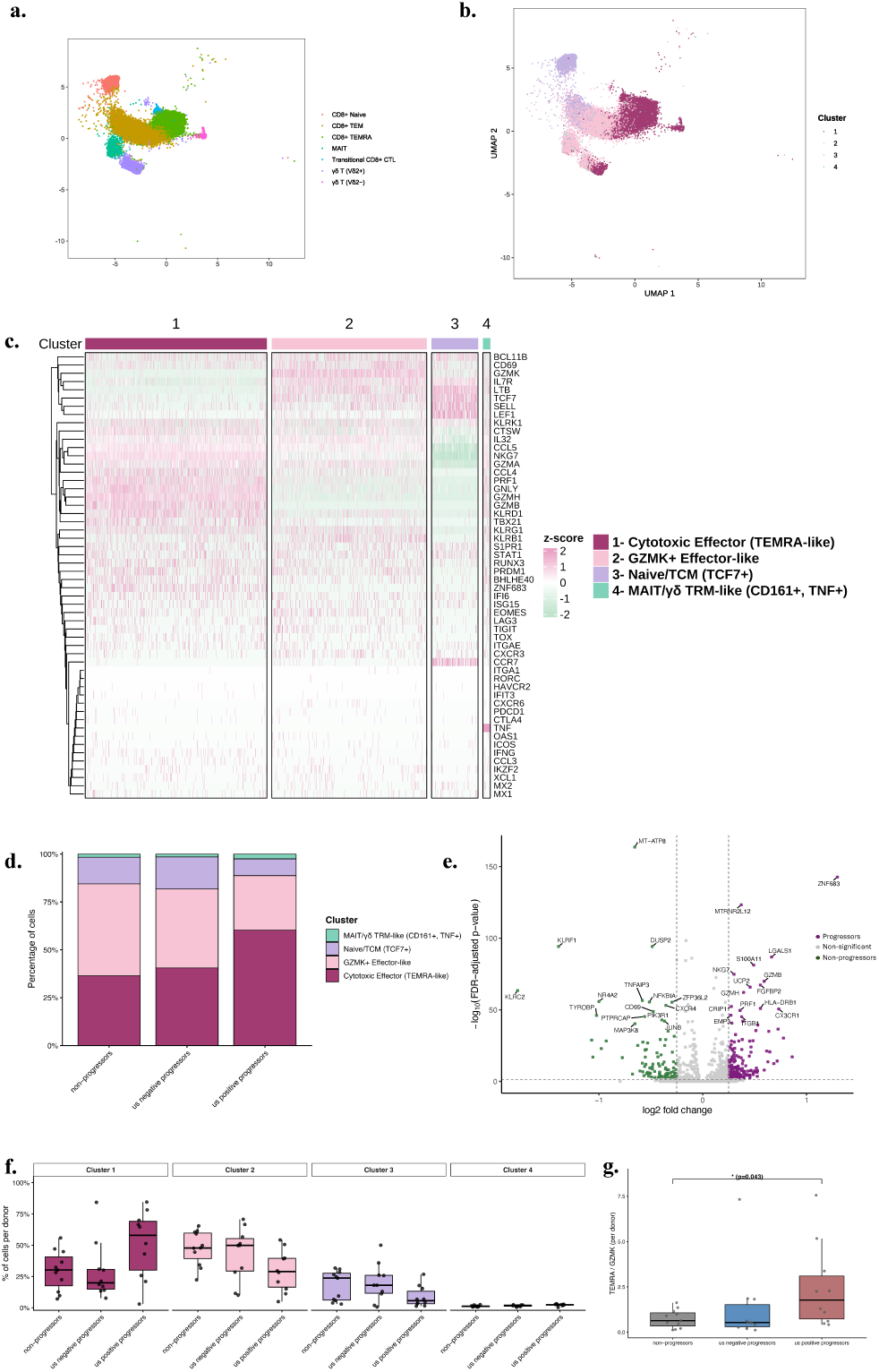
a-b) UMAP projections of CD8+ T cells demonstrated distinct transcriptional states across NP, USneg future progressors and USpos future progressors, with clusters corresponding to naïve or central memory-like, *GZMK+* effector-like, cytotoxic TEMRA-like and MAIT or γδ TRM-like programmes. c) A heatmap of these clusters confirmed clear transcriptional separation, with each state defined by coherent and functionally divergent gene-expression profiles. d) The relative abundance of each cluster differed across clinical groups, with progressors (purple) showing higher frequencies of cytotoxic and TRM-like cells than NP (green). e) Differential expression analysis showed that progressors exhibited cytotoxicity- and interferon-associated signatures, whereas NP expressed higher levels of immediate-early and regulatory transcripts. f) Donor-level quantification demonstrated consistent shifts in cluster composition across individuals within each clinical group. g) Comparisons of key subsets showed that the TEMRA-like/*GZMK+* effector cluster ratio were higher in USpos future progressors than USneg future progressors and NP.

USpos future progressors showed higher proportions of TEMRA-like and MAIT/γδ TRM-like populations suggesting a marked expansion of these populations at the onset of subclinical synovial inflammation. In contrast, naïve and GZMK+ effector-like cells were reduced, potentially reflecting migration of these cells into synovial tissue (Fig. 3d). Differential expression analysis demonstrated higher *PRF1, GZMB, CX3CR1, FGFBP2*, and *ZNF683* in all future progressors (USpos and USneg), while NP retained higher levels of *DUSP2, JUNB, NR4A2*, and *TNFAIP3* (Fig 3e). These profiles reflect an enrichment of cytotoxic and residency imprints alongside a contraction of naïve and *GZMK*+ effector-like states in individuals who are imminently progressing to RA (Fig 3f). In line with this, the ratio of TEMRA-like to *GZMK*+ clusters were significantly higher in USpos future progressors compared with NP (p = 0.043) (Fig. 3g).

We next compared expanded CD8+ clonotypes and their transcriptional profiles between the three at-risk groups. Expanded clonotypes were strongly concentrated within TEMRA-like regions of the UMAP and were most frequent in USpos future progressors (Fig. 4a). Donor-level clonality increased in a stepwise fashion from NP to USneg and USpos future progressors. Shannon entropy was reduced in progressors, consistent with diminished repertoire diversity and increased clonal dominance (Fig. 4b) Clone-class distribution analyses further confirmed progressive expansion of large clonotypes from NP to USneg and USpos groups (Fig. S10 a-c).

**Figure 4.**
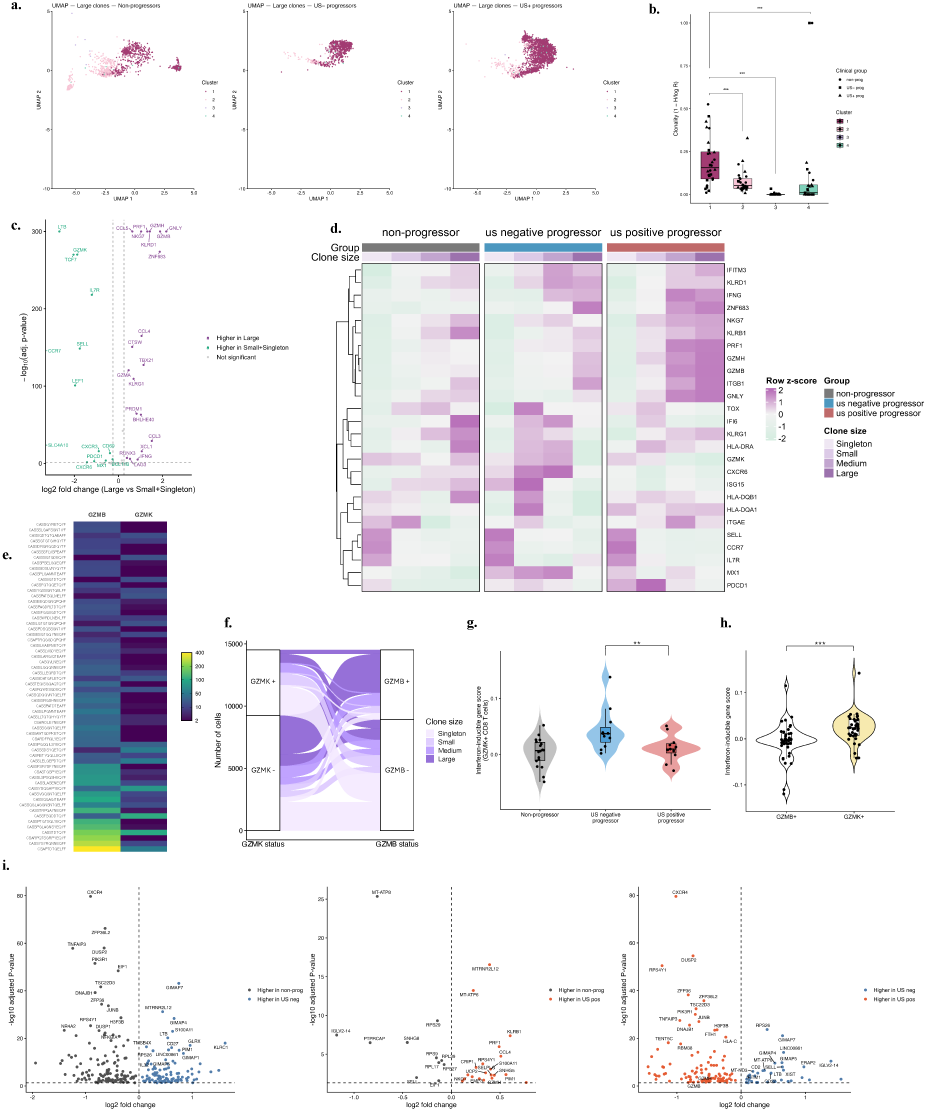
CD8 clonality in ACPA+ at-risk groups. a) UMAP projections displaying large-clone CD8+ T cells overlaid onto the transcriptional clusters defined in Figure 4 showed that USpos future progressors had a marked increase in large clones localising to the cytotoxic effector (TEMRA-like) cluster. b) Donor-level clonality analysis across clusters shows significantly higher clonality in the cytotoxic effector (TEMRA-like) cluster (*** p<0.001, ** p<0.01, * p<0.05). c) Volcano analysis of large versus small or singleton clonotypes demonstrates cytotoxic gene enrichment in expanded clone. d) A heatmap of transcriptional profiles stratified by clone size and clinical group highlights gene-expression differences between expanded and smaller clonotypes. e) Shared clonotype analysis between *GZMB+* and *GZMK+* cytotoxic states show minimal overlap and asymmetric expansion of the few shared CDR3 identities. f) The distribution of *GZMB+*, *GZMK+* and double-positive cells across clone-size classes indicates that each cytotoxic phenotype arises from distinct clonotypic backgrounds. g) *GZMK+ CD8+* T cell type 1 interferon inducible signature is higher in USneg future progressors compared with USpos future progressors (p<0.01) and NP (non-significant trend) h) *GZMK+ CD8+* T cells exhibited a stronger interferon inducible signature marked by elevated *MX1, ISG15, IFI6, OASL, and IFIT3* compared with *GZMB+ CD8+* T cells. i) Gene expression in *GZMK+ CD8+* T cells across three at-risk groups, CXCR4 expression was higher in both NP and USpos future progressors than in USneg future progressors. In contrast, increased expression of cytotoxic genes in USpos future progressors, whereas *CD27* expression was higher in USneg future progressors.

Across all patients, large clonotypes exhibited higher expression of *PRF1*, *GZMB*, *GNLY*, *NKG7*, *CX3CR1*, and *ZNF683*, whereas small or singleton clonotypes showed higher *CCR7*, *SELL, IL7R, TCF7, LEF1, DUSP2, JUNB*, and *NR4A2* (Fig. 4c). Donor-equal heatmap analysis showed that USneg future progressors exhibited higher *GZMK, CXCR6*, and *ISG15* expression within small and medium clonotypes, whereas USpos future progressors displayed increased cytotoxic signatures predominantly within large clones (Fig. 4d). Across all patients, *GZMB*+ and *GZMK*+ cytotoxic programmes were supported by largely distinct clonotypes, with shared CDR3 identities representing only a very small minority of the repertoire (Fig. 4e). *GZMK* expression was observed across smaller clonotypes, whereas *GZMB* expression was more prominent in larger clonotypes, with double-positive cells representing only a small fraction of the repertoire (Fig. 4f).

Given the enrichment of *GZMK*+ and IFN-1–inducible genes in USneg future progressors on heatmap analysis (Fig. 4d), we compared IFN-1 gene signatures in *GZMK+* CD8+ T cells between USneg future progressors and the other patient groups. *GZMK*+ CD8+ T cells from USneg future progressors showed higher expression scores for interferon-inducible genes including *MX1*, *ISG15, IFI6, OASL*, and *IFIT3* compared with NP (Fig. 4g). IFN-1 signature scores were higher in *GZMK*+ than in *GZMB+* CD8+T cells (Fig. 4h). When *GZMK*+ CD8+ T cells were compared across clinical groups, *CXCR4* expression was higher in both NP and USpos future progressors than in USneg future progressors. In contrast, cytotoxic genes were increased in USpos future progressors, whereas *CD27* expression was higher in USneg future progressors (Fig. 4i).

### 5. Proteomics reveals transition from TLR signalling to neutrophil degranulation in future progressors

To determine whether the systemic proteomic profile reflected the stage-specific immune signatures described above, we performed plasma proteomic profiling (Olink Explorer HT) on all study participants. Differential abundance analysis was performed using the limma package on PCA-normalised NPX values which identified a total of 549 differentially expressed proteins across the three at-risk groups (NP, USneg, USpos). These corresponded to 233 proteins upregulated in USneg future progressors vs NP, 230 in USpos future progressors vs NP, and 202 in USpos future progressors vs USneg future progressors (Fig. 5a). To explore if these proteins reflect a coordinated biological program rather than isolated changes; we performed unsupervised k-means clustering on the 549 protein (*10*). This analysis resolved four coherent expression modules (C1-C4), each corresponding to a distinct immunologic program (Fig. 5b, S11). Cluster 1 (C1) proteins, which were enriched in USneg future progressors, showed upregulation of chemokines (CXCL3, CXCL5, CXCL6, CXCL11) together with signalling and immune regulatory proteins (MAP3K4, TLR2, TNFRSF13B, and ATRX), consistent with early activation of innate inflammatory pathway (*27–30*).

**Figure 5.**
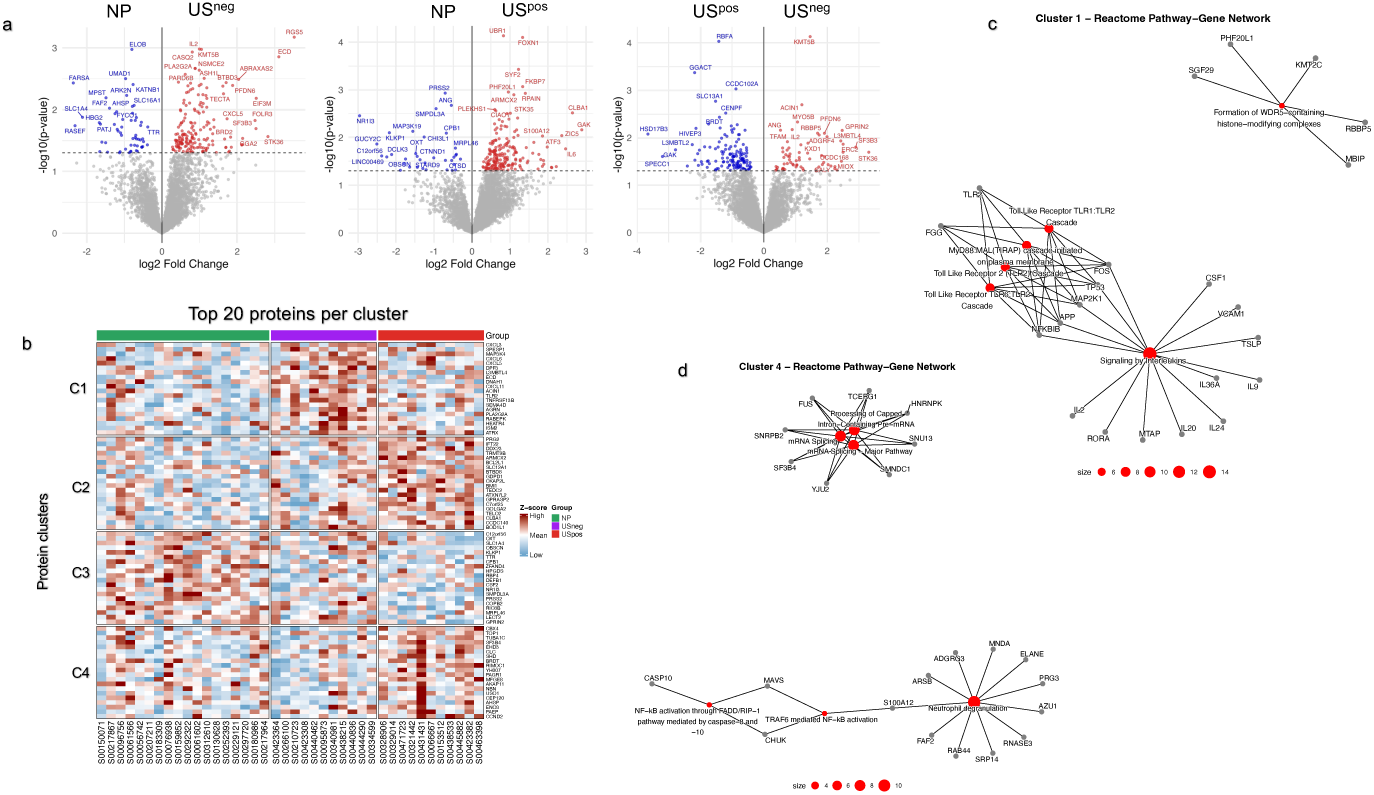
Differential plasma proteomic signatures and protein clustering across ACPA+ at-risk groups. a) Volcano plots show differential protein abundance between NP, USneg, and USpos groups based on limma analysis of Olink NPX values. Dotted lines indicate significance (p < 0.05) thresholds. b) Heatmap showing top 20 proteins within four protein clusters (C1-C4) resolved through unsupervised k-means clustering (k = 4) of z-scored normalized NPX values of 549 differentially expressed proteins. c) Functional enrichment of cluster-associated proteins using Reactome pathway analysis revealed immune activation and trafficking pathways (e.g., Toll-like receptor signalling, NF-κB activation) in Cluster 1 and d) metabolic, vesicle transport, and RNA processing pathways in Cluster 4.

Clusters 2 and 4 were more enriched in USpos future progressors, with most abundantly upregulated proteins included PRG2 – an epigenetic regulator linked with RA severity (*31*), IFT22 - a Rab-like GTPase involved in endosomal trafficking and cytokine receptor signalling (*32, 33*), and SF3B4, a component of mRNA splicing proteins, which has been associated with neutrophil-mediated immunity and immune infiltration (*34*). Together, these features are compatible with a shift towards a more inflammatory and tissue-directed phase. Cluster 3 proteins were most enriched in NP, including SLC1A4, a metabolic transporter suggesting altered metabolic pathways in auto-immune prone individuals, ZFAND4, and SMPDL3A, which has been recently shown to degrade cGAMP and inhibit cGAS-STING-mediated interferon signalling (*35, 36*) (Fig. 5b). Pathway enrichment analysis identified significant associations for cluster 1 and 4. Cluster 1 was enriched for interleukin signalling and Toll-like receptor cascades, consistent with early innate activation (Fig. 5c) and cluster 4 showed association for NF-κB signalling, neutrophil degranulation, and mRNA splicing, characteristic of adaptive and effector immune responses (Fig 5d), suggesting a transition from innate activation in USneg to effector programmes in USpos future progressors. Cluster 2 and 4 showed no significant pathway enrichment, likely due to limited functional annotation of novel proteins.

## Discussion

In this study, we provide a comprehensive single-cell atlas of peripheral immune dysregulation, comparing ACPA+ individuals with arthralgia who went on to imminently develop rheumatoid arthritis with those who did not progress. To understand the specific immune cell signatures that directly precede and follow the onset of early joint inflammation even before clinical arthritis occurs, we further stratified future progressors based on the presence or absence of subclinical synovitis on whole-body multi-joint ultrasound assessment. Previous longitudinal data has demonstrated that the majority of ACPA+ individuals with arthralgia first develop subclinical joint inflammation when progression to clinical rheumatoid arthritis is imminent (*37*). Our key findings reveal a biphasic model of immune activation in imminent RA. A robust IFN-I gene signature in CD14+ and IL1ß^hi^ monocytes, CD56^dim^ and mitochondrial-high NK cell subsets, and in a restricted subset of *GZMK*+ CD8+ T cells precedes the onset of subclinical joint inflammation in USneg future progressors. With the emergence of early subclinical joint inflammation in USpos future progressors, we identified an expansion of clonally expanded cytotoxic *GZMB*+ CD8+ T cells and IL1ß^hi^ classical monocytes. In contrast, NP did not engage this biphasic inflammatory programme, maintaining a quiescent immunological state. These stages were also accompanied by different proteomic profiles, further supporting distinct immune cell characteristics.

The proportions of these immune cell subsets also revealed phase-specific perturbations. The CD14+ classical monocytes proportions remained comparable between USneg future progressors and NP, however, USpos future progressors showed increased abundance of classical monocytes transitioning toward an IL1ß-high inflammatory phenotype. IL1ß-expressing monocytes have been described in multiomic studies of at-risk cohorts as well as in the synovial tissue of patients with early RA (*8–10, 38*), suggesting this population emerges early during synovial engagement and reflects initiation of myeloid driven early joint inflammation. Furthermore, these IL1ß^hi^ monocytes were also selectively enriched for CXC and CC chemokine ligands (*CXCL2*, *CXCL8*, *CCL2*, *CCL3*) in USpos future progressors. *CXCL2* and *CXCL8* drive neutrophil recruitment via *CXCR2* signalling, while *CCL2* and *CCL3* promote monocyte trafficking to the synovium (*27*). Concordantly, plasma proteomic profiling in USpos future progressors also revealed enrichment of neutrophil degranulation pathways, supporting early activation of neutrophil effector programmes during subclinical joint involvement. Neutrophils are among the earliest immune cells detected in RA synovium and their degranulation releases proteases, reactive oxygen species, and inflammatory mediators which contribute to the propagation of joint inflammation (*39*).

In parallel with enrichment of IL1ß^hi^ monocytes in USpos future progressors, we also observed a significant increase in proportions of terminally differentiated cytotoxic CD8+ T cells in USpos future progressors compared to both USneg and NP. Evidence from checkpoint-inhibitor induced arthritis indicates that IL1ß^hi^ monocyte/macrophage niches in synovium promote accumulation of cytotoxic CD8+ T cells through CXCL10-CXCR3 and CCL3-CCR1 signalling (*40*). Our data, therefore, suggest a similar myeloid-CD8 T cell axis may operate during the transition from ACPA-related autoimmunity to subclinical joint-centric inflammation preceding the onset of clinical arthritis. It is likely that IL1ß+ myeloid and cytotoxic CD8+ T-cell subsets in the peripheral blood at the stage of joint involvement may predominantly reflect recirculation of tissue-primed effector cells from the subclinically inflamed joints.

At a transcriptomic level, we observed an IFN-I gene signature in both CD14+/IL1ß+ classical and CD16+ non-classical monocytes, CD56^dim^ and mitochondrial-high NK cell subsets, and *GZMK*+ CD8 T cells only in USneg future progressors before the onset of subclinical inflammation. This signature was absent in NP as well as diminished in USpos future progressors after the onset of subclinical inflammation, suggesting a potential role in early systemic pro-inflammatory immune cell priming prior to joint inflammation. While IFN-1 signatures have been reported in at-risk RA cohorts (*10, 41, 42*), their temporal relationship to the onset of joint involvement remains unclear. Functional studies indicate that CD14+ classical monocytes are an early source and target of interferon signalling in ACPA+ individuals, driven by heightened innate immune responsiveness, including TLR-dependent activation, and PAD4-mediated histone citrullination (*9*). Recent data also implicates mucosal barrier disruption as a potential extra-articular trigger for interferon-primed HLA-DR^hi^ *ISG15*+ monocytes detected in the circulation in RA patients with periodontal disease (*43*). In longitudinal multiomic analyses, He et al., identified interferon-associated immune activation in individuals at risk of RA and the emergence of TNF+/IL1ß+ inflammatory monocytes and rising systemic cytokine levels at arthritis onset (*10*). However, these studies did not characterise immune events before and after the initial onset of joint inflammation in patients with imminent RA.

The terminally differentiated cytotoxic CD8+ T cell subsets also displayed transitional phenotype aligned with subclinical progression. These subsets were defined by expression of *GZMK*, *GZMB*, *PRF1*, *CX3CR1*, and *ZNF683* genes, consistent with a cytotoxic and tissue-resident effector phenotype. In USneg future progressors, the *GZMK*+ T cells showed limited clonal expansion but greater interferon-stimulated gene expression (*ISG15*, *MX1*, *IFI3*, *IFITM3*) compared with NP and USpos future progressors. In contrast, the USpos future progressors represented a clonally expanded *GZMB*+ T cell phenotype with diminished IFN-I activity. Recent single-cell transcriptomic studies have shown that GZMK+ CD8+ T cells represent a dominant tissue-associated effector population in inflamed synovium, and other tissues, that can recirculate while retaining a tissue-specific state (*44*). In line with this, the expression of *CXCR6* within *GZMK*+ CD8+ T cells in USneg progressors suggests a pool of interferon-high, cells primed for tissue entry, which appear to precede the emergence of clonally expanded cytotoxic *GZMB*+ CD8+ T cells at the stage of subclinical joint inflammation. Interestingly, we observed largely distinct *GZMK*+ and *GZMB*+ clonotypes in ACPA+ future progressors, suggesting preserved CD8+ T cell phenotypes, which are already established prior to the onset of subclinical and clinical arthritis. Thus, there may be an opportunity, in USneg individuals with imminent RA, to ‘intercept’ the tissue-centric expansion of pro-inflammatory *GZMK*+ CD8+ T cells that have been well described as a putative driver of tissue inflammation and damage (*44, 45*).

The plasma proteome showed evidence of an immune shift before and after the onset of joint inflammation in imminent RA. Although we did not directly detect elevated IFN-I proteins in the plasma of USneg future progressors, possibly due to a relatively small sample size, we detected elevated levels of innate immune sensors (TLR, MAP3K4), chemokines (CXCL3/5/6/11) and also a chromatic remodelling protein ATRX, which has not previously been described in RA. ATRX upregulation has been linked with ISG transcription through chromatin remodelling at IRF3-responsive promoters. Conversely, its loss impairs interferon induction and cytokine responses (*28, 29*). The detection of CXCL chemokines, TLR/MAP3K4 and ATRX proteins may suggest that in USneg progressors interferon pathways are transcriptionally primed.

Limitations of the study included the limited number of samples per group for plasma proteomic analysis, which may have reduced the ability to demonstrate inter-group differences in relatively low-abundance interferon signals (*46*). In addition, the absence of longitudinal paired blood and synovial tissue samples limited our ability to confirm whether interferon-primed monocytes and T-cell subsets subsequently traffic from peripheral blood into the joint and directly contribute to the very early subclinical synovitis seen in some patients with imminent RA.

A key strength of this study is the resolution of stage-specific immune programmes anchored to the earliest detectable joint inflammation in patients with imminent rheumatoid arthritis, demonstrating that immune signatures immediately preceding subclinical synovitis are distinct from those emerging once inflammation becomes detectable, even before clinical arthritis has occurred. The transition from interferon-driven systemic priming to tissue-associated cytotoxic and effector inflammatory programmes at joint involvement reveals potential targets for a therapeutic window that is key to the ultimate goal of arthritis interception and likely to attract growing therapeutic interest in the coming years.

## List of Supplementary Materials

Table S1

Figure S1 – S11

## Methods

### Study design

Study participants were selected from the Leeds CCP Cohort (*1*), a UK longitudinal observational study of individuals at risk of RA. All participants were anti-cyclic citrullinated peptide antibody (ACPA) positive with musculoskeletal symptoms but had no evidence of clinical arthritis. Forty high risk individuals, who had high levels of ACPA antibodies and inflammatory joint symptoms were selected for peripheral blood single-cell transcriptomic profiling. Participants were stratified initially by time to future progression to inflammatory arthritis. We identified individuals (n=22) who imminently progressed to RA within 6 months of their baseline visit and those who remained arthritis-free (non-progressors, NP =18) during follow up (up to 24 months). To investigate divergences in immune signatures before and after the initial development of joint tissue involvement, we further stratified imminent future progressors according to muti-joint ultrasound (US) evidence of subclinical synovitis at baseline. Ultrasound assessment was conducted in 33 joints and 10 tendons according to a standardised protocol described previously (*37*).

1. Ultrasound-positive (USpos) future progressors (n=11) demonstrated ultrasound power Doppler (PD) and Greyscale (GS) of 31 in one or more joints.
2. Ultrasound-negative (USneg) progressors (n=11) had no detectable subclinical synovitis in any joint on ultrasound.

Baseline peripheral blood mononuclear cells (PBMCs) and plasma samples underwent transcriptomic and proteomic profiling.

### Single cell RNA sequencing and library preparation

Cryopreserved PBMCs from individual study participants were thawed, washed, and resuspended in PBS containing 0.04% BSA to generate single-cell suspensions. Equal numbers of viable PBMCs from four individuals were combined to create 7 pooled suspensions containing approximately 60,000 cells (∼15,000 cells per participant). Some cell from the PBMCs suspension from each participant was reserved for bulk genotyping to enable single-nucleotide polymorphism (SNP)-based sample demultiplexing. Single-cell emulsions were generated using the Chromium Next GEM Single Cell 5′ Reagent Kit (v2 Dual Index, 10x Genomics) according to the manufacturer’s protocol (CG000331, Rev E). Briefly, single cells were encapsulated into gel bead-in-emulsion (GEM) droplets together with barcoded oligonucleotides containing unique cell barcodes and unique molecular identifiers (UMIs). Following reverse transcription and cDNA amplification, 5′ gene expression and V(D)J libraries were constructed according to standard 10x Genomics procedures. Libraries were sequenced on an Illumina NovaSeq 6000 platform to achieve a mean depth of approximately 20,000 reads per cell for gene expression libraries and 3,000 reads per cell for immune receptor libraries.

### Data analysis

ScRNAseq data were aligned to the GRCh38 human reference genome and quantified using CellRanger v6.0.1 (10x Genomics Inc.) Donor deconvolution was performed using genotype-imputed reference data. CellSNP-lite (v1.2.0) was used to call bi-allelic SNPs per cell, followed by donor assignment and sample demultiplexing using Vireo (v.0.2.3). For downstream analysis, raw count matrices from each of 7 pools were converted to Seurat object (v5.1.0) in R (v4.3.3). Cells were initially clustered at low resolution for quality control (QC). QC thresholds were applied to remove poor quality cells by visual inspection of the relevant metric distribution (Extended Table 1) and removal of unassigned donors by Vireo. Clusters were also removed if they contained high proportions of QC failure (guilt by association). All samples showing consistent QC metrics within pools were retained for further analysis. Putative doublets were called using Doublet detection was performed using Scrublet (v0.2.3), scDblFinder (v1.16.0), and scds (v1.18.0), and cells flagged by any tool or unassigned by Vireo were excluded. Ambient RNA contamination was corrected using SoupX (v1.2.2). Following doublet removal, the data were normalised using SCTransform (v2). PCA components were identified using RunPCA () and pools were integrated using reciprocal PCA integration (implemented in IntegrateLayer) to remove batch effect. Clustering was carried out using the Leiden algorithm with resolution of 1 to identify cell clusters. Annotation of cell clusters was initially guided by the COMBAT COVID-19 Cell Atlas (31), a large-scale single-cell reference dataset comprising immune cell subsets. Cell identities were initially assigned and by mapping cluster-specific gene expression profiles to reference labels in COMBAT dataset. COMBAT atlas annotations were stored in the metadata and guided final cell-type identification. We also integrated another independently processed scRNAseq dataset with existing Seurat object to increase ‘N’ of the study group. To enable unified downstream analysis, another layer of integration and downstream analysis steps were performed. Prior to integration, each dataset was independently normalised and variance-stabilised using SCTransform (v2). Objects were merged using Seurat’s merge() function. To correct for batch effect, cells were grouped by sample origin (e.g. run 1 vs. run 2, Supplementary Figure 1A) and the merged object was split accordingly. Each batch-specific object was then log-normalised and 2000 variable features were identified using FindVariableFeature(). Integration anchors were computed using FindIntegrationAnchors() with 2000 features and reciprocal PCA (rpca) as the reduction method. Batch correction and data integration were then performed using IntegrateData() with LogNormalise normalisation. Following integration, the integrated assay was used for downstream analysis. Data were scaled using ScaledData(), and PCA and UMAP embedding were computed using 30 or 50 principal components. Clustering was performed using FindNeighbors() and FindClusters() with Leiden resolution of 1. Final cell-type annotations were assigned using the FindAllMarkers() function to identify differentially expressed genes per clusters. These annotations were cross-checked with COMBAT atlas-derived labels stored in metadata to validate cell identity assignments.

### Differential gene expression analysis using DESeq2

Pseudobulb RNA-seq data from 31 immune cells subsets were analysed for differential gene expression analysis using DESeq2 (v 1.46.0) (*47*). Genes with total count of <10 were excluded as lowly expressed genes. Group comparisons were performed between US^neg^ vs NP, US^pos^ vs NP, and US^neg^ vs US^pos^. Normalization used DESeq2’s median-of-ratios method, and Wald tests were applied with Benjamini–Hochberg correction (*FDR* < 0.05, log₂FC > 1). Volcano plots were generated with ggplot2 and ggrepel, highlighting the top 20 significant genes per contrast.

### Flow cytometry validation of intracellular MX1 and ISG15 expression

Intracellular protein expression of MX1 and ISG15 in CD14+ classical monocytes and CD56^dim^CD16^hi^ NK cells was validated by flow cytometry using cryopreserved PBMCs collected during the same clinical visit as those used for scRNA sequencing. PBMCs were thawed at 37 °C in a water bath and washed twice in PBS + FBS. One million cells in 1 ml suspension were stained with Fixable Viability Dye eFluor 780 (Invitrogen/eBioscience), and subsequently Fc block (BD Biosciences) was applied to prevent nonspecific antibody binding, as per the manufacturer’s instructions. Cell surface staining was performed for 30 min in the dark using a multicolour antibody panel containing anti-CD14, anti-CD16, anti-HLA-DR, anti-MERTK, anti-CD3, and anti-CD56. After surface staining, cells were fixed and permeabilized using the FIX & PERM Cell Permeabilization Kit (Invitrogen), according to the manufacturer’s protocol. Intracellular staining was then performed with anti-MX1 (AF488) and anti-ISG15 (PE) antibodies, along with matched isotype controls. Stained samples were acquired on a CytoFLEX LX flow cytometer (Beckman Coulter), collecting approx. 50,000 events per sample and data were analyzed using CytExpert v2.6 software. Fluorescence compensation was applied using matrices generated from single-stained controls to correct for spectral spillover. Median fluorescence values for *MX1* and *ISG15* were obtained from the statistics table in CytExpert v6 for gated HLA-DR^+^CD14⁺CD16⁻ monocytes and CD56^dim^CD16^hi^ NK cells (Supp Table 2). Median values were used for statistical analysis, and differences in protein expression across three anti-CCP⁺ at-risk groups (NP, US^neg^, US^pos^) were assessed using one-way ANOVA in R (v4.4.1).

### T cell clonotype and transcriptional program analysis

To assess TCR repertoire and functional states of CD8+ T cells, we subsetted the Seurat object to CD8+, MAIT, and γδT cells based on canonical marker expression (CD8A, CD8B, KLRB1, TRDC). Raw counts were normalized using NormalizeData() function (normalization.method=“LogNormalize”, scale.factor=1e4) function in Seurat. Highly variable genes (HGVs) were identified with FindVariableFeatures(nfeaturs = 3000) and gene-wise z-scaling was applied with ScaleData. PCA analysis was performed on the scaled HVGs for dimensionality reduction and based on elbow point of the variance curve, 30 PCs were retained for downstream analyses, including nearest-neighbour graph construction (knn), UMAP, and clustering. To correct for inter-sample and library-preparation batch effects, a secondary correction was performed using Harmony. RunHarmony() was applied to the first 30 principal components derived from the RNA assay (reduction = “pca”, dims = 1:30), grouping by donor ID (group.by. vars = “vireo_geno_donorID”) and including chemistry or pool when applicable. Harmony embeddings were used to assess standard neighbourhood-mixing checks (e.g., iLISI/kBET on k-NN graphs, when computed) showed no deterioration of lineage separability; the lineage-aware embeddings used downstream remained topologically stable before vs after correction.

### CD8+/MAIT/ γδ T cell lineage aware embeddings and TCR repertoire analysis

To emphasize immune lineage identity over activation state, PCA and UMAP were recomputed on a fixed marker panel comprising canonical markers for pan-T (CD3D/E/G, CD2, PTPRC), CD4 (CD4), CD8 (CD8A/B), B cells (MS4A1, CD79A/B, BANK1, CD74), NK cells (KLRD1, KLRK1, NCR1, FCGR3A), MAIT (KLRB1, ZBTB16, RORC, CXCR6, IL7R), γδ T cells (TRDC, TRDV/TRGV), and Tregs (FOXP3, IL2RA, IKZF2). UMAP was computed using the utow package with cosine metric, n.neighbors = 15, min.dist ≈ 0.18, and a fixed seed. This shared embedding was reused across overlays for consistency. A ternary label (“Tclass3”) was applied to distinguish CD8⁺, MAIT, and γδ T cells. Cells in the CD8⁺/MAIT/γδ compartment were reclustered using a 20-PC space (FindNeighbors, dims = 1:20), followed by Louvain clustering (FindClusters, resolution = 0.4). A parallel k-means clustering (k = 4) was performed on the same PC space and stored for comparison. Kernel-density contours (stat_density_2d, 6 levels) were overlaid to illustrate cluster separation on the UMAP.

To assess functional states, a 55-gene panel was curated to represent cytotoxicity, cytokine signaling, naïve/memory phenotype, interferon response, activation/exhaustion, residency, and transcriptional regulators (TCR genes excluded). Z-scored expression (ScaleData, clipped at ±2) was visualized using ComplexHeatmap with genes ordered by cluster of peak expression. For group-wise program summaries, log-normalized expression was first averaged per donor per clone class, then donor-averaged across groups with equal weight. Columns were ordered by clone-size class (Singleton → Small → Medium → Large), and rows were clipped z-scores. Cluster and group compositions were visualized using 100% stacked bar plots and boxplots. Donor-level statistics were tested using Wilcoxon or Kruskal–Wallis tests with Benjamini–Hochberg FDR control.

Differential gene expression (DGE) was tested in two comparisons: (i) CD8⁺ T cells from progressors (USneg and USpos combined) vs NP, and (ii) large clones (≥20 cells) vs non-large (singleton/small/medium). DGE was performed with FindMarkers (Wilcoxon, min.pct = 0.10, logfc.threshold = 0). Significant genes were defined by FDR < 0.05 and |log₂FC| ≥ 0.25. Volcano plots used consistent color coding and ggrepel-based gene labeling. Clonotypes were defined primarily by TRB CDR3 amino-acid sequence (Fairfax et al.) (*23*), with TRA used for validation but not required. Clone sizes were calculated per donor and classified as Singleton (1), Small (2–4), Medium (5–19), or Large (≥20) in line with recent RA study (*48*). Diversity per donor was calculated from clone frequencies pi as Shannon entropy H = −∑ pi ln pi (natural log). Pielou evenness J = H / ln(S), where S is the number of distinct clones. The “clonality” index in Figure 2e was 1 − H/ln(S), approaching 1 when one/few clones dominate and 0 for even repertoires. TRBV and TRBJ usage was parsed from standardized V/J gene columns and visualized via chord diagrams. Links were filtered to retain the top ∼400–800 most frequent V–J pairs per group to ensure readability. Diversity per donor was calculated from clone frequencies pi as Shannon entropy H = −∑ pi ln pi (natural log). Pielou evenness J = H / ln(S), where S is the number of distinct clones. The “clonality” index in Figure 2e was 1 − H/ln(S), approaching 1 when one/few clones dominate and 0 for even repertoires. TRBV and TRBJ usage was parsed from standardized V/J gene columns and visualized via chord diagrams. Links were filtered to retain the top ∼400–800 most frequent V–J pairs per group to ensure readability.

### Plasma proteomics

Plasma proteomics profiling was performed using the Olink Explore 5K panel (v 7.2.1), to quantify approximately 5000 protein targets in at-risk cohort (n=40) collected during baseline visit. These samples formed a small subset of large cohort of profiled samples. Normalized Protein Expression (NPX) values and metadata for all samples were accessed from the central Olink dataset in Parquet format. Samples corresponding to the study cohort were extracted to generate a subset dataset for downstream analysis in R (v4.4.1). Initial quality control (QC) and data visualization were performed following the Olink Analyze Vignette data analysis guidelines, using the OlinkAnalyze package for descriptive statistics, data structure visualization, and distribution assessment. QC filtering excluded proteins or samples failing the SampleQC or AssayQC criteria, as well as internal extension and amplification controls (EXT1, INC1, AMP1). Differential protein expression analysis was performed using limma package (v3.62.2) (*49*) on Principal Componenet (PC)-normalised, log-2 transformed-NPX values. Linear models were fitted to compare protein abundance across NP, USneg, and USpos groups. Pairwise contrasts (USneg vs NP, USpos vs NP, USpos vs USneg) were tested, and empirical Bayes moderation was applied to stabilize variance estimates. Proteins with nominal p < 0.05 were considered differentially expressed for downstream analysis. PCNormalise NPX values of significant proteins were converted to Z-score-scaled by row (per protein) and subjected to unsupervised k-means clustering. Optimal number of protein clusters within-cluster dispersion across k = 1-10 clusters were identified by both gap statistics (clusGap, cluster v2.1.8.1) and elbow method. Cluster assignments were used to define protein modules, which were visualized using the ComplexHeatmap package (v2.22.0). For pathway enrichment analysis, proteins within each k-means cluster were converted to Entrez Gene IDs and analysed for pathway over-representation using Reactome pathway enrichment implemented in the ReactomePA package. Pathways with FDR < 0.05 were visualized using gene-pathway network diagrams generated with cnetplot.

## Data Availability

Data supporting the findings of this study are available from the corresponding author upon reasonable request.

## Acknowledgments

We would like to thank Jacqueline Nam, Kate Harnden, Laurence Duquenne, Andrea Di Matteo, Helen Sugden who contributed to patient recruitment. Tim Hardy, Nicola Burnett, Georgina Young, Oksana Danyliuk, Katie Mbara for study co-ordination. Aamir Aslam for support and advice with flow cytometry. Richard Wakefield and Kate Smith for performing ultrasound assessments.

## Funding

This study was supported by grant funding awarded to KM from Arthritis UK. OLink HT was supported through an investigator-initiated collaboration with Astra Zeneca.

## Author contribution

The study was conceived by K.M., P.M., M.H.B., and D.N. Participants were clinically evaluated by KM, and the patient cohort was curated by K.M. and F.T. Plasma samples were curated, and proteomics data were generated by S.S. Experimental works were performed by F.T., P.M., K.A., S.M., D.M. Data analyses were performed by F.T., P.M., K.A., S.S., W.Y. Senior oversight was provided by K.M., M.M., H.M., M.H.B., D.N., B.P.F., and P.E. Figures and tables were developed by F.T., and K.A. The manuscript was written by F.T. and K.A. All authors contributed to editing and approved the final manuscript. All authors declare that they have no competing interests.

---

**Extended Table 1.** Quality control thresholds applied per sample pool.

| Sample pool | Minimum number of features | Minimum number of counts | Maximum mitochondrial percentage |
| --- | --- | --- | --- |
| L1 | 600 | 1500 | 8 |
| L2 | 500 | 1000 | 9 |
| L3 | 400 | 900 | 9 |
| L4 | 700 | 1500 | 8 |
| L5 | 500 | 1250 | 8 |
| L6 | 500 | 1000 | 9 |
| L7 | 700 | 1250 | 8 |

## Notes

### Competing Interest Statement

The authors have declared no competing interest.

### Funding Statement

This study was supported by grant funding from Arthritis UK. Olink HT proteomic analyses were supported through an investigator-initiated collaboration with AstraZeneca. The authors and their institutions did not receive any additional payments or services from third parties for any aspect of the submitted work.

### Author Declarations

The Research Ethics Committee of the UK National Health Service gave ethical approval for this work (reference numbers 06/Q1205/169 and 17/YH/0177). All participants provided informed consent.

