## Supplementary data for "Type-1 interferon-driven innate and *GZMK*+ CD8 T cell activation precedes subclinical joint inflammation when rheumatoid arthritis is imminent"

Tariq et al., 2026

| **Characteristics** | | **Non-progressors (NP)** | **US-negative (US^neg^) progressors** | **US-positive**  **(US^pos^) progressors** |
| --- | --- | --- | --- | --- |
| Total (n) | | 18 | 11 | 11 |
| Age at baseline visit, (years) | | 54 (49.5–64.0) | 54 (42–58) | 46 (38–63) |
| Sex, n (%) | Female | 11 (61.1%) | 7 (63.6%) | 7 (63.6%) |
|  | Male | 7 (38.9%) | 4 (36.4%) | 4 (36.4%) |
| Anti-CCP, U/mL (median [IQR]) | | 66.7 [24.5–226] | 300 [67–300] | 300 [80.8–300] |
| CRP, mg/L (median [IQR]) | | 4 [1.26–5.35] | <5 (<5) | 4 [4–4] |
| ESR, mm/h (median [IQR]) | | 8 [3–11] | 12 [10.5–23] | 18 [6–32.5] |
| HLA+ status, n (%) | Yes | 18 (100 [55.6% DRB1, 44.4% other]) | 6 (54.5) | 7 (63.63) |
|  | Unknown | 0 | 5 (45.45) | 4 (36.36) |
| Smoking n, (%) | Yes | 11 (61.1%) | 7 (63.6%) | 6 (54.5%) |
|  | No | 7 (38.9%) | 3 (27.3%) | 4 (36.4%) |
|  | Unknown | NA | 1 (9.1%) | 1 (9.1%) |
| Subclinical synovitis (PD/GS >= 1) on ultrasound imaging (n) | | No | No | Yes (11) |
| Time to progression, months (median [IQR]) | | NA | 3.5 [2.3–6] | **2.5 [1.5–3.5]** |

**Table S1. Characteristics of ACPA+ at-risk RA groups.** This table summarizes key demographic, serologic, genetic, serum inflammatory and joint ultrasound characteristics of study participants in the three at-risk groups. Data are presented for non-progressors (NP), ultrasound-negative (USneg) future progressors, and ultrasound-positive (USpos) future progressors. Continuous variables are reported as median [interquartile range] and categorical variables as n (%). Age refers to the participant’s age at baseline. Anti-CCP values are shown in U/mL, and CRP (C-reactive protein) and ESR (erythrocyte sedimentation rate) reflect systemic inflammation, with CRP values below detection limit (<5 mg/L) indicated. HLA+ status denotes the presence of RA-associated alleles, with subtypes (DRB1 vs other) shown where available; “Unknown” indicates missing data. Smoking status is reported as Yes/No/Unknown. **Subclinical synovitis was assessed by musculoskeletal ultrasound and defined by the presence of power Doppler (PD) and/or greyscale (GS) synovitis.** Time to progression represents months from baseline to clinically apparent RA in progressors and is not applicable for NP.

Experimental batches

Seurat clusters

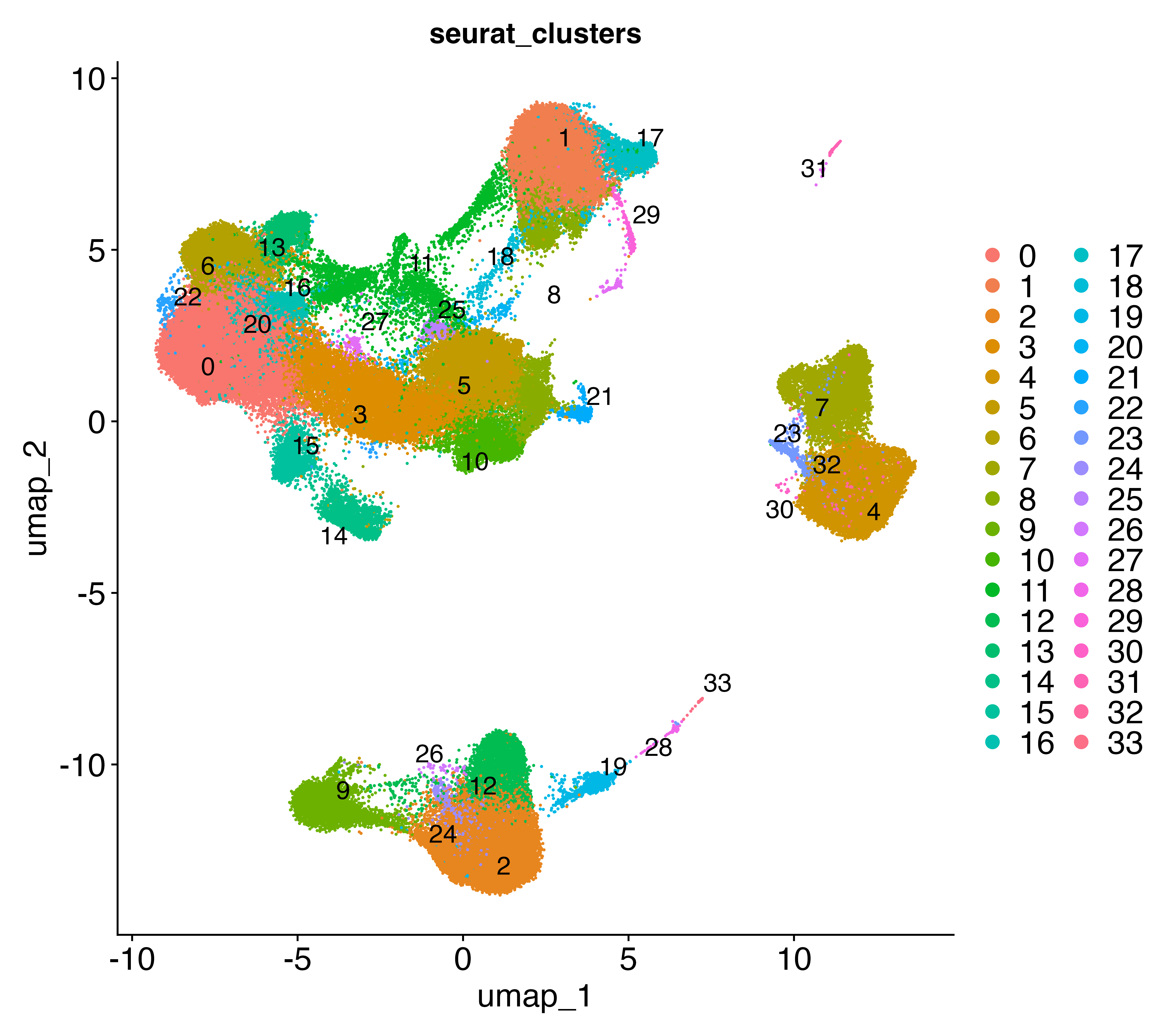

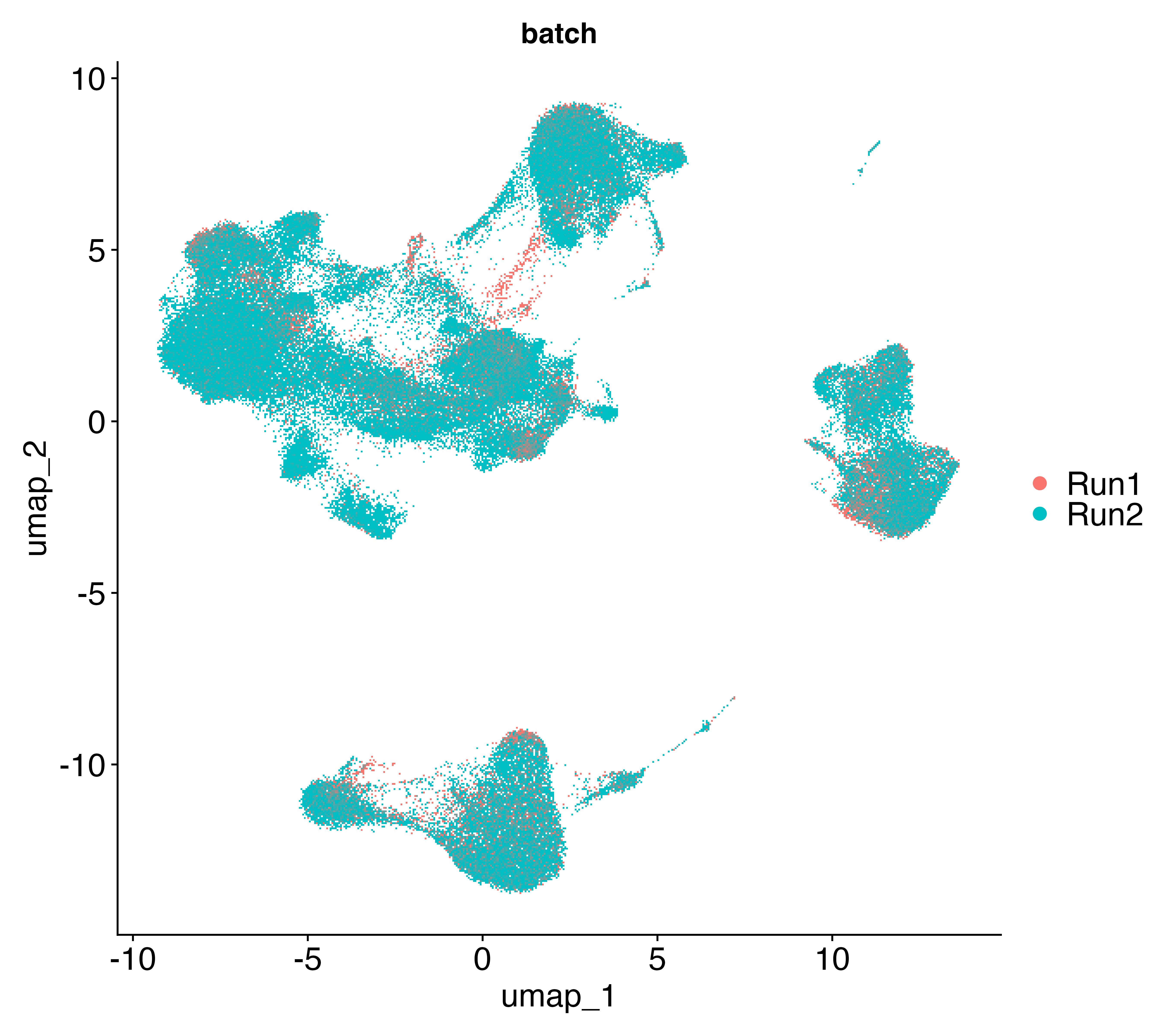

a

b

Samples

Groups

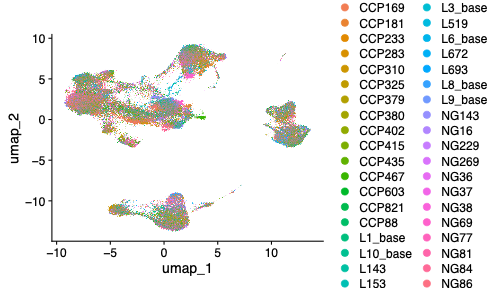

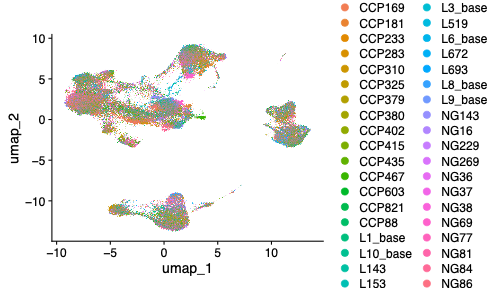

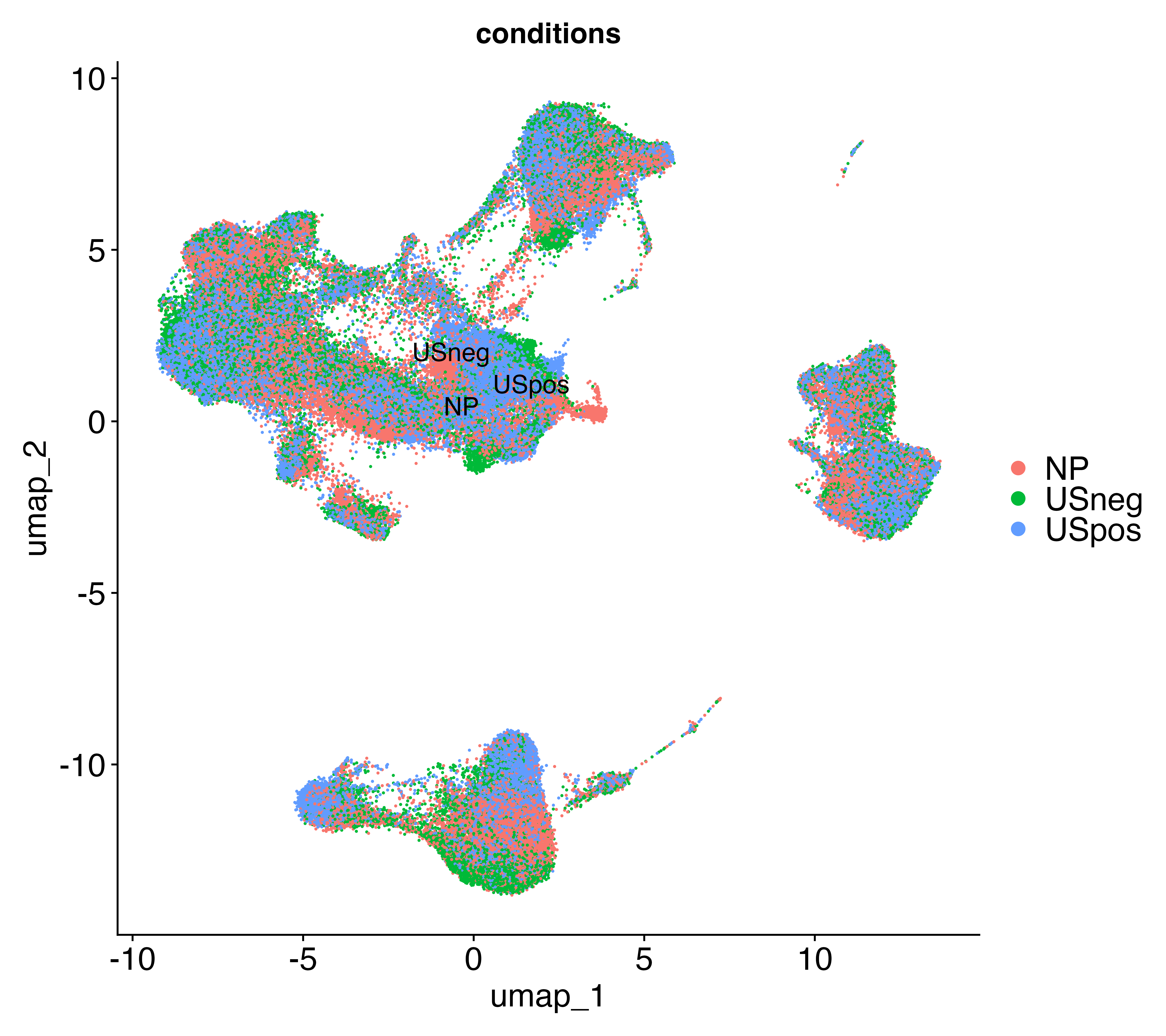

c

d

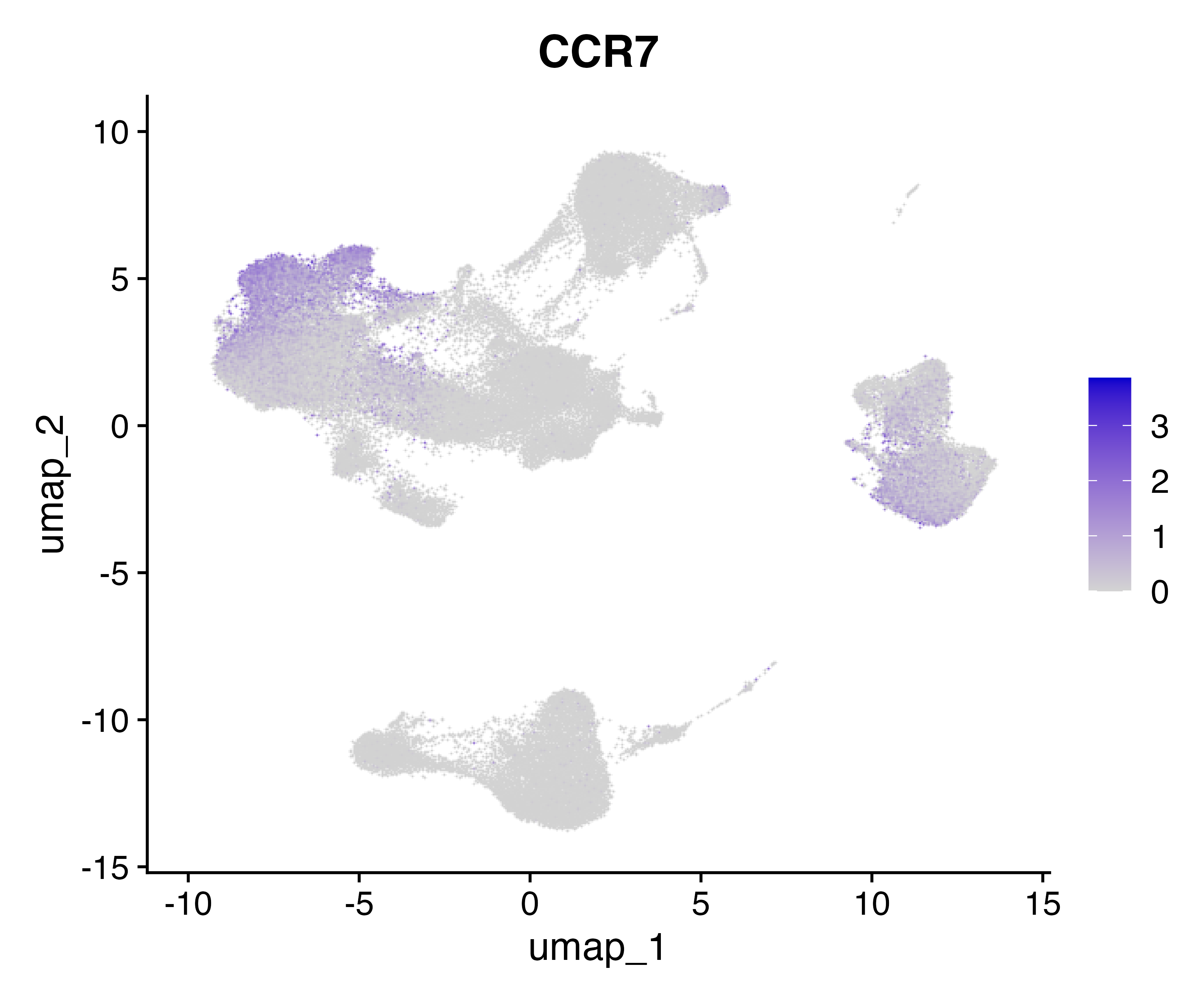

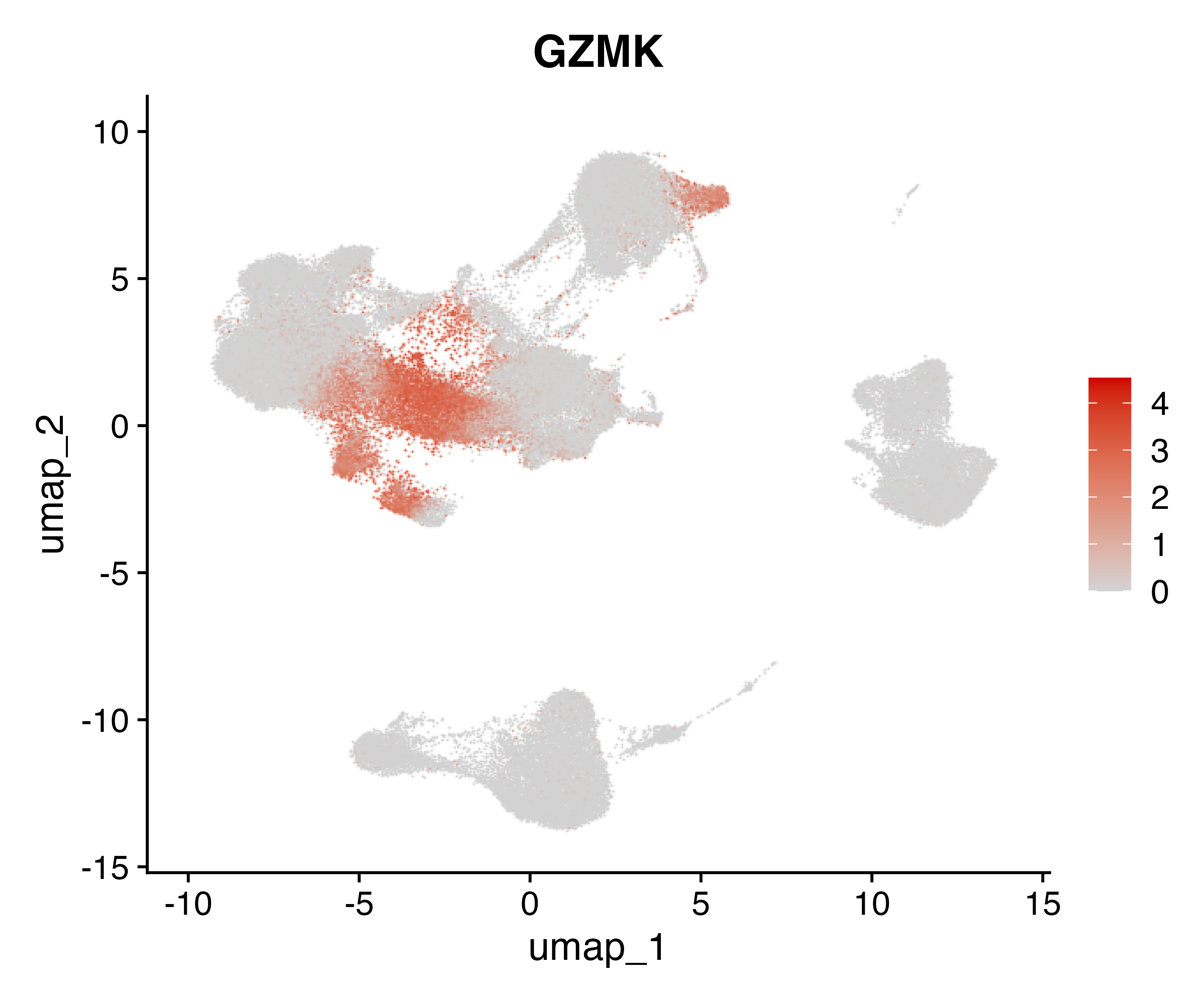

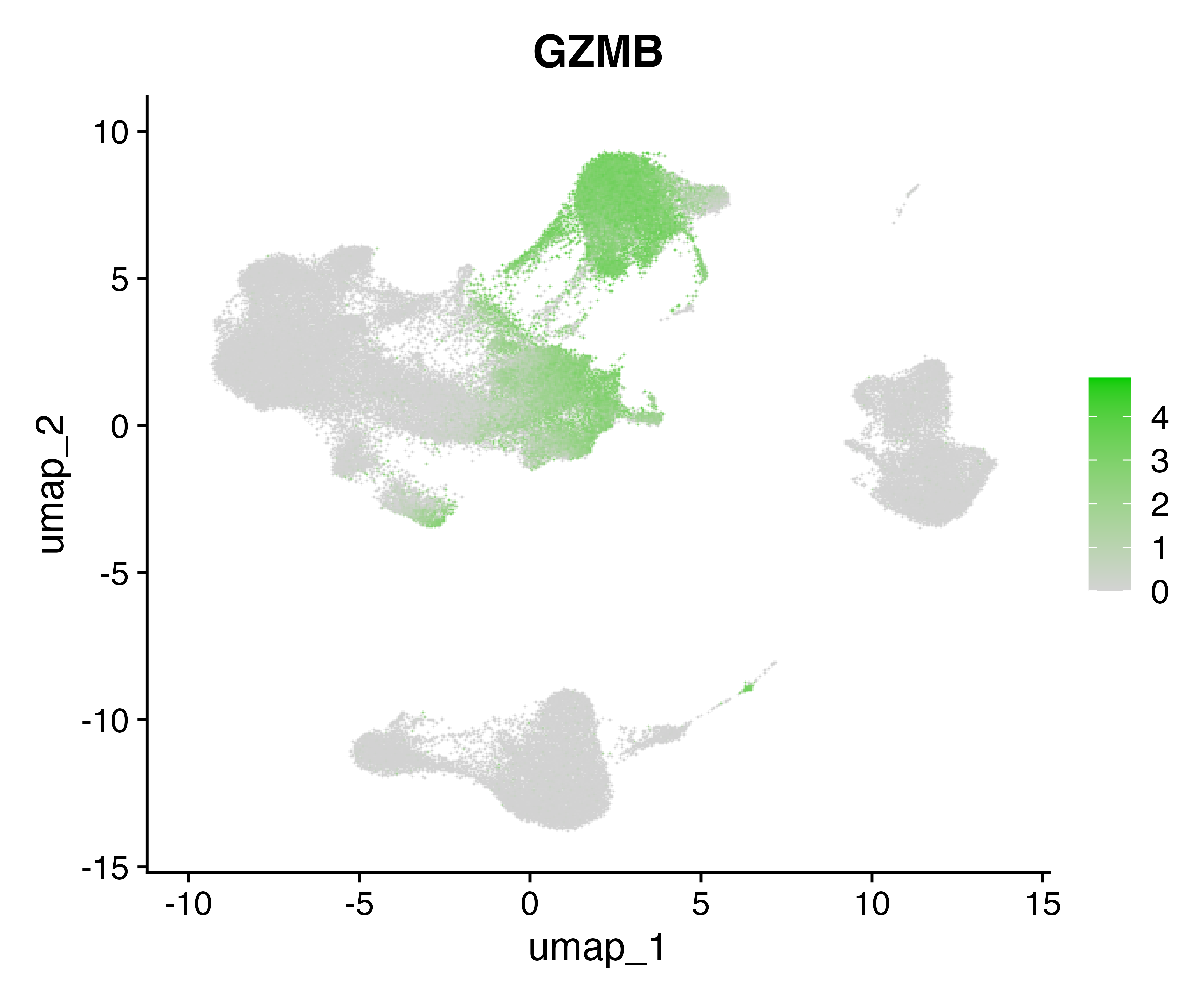

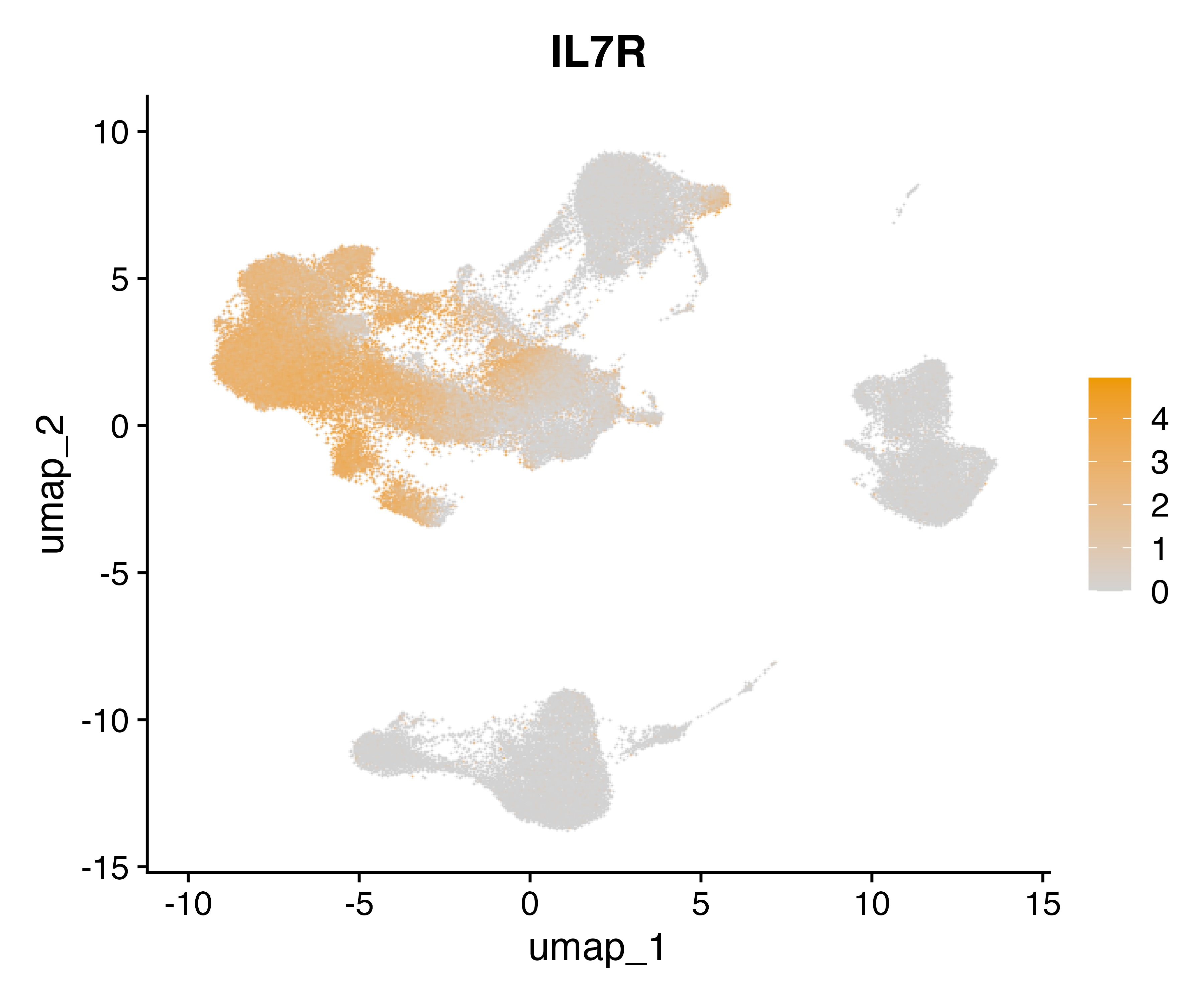

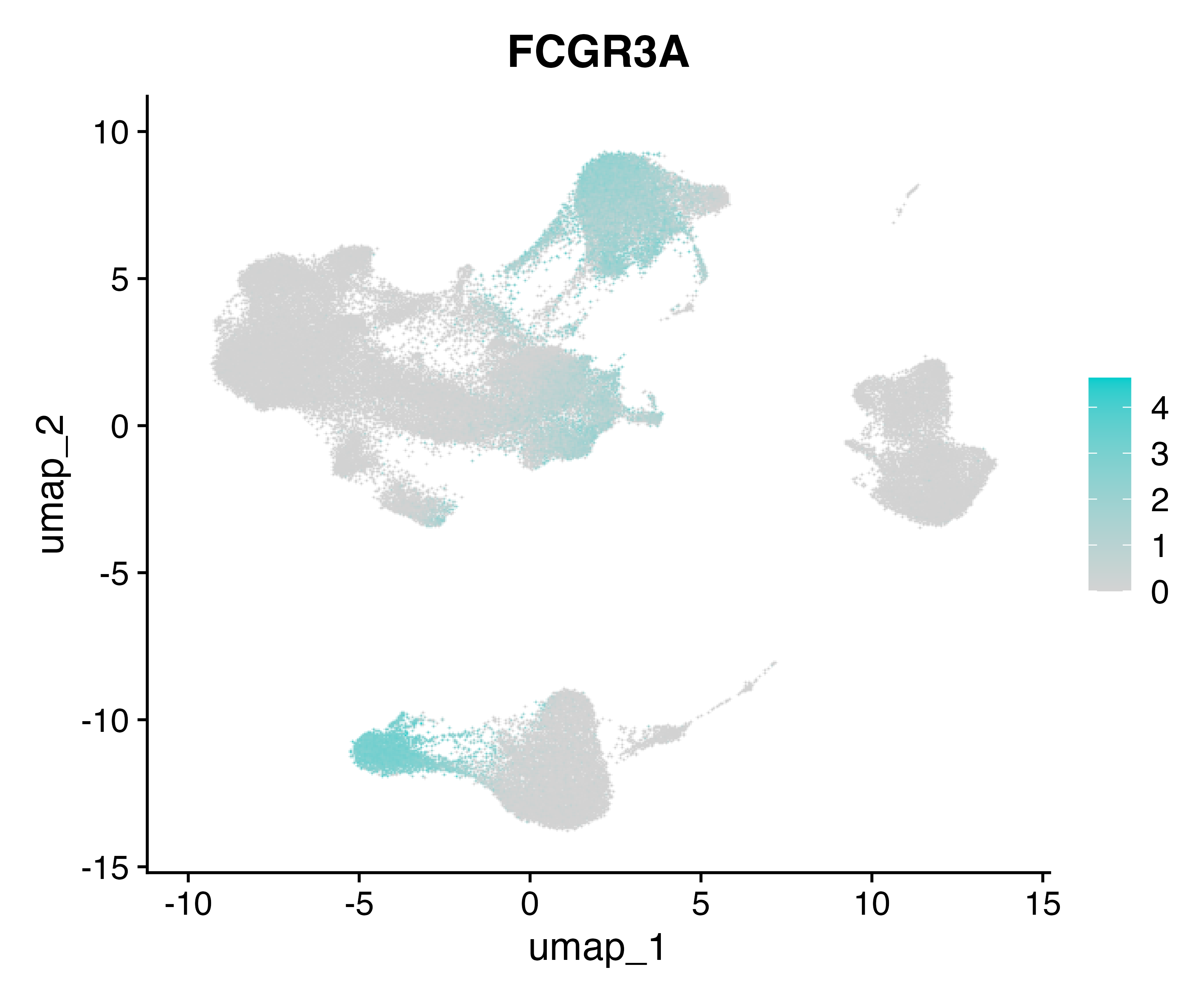

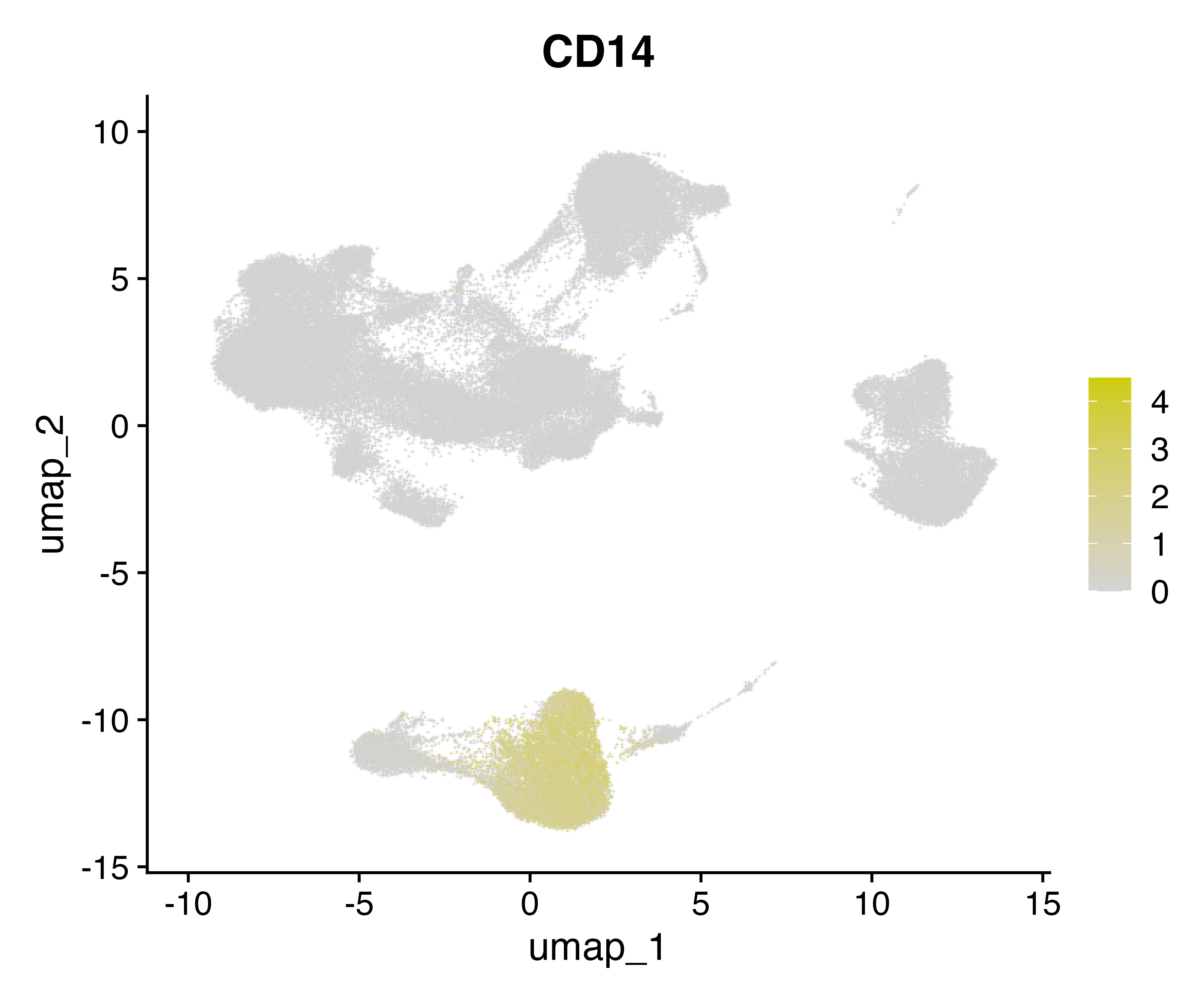

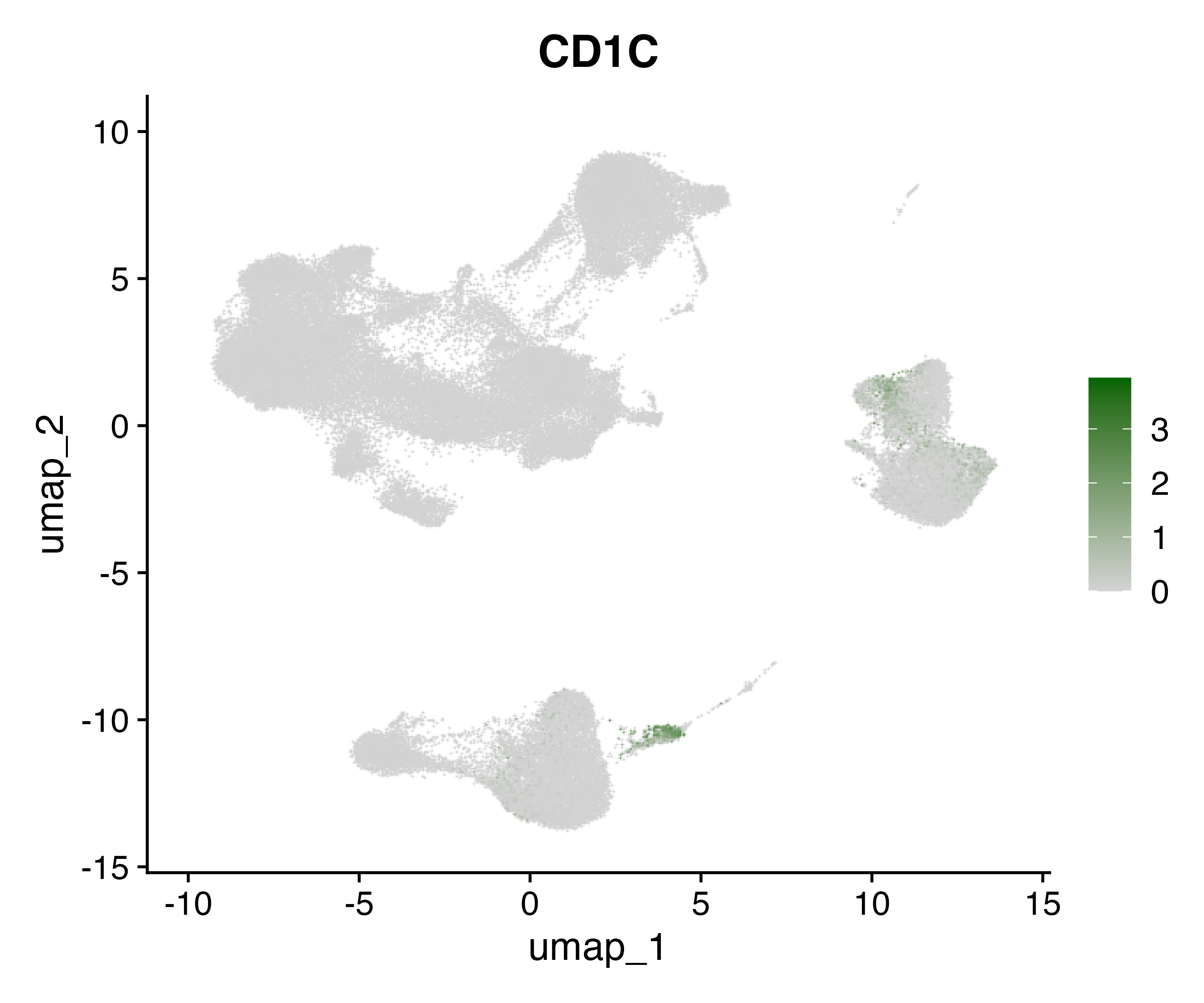

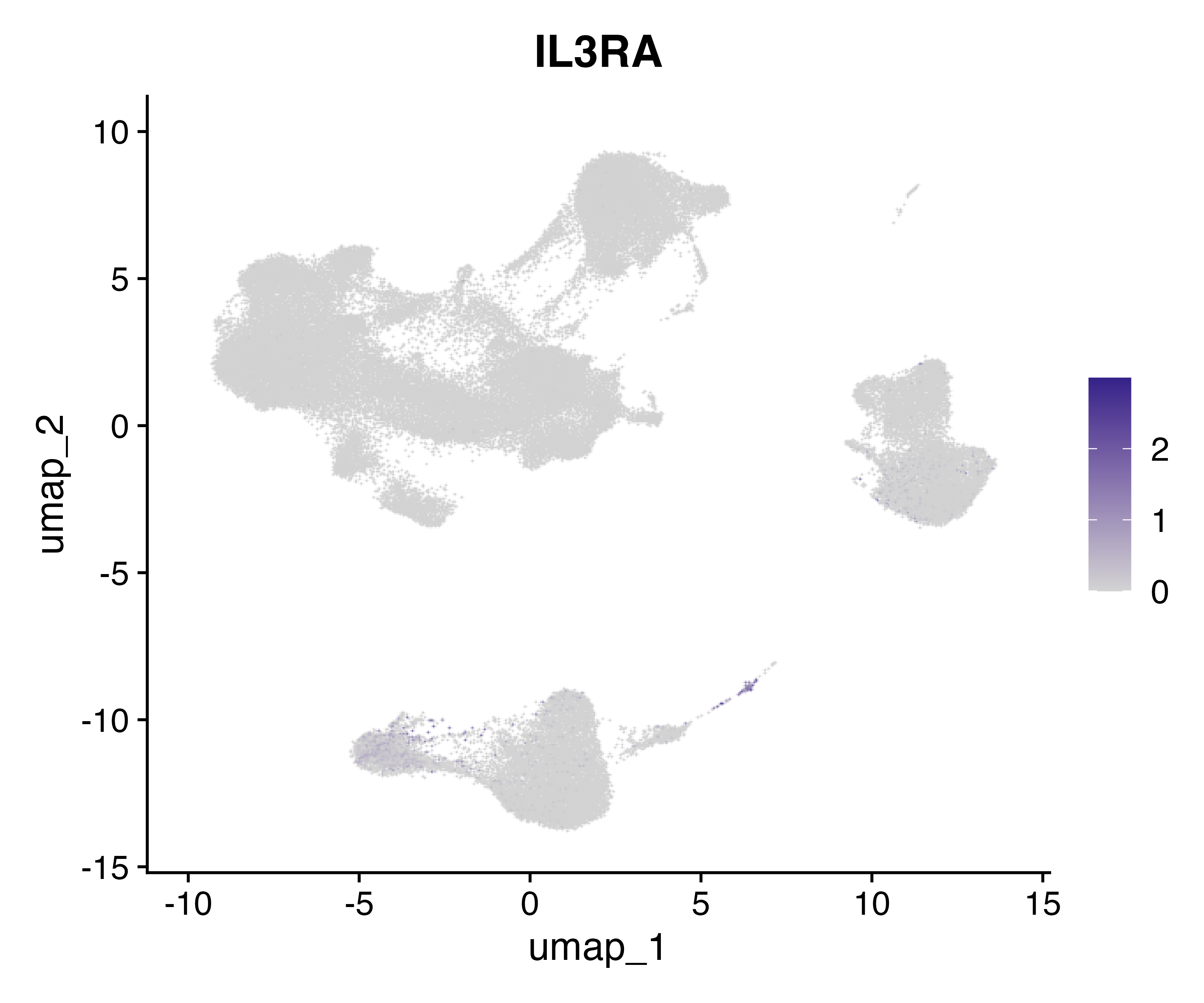

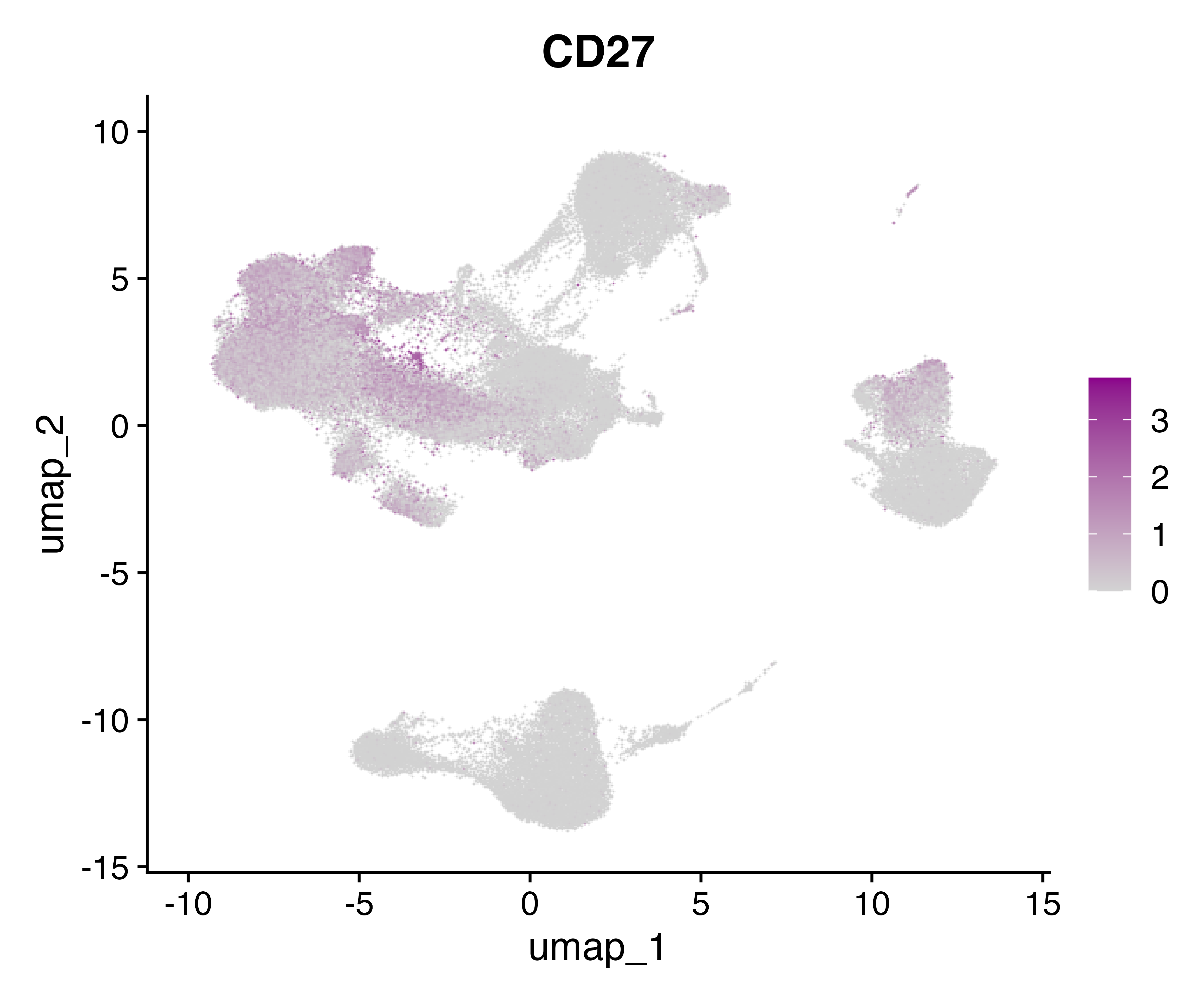

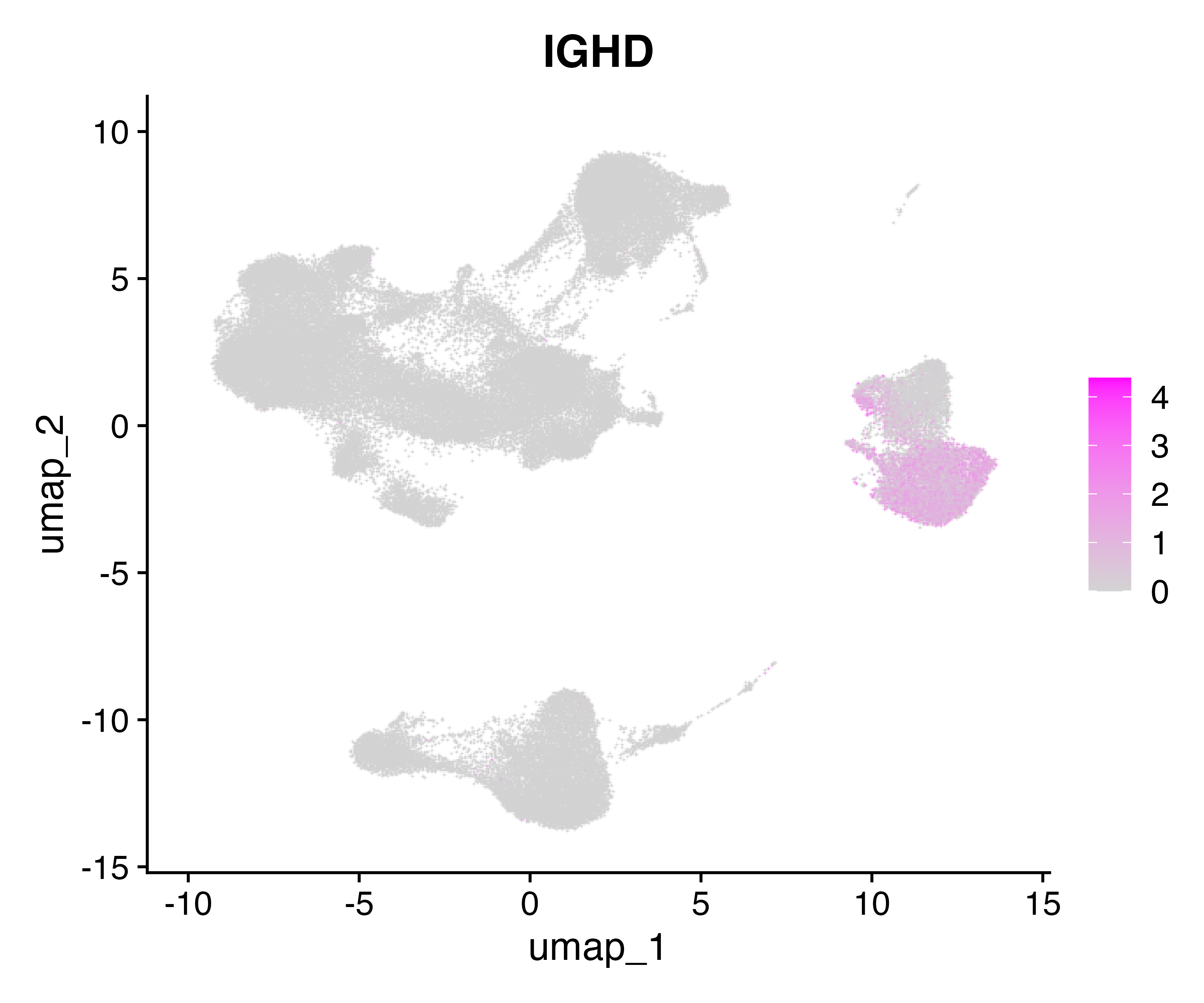

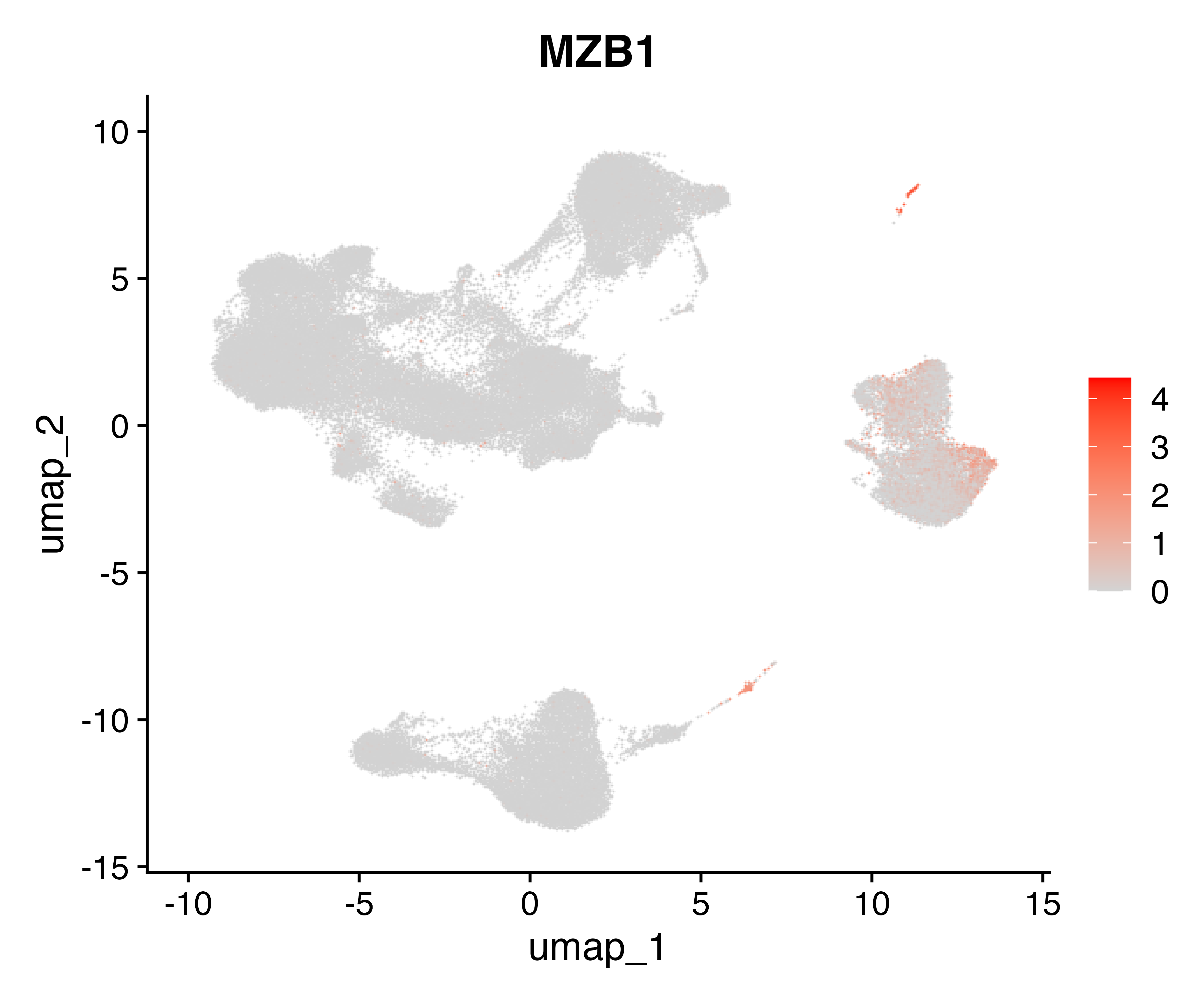

e

**Figure S1. UMAP visualization of scRNAseq dataset.** a) Unsupervised clustering of integrated scRNAseq data, showing transcriptionally distinct clusters identified using Seurat's Leiden algorithm (resolution = 1). Each colour represents a unique cluster. b) Distribution of cell clusters coloured by three at-risk groups: non-progressors (NP), ultrasound-negative future progressors (USneg), and ultrasound-positive future progressors (USpos). c) Cells coloured by different scRNAseq runs, confirming successful batch effect correction after reciprocal PCA-based integration using Seurat v5. Cells from different batches are well mixed across clusters, indicating effective integration. d) Expression of canonical **marker genes** used for cell cluster identification and annotation plotted on the integrated UMAP, including **CD3E** (T cells), **GZMK** (cytotoxic T cells), **GZMB** (cytotoxic lymphocytes), **IL7R** (naïve/memory T cells), **FCGR3A** (NK/monocytes), **CD14** (classical monocytes), **GNLY** (NK and cytotoxic T cells), **IL3RA/CD1C** (dendritic cells cells), **CD27** (memory T/B cells), **MS4A1** (B cells), and **NKG7** (cytotoxic effector cells).

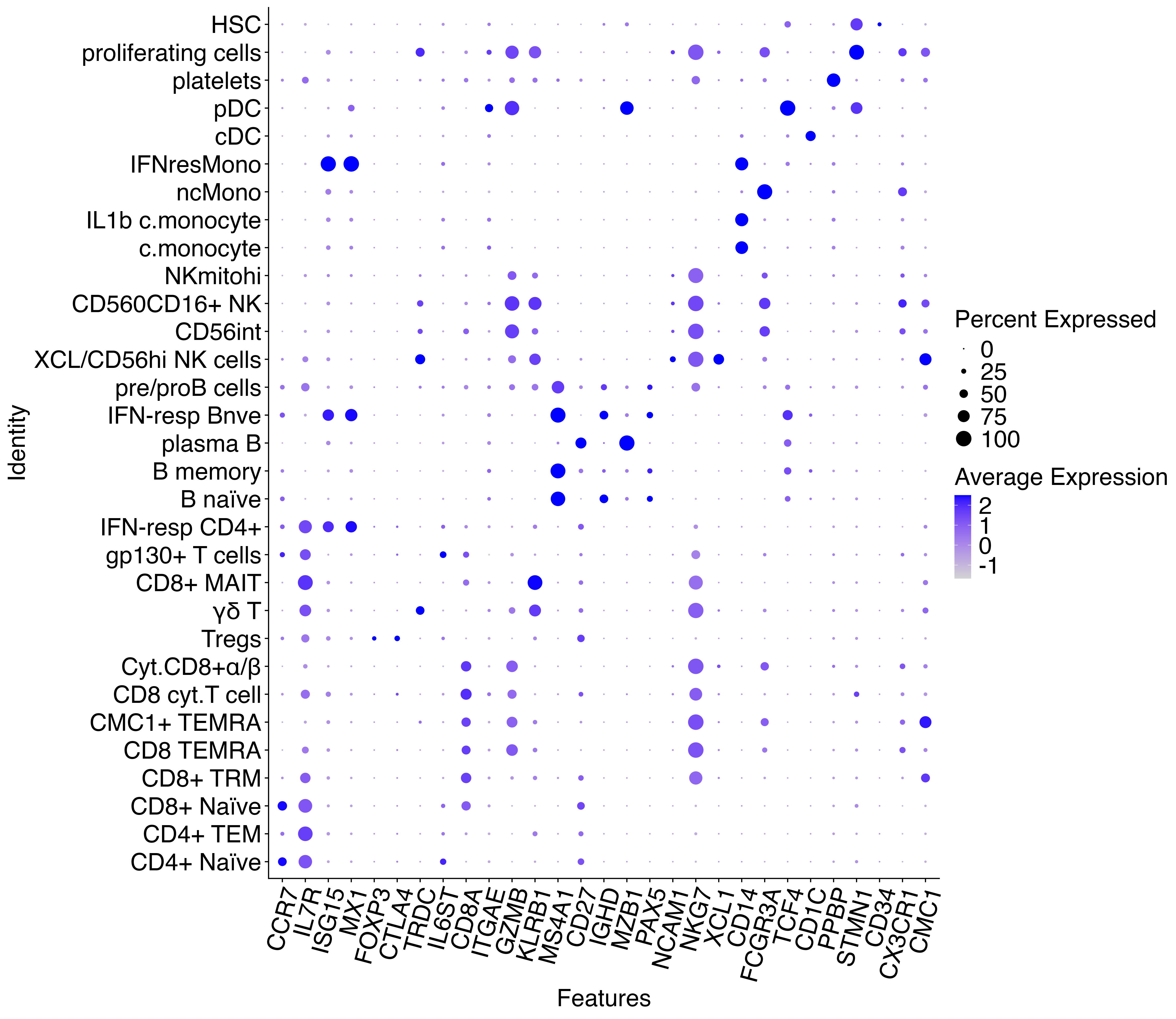

**Figure S2. Expression of canonical marker genes across identified immune cell clusters.** Bubble plot showing average expression and percentage expression of key lineage marker genes across all Seurat-identified cell subsets. Dot colour represents the scaled average expression level, while dot size indicates the percentage of cells within each cluster expressing a given gene.

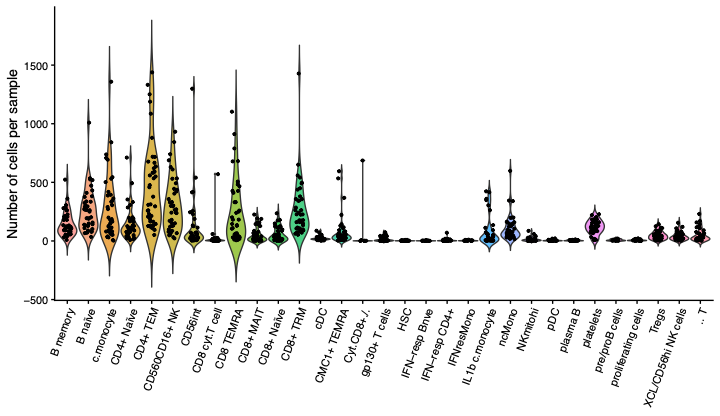

**Figure S3. Distribution of cell counts per donor across cell subsets.** Violin plots showing the number of cells per donor for each annotated immune cell subset identified by single-cell RNA sequencing. Each point represents an individual donor, and violins indicate the distribution of cell counts across donors for each subset.

a

CD4+ T cells

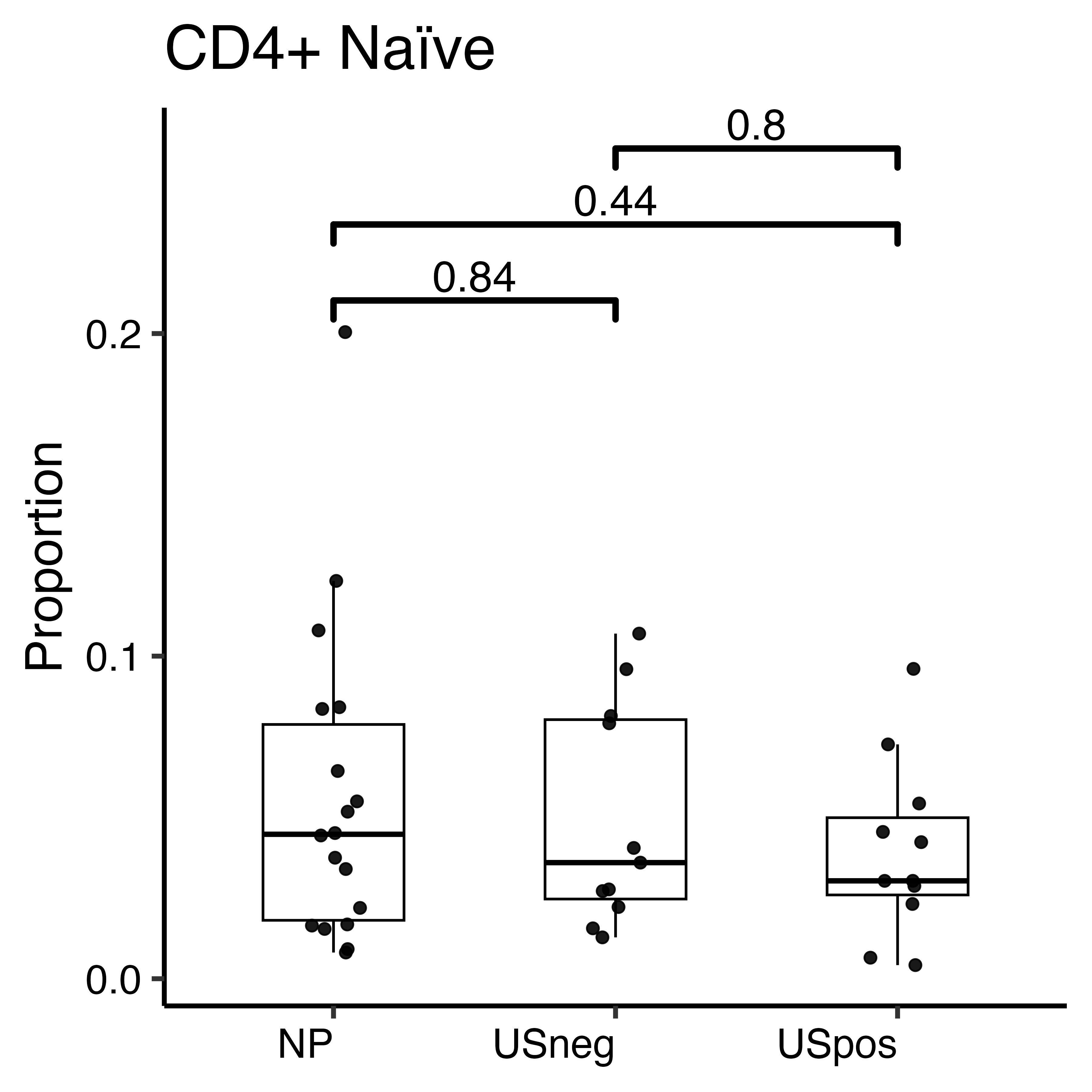

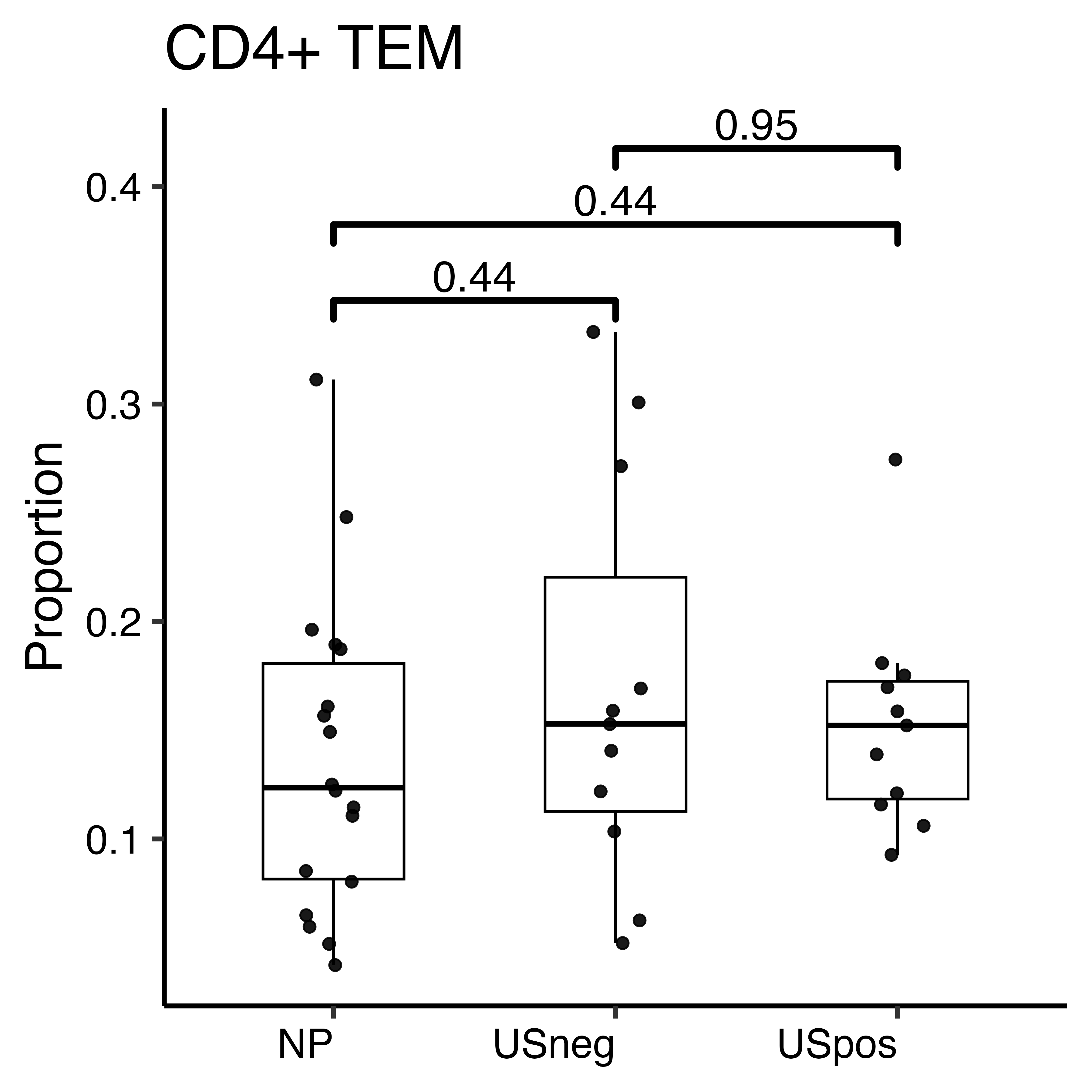

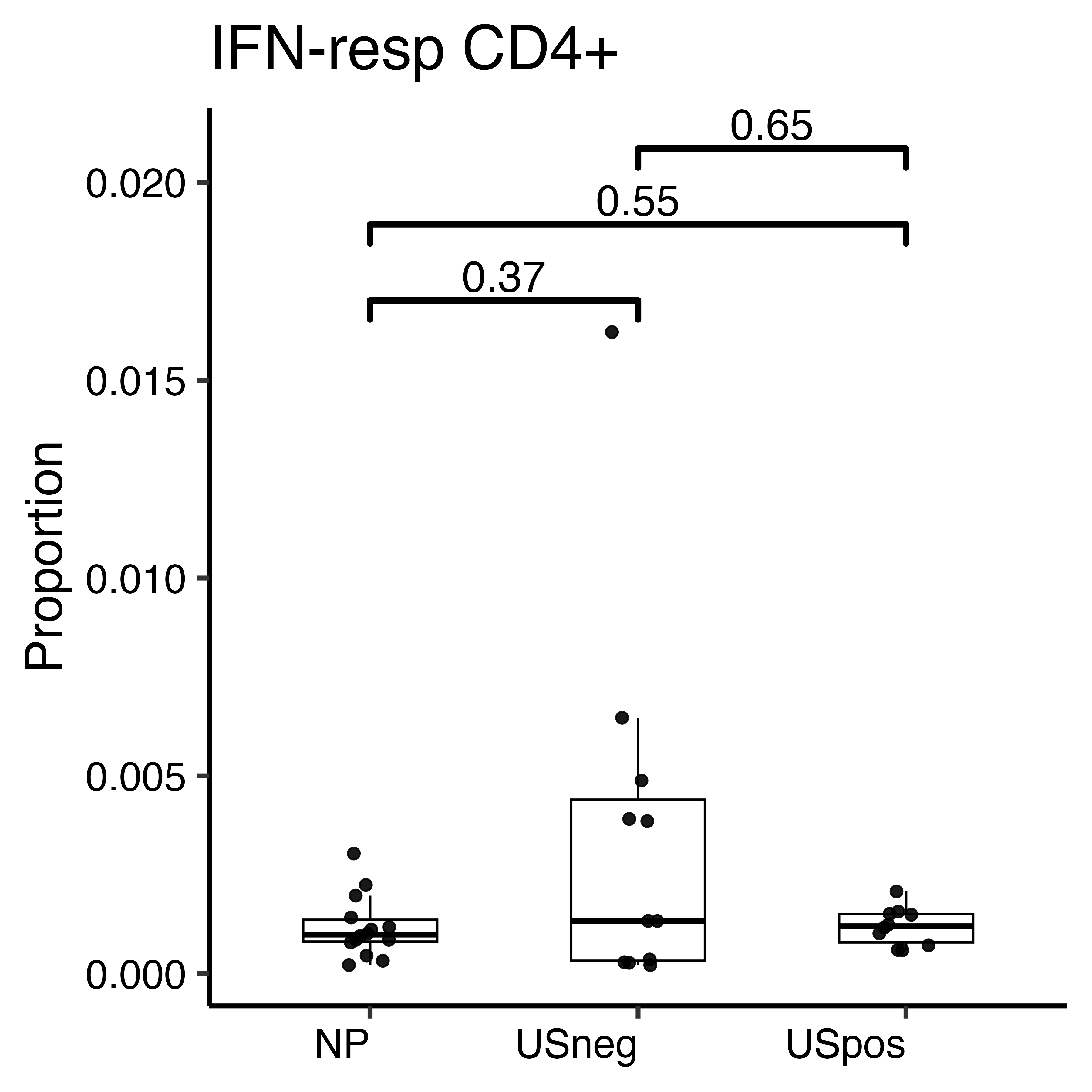

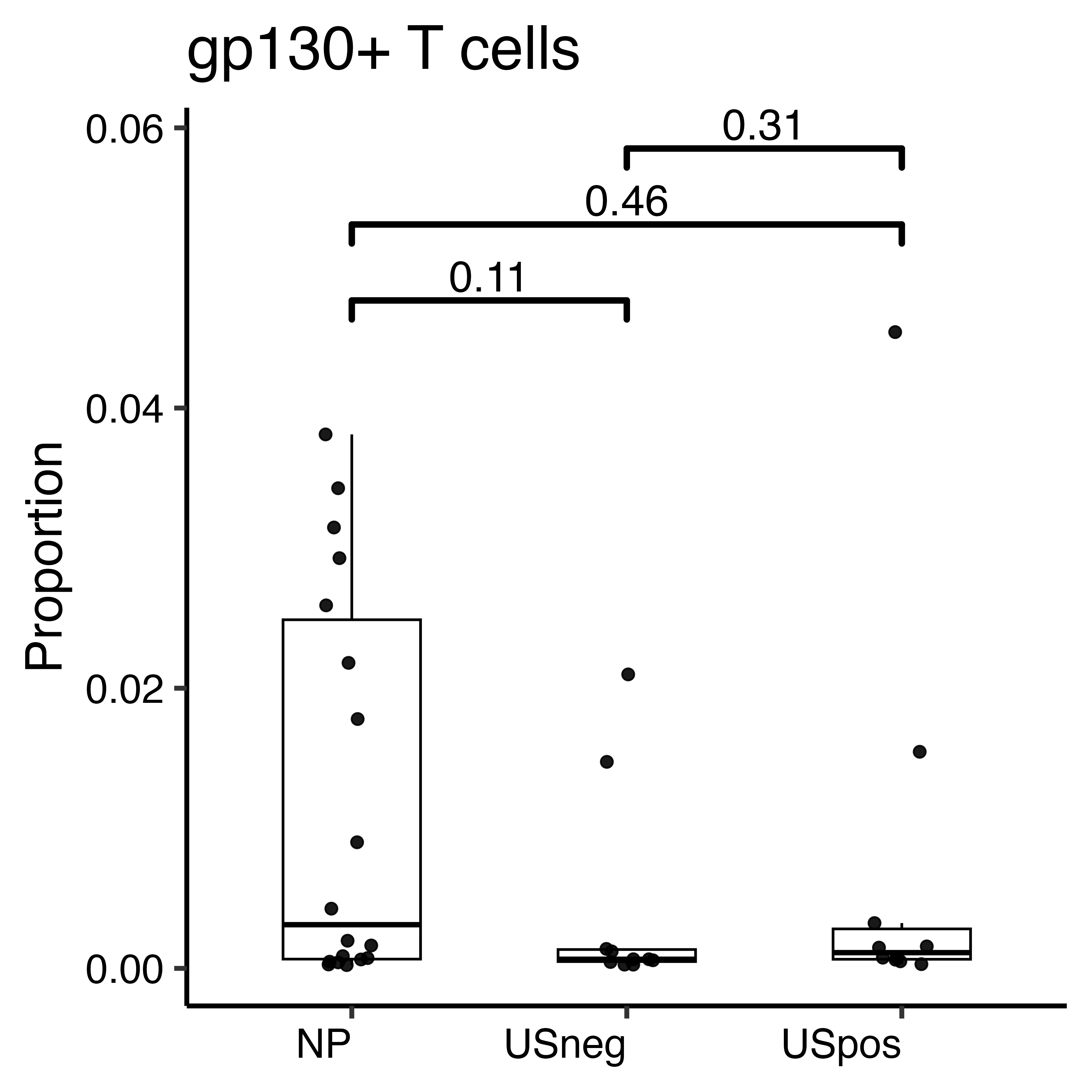

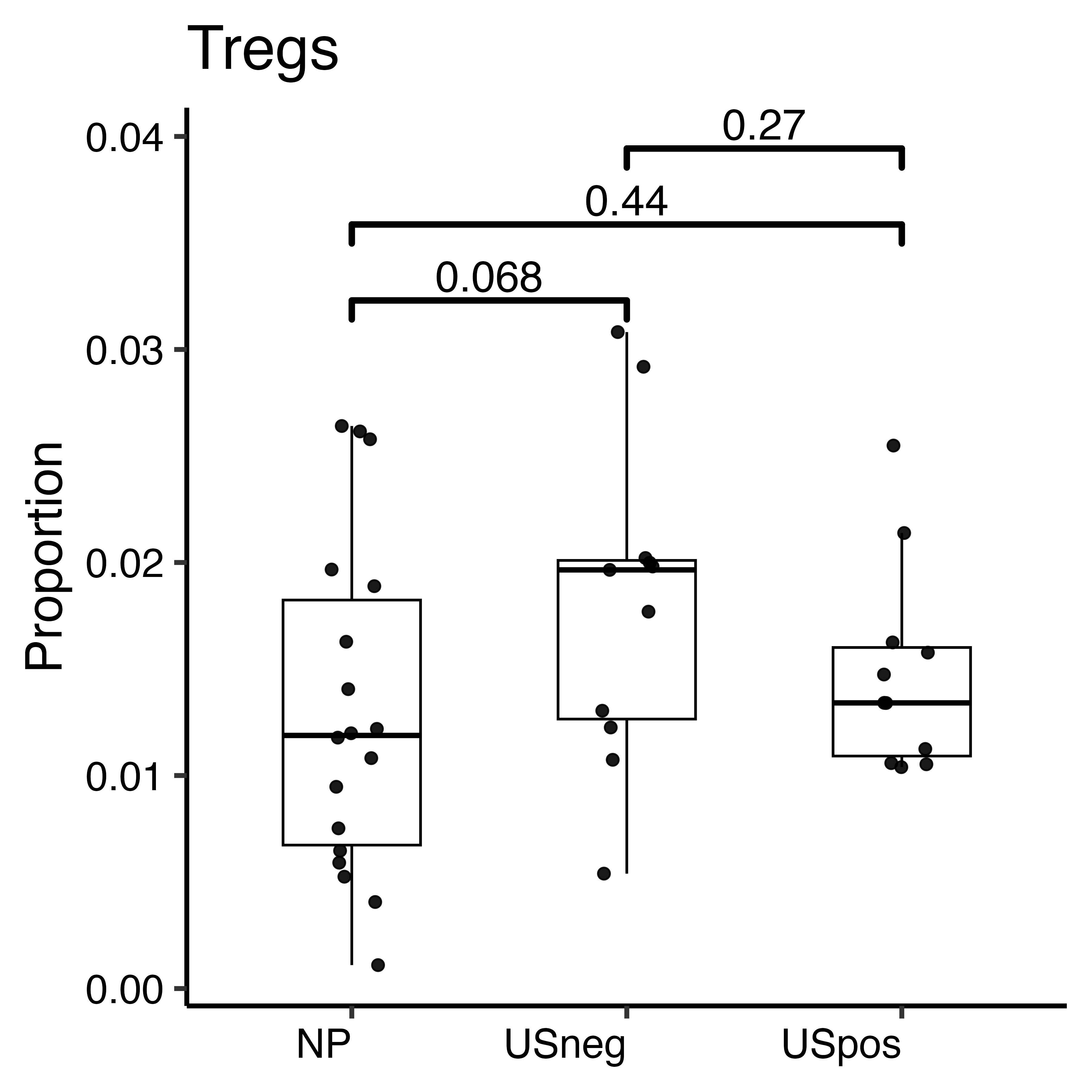

b

CD8+ T cells

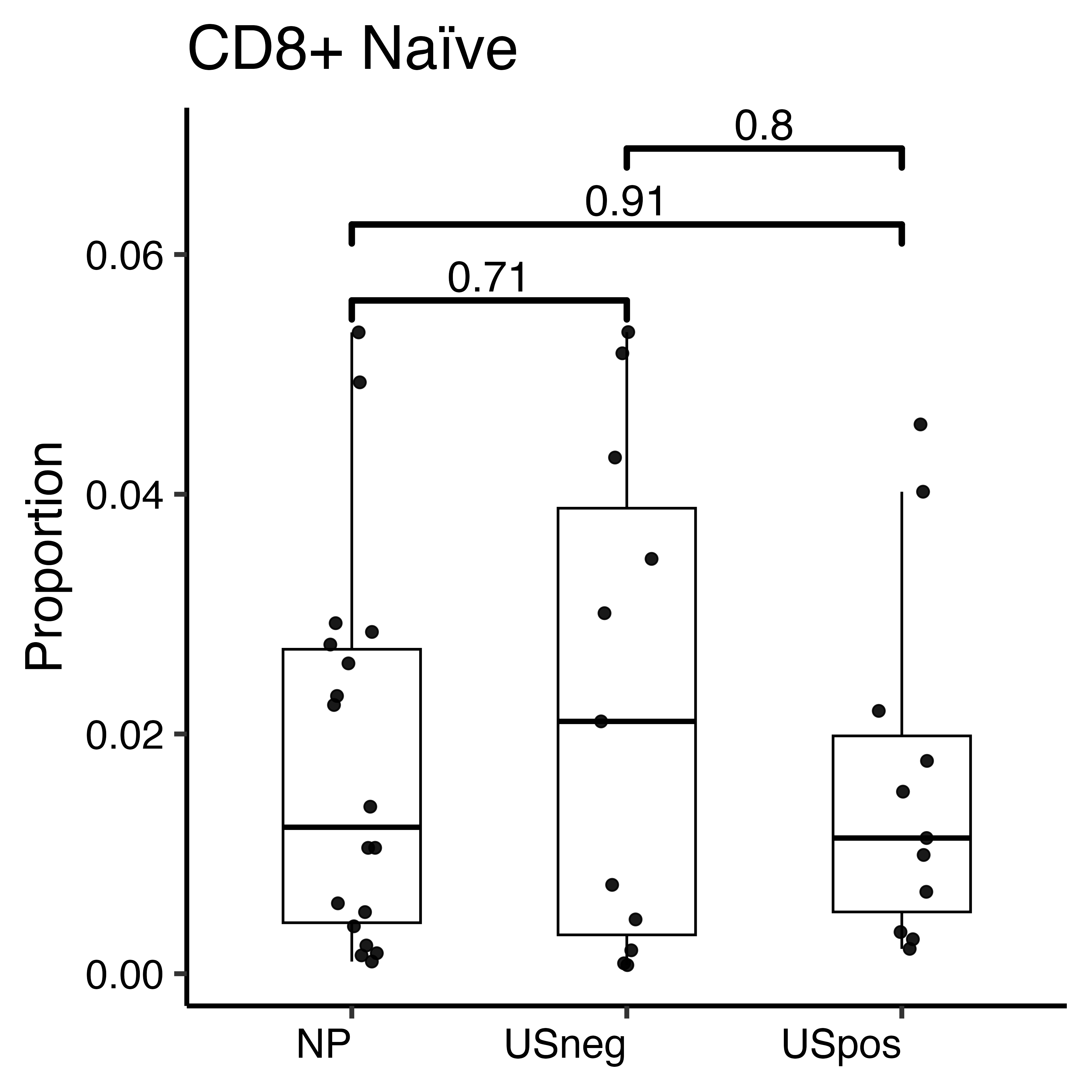

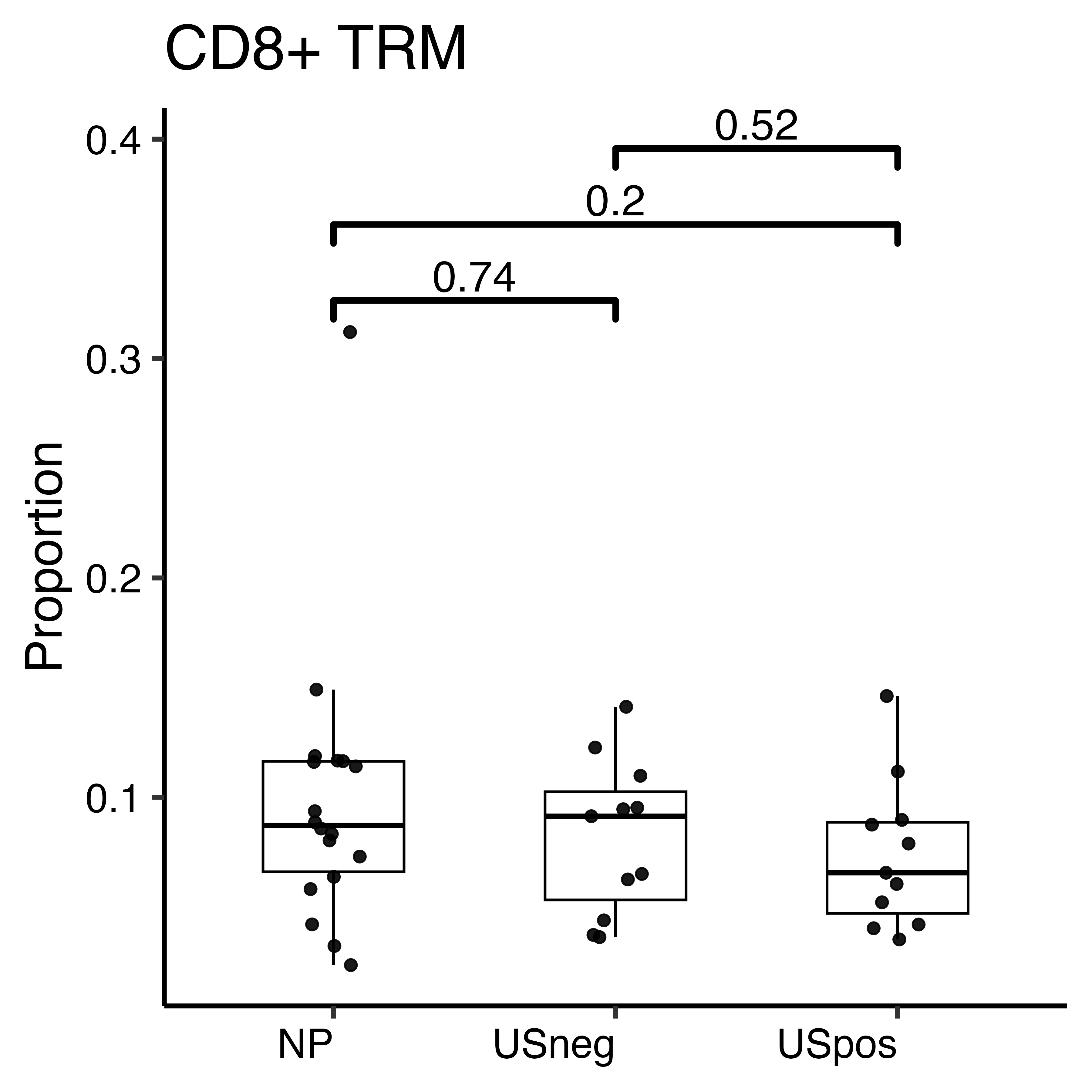

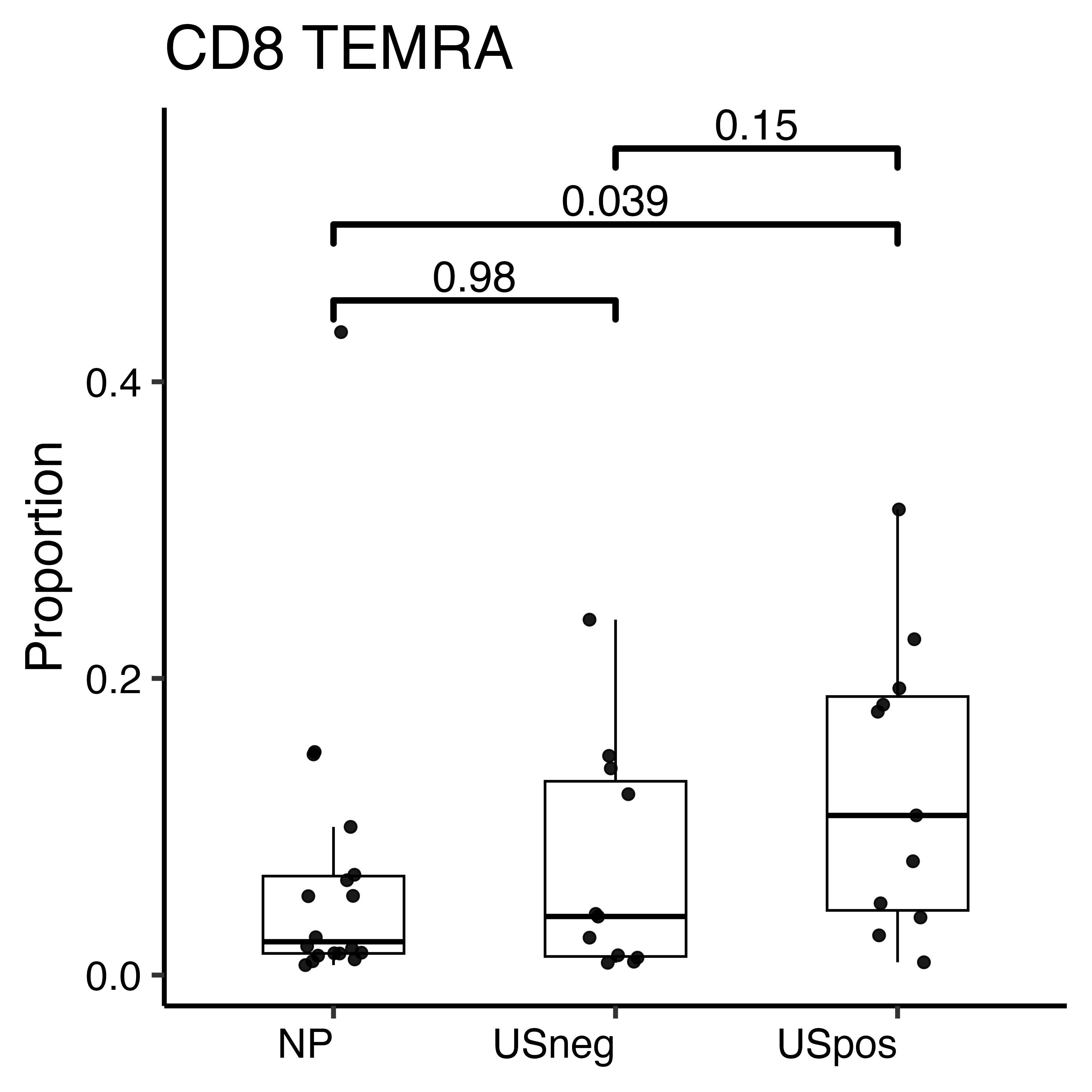

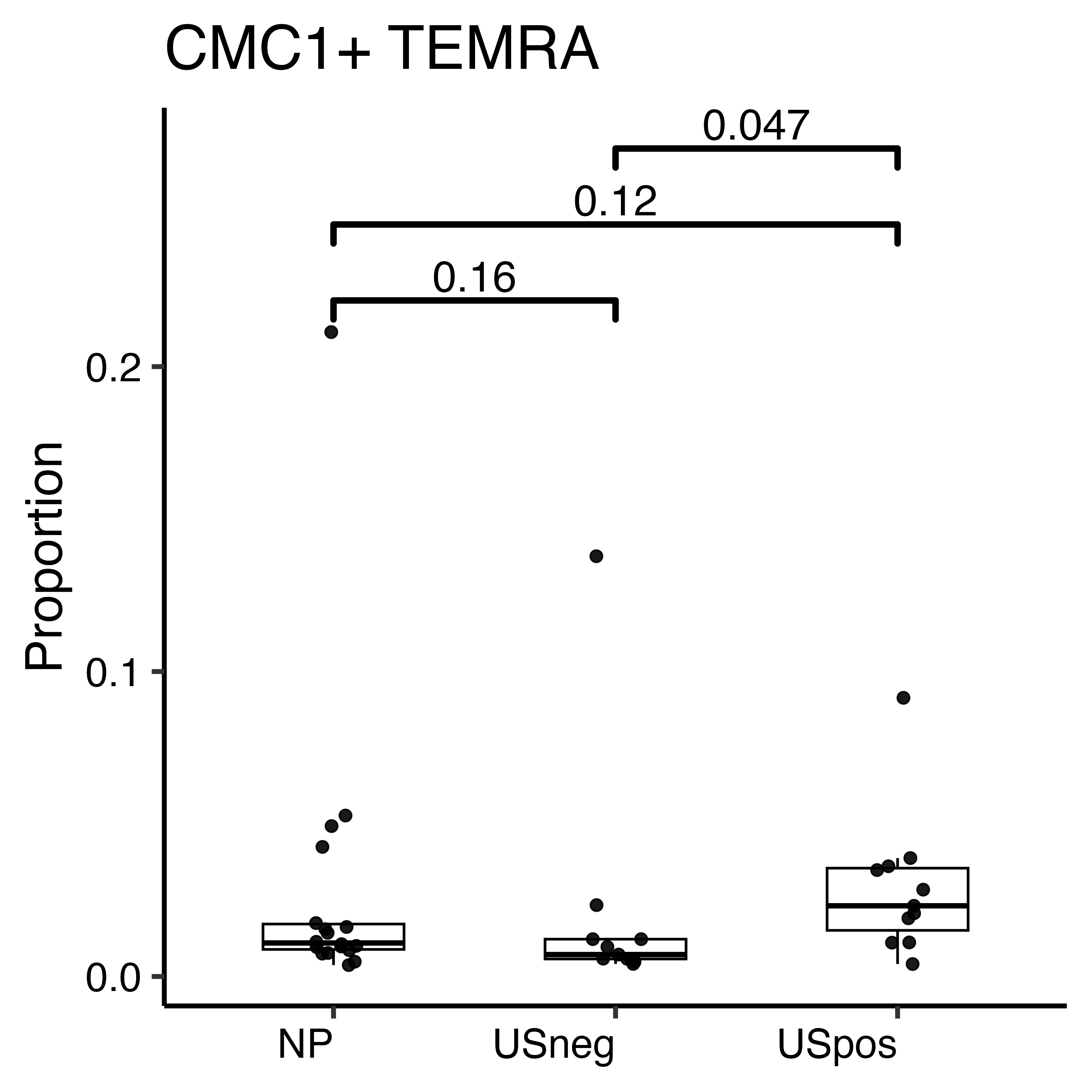

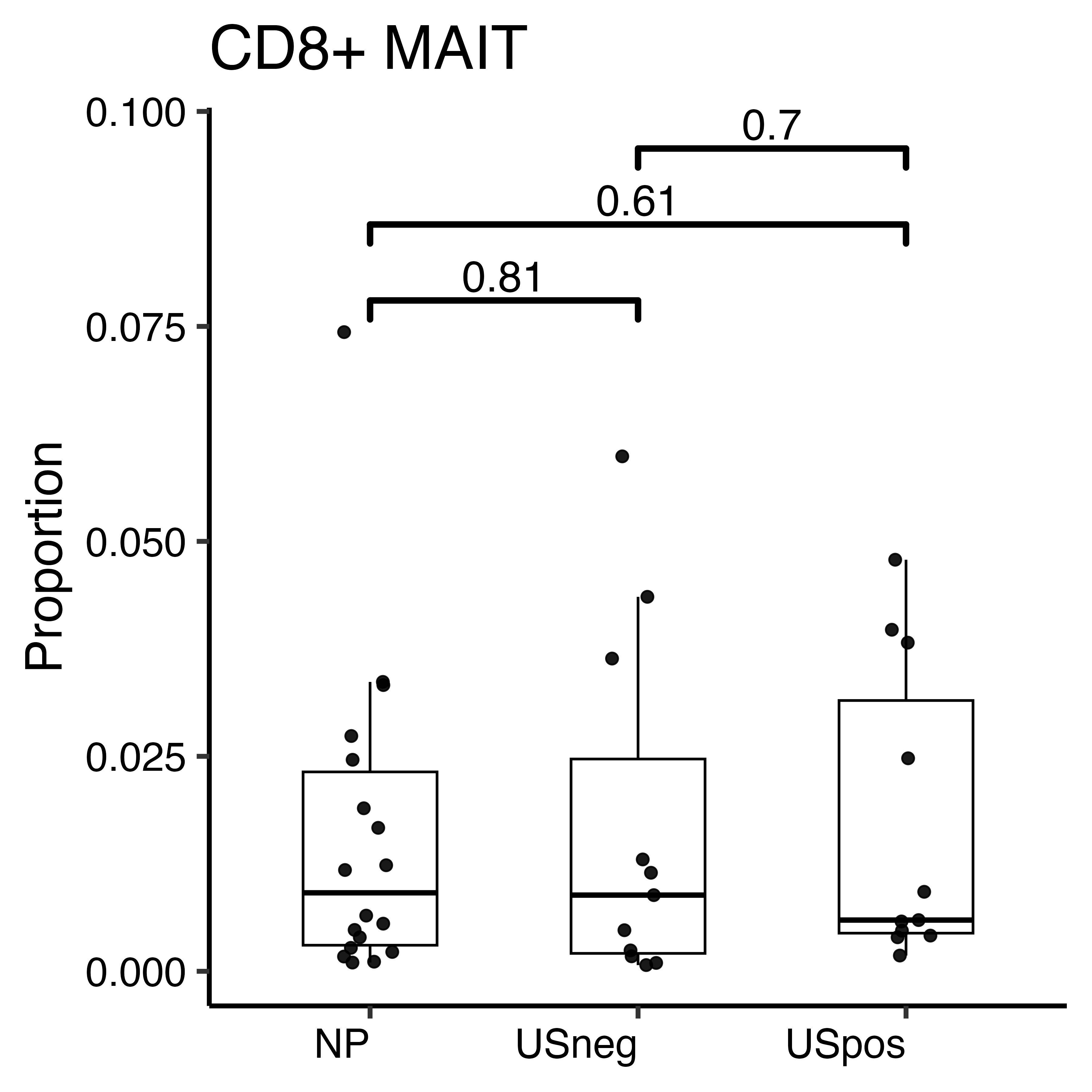

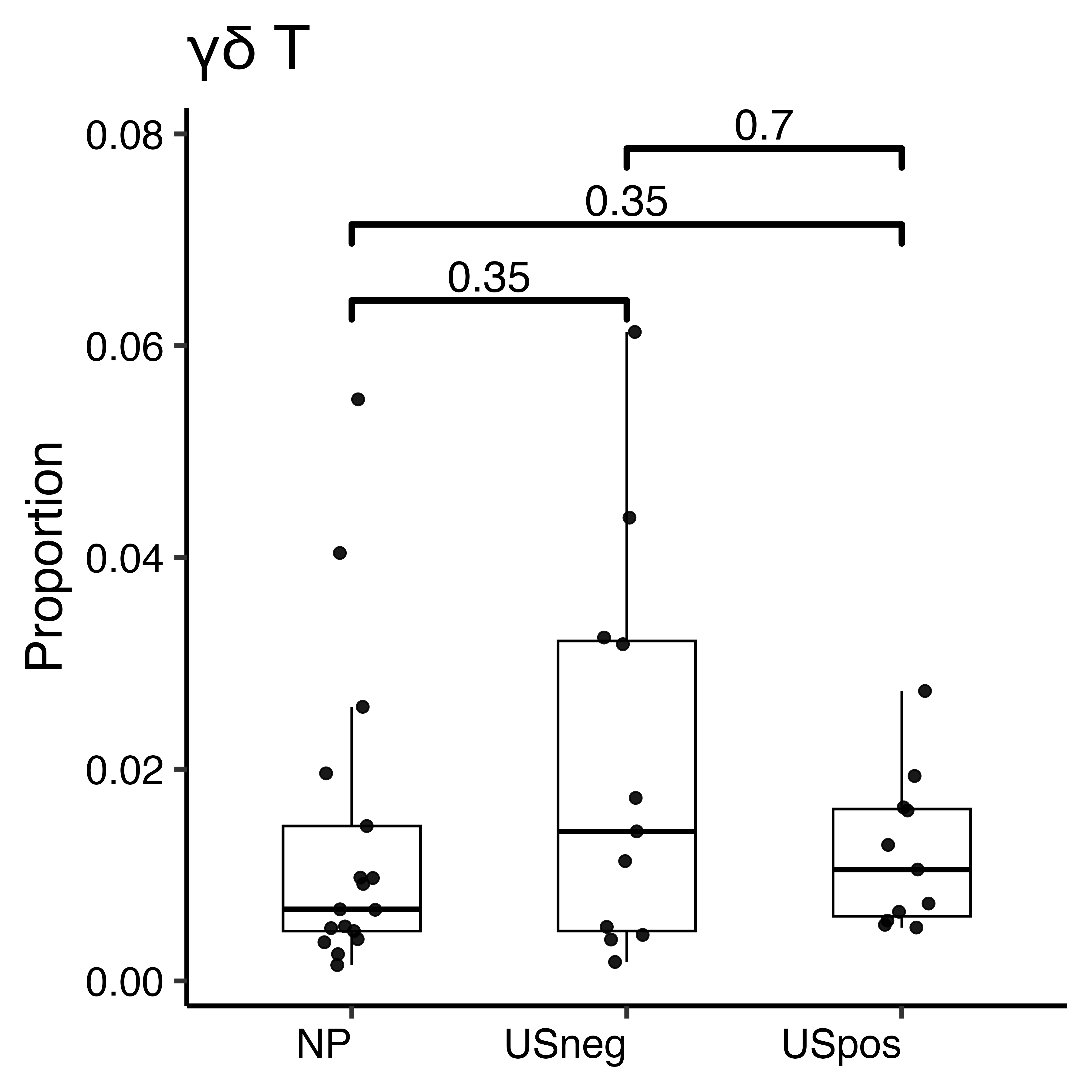

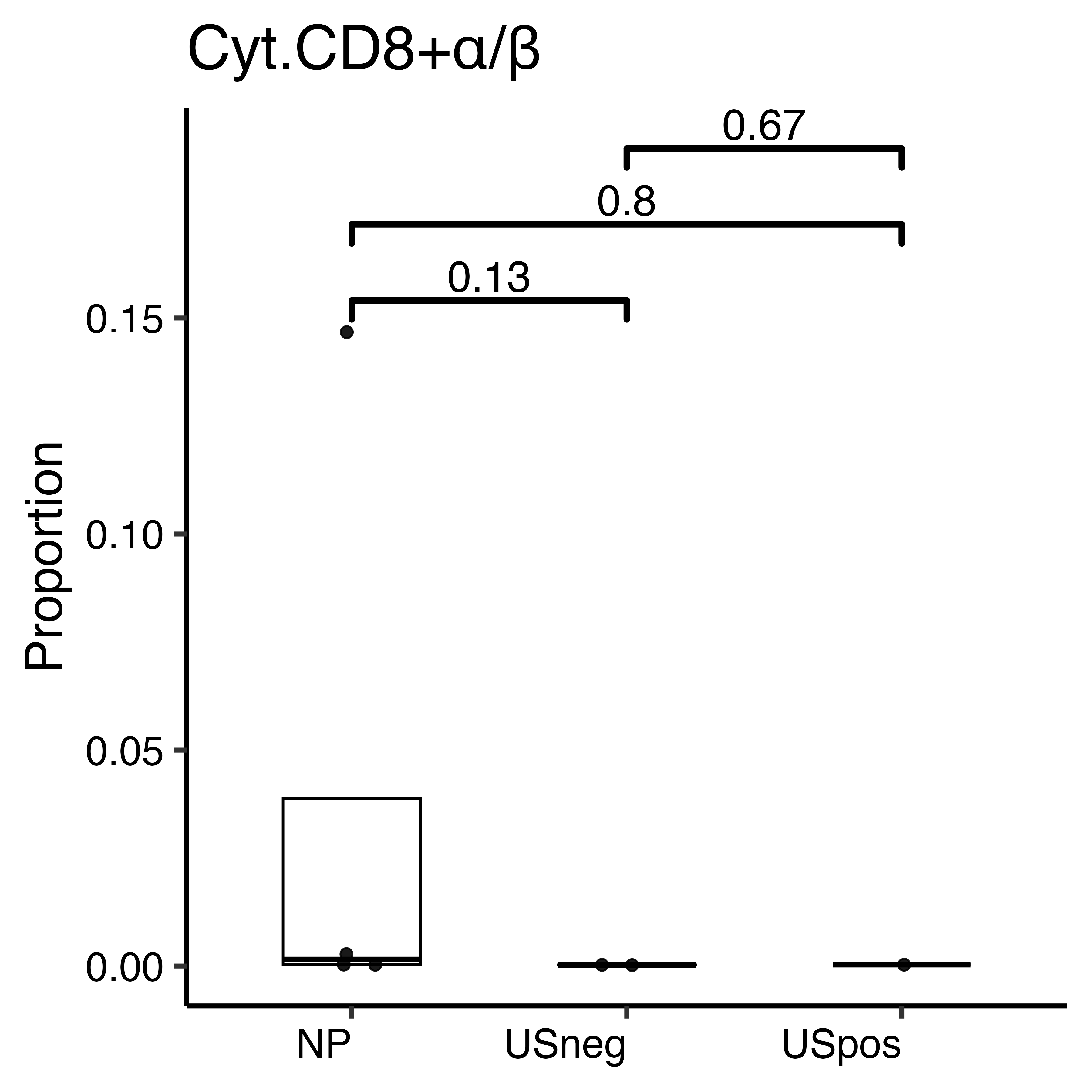

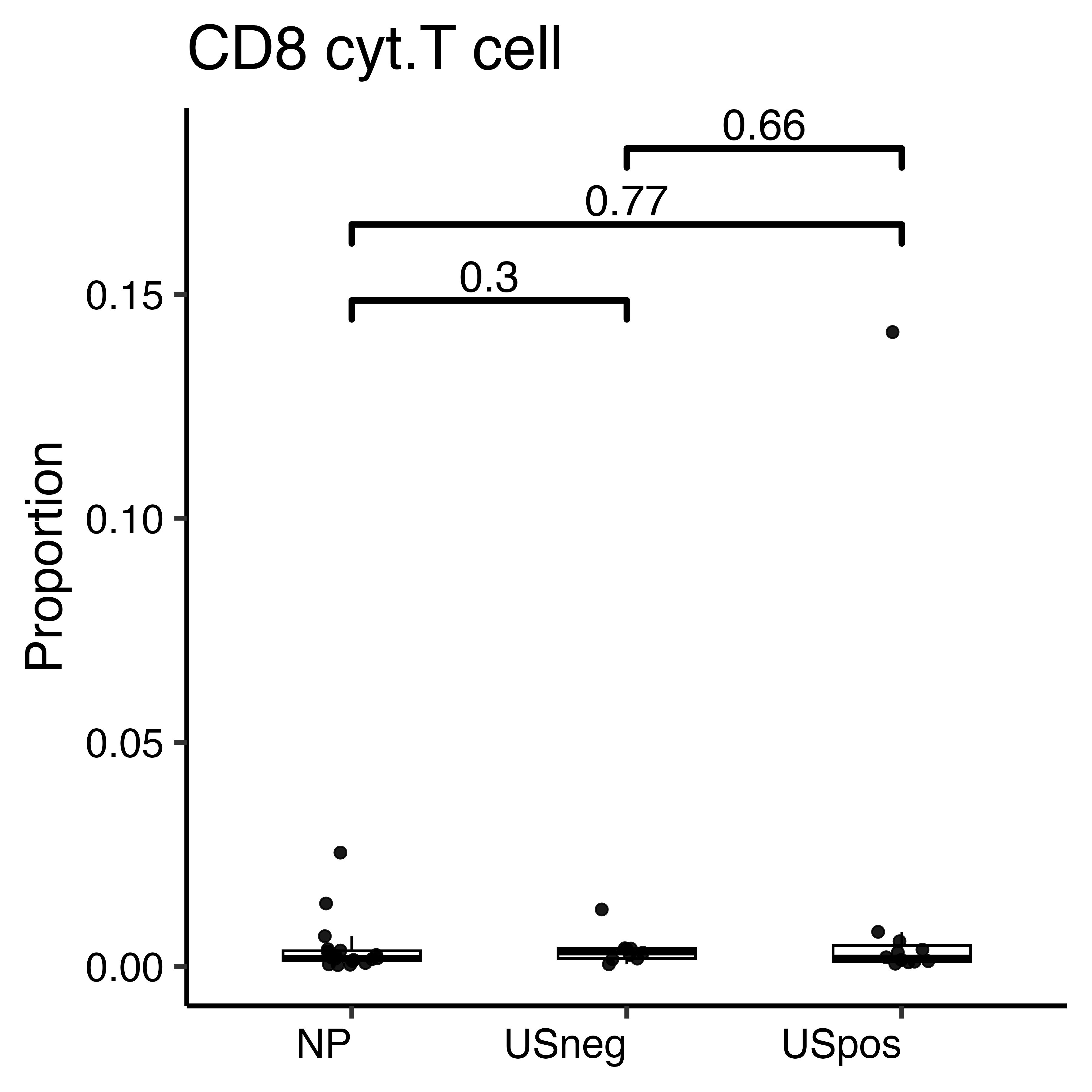

c

NK cells

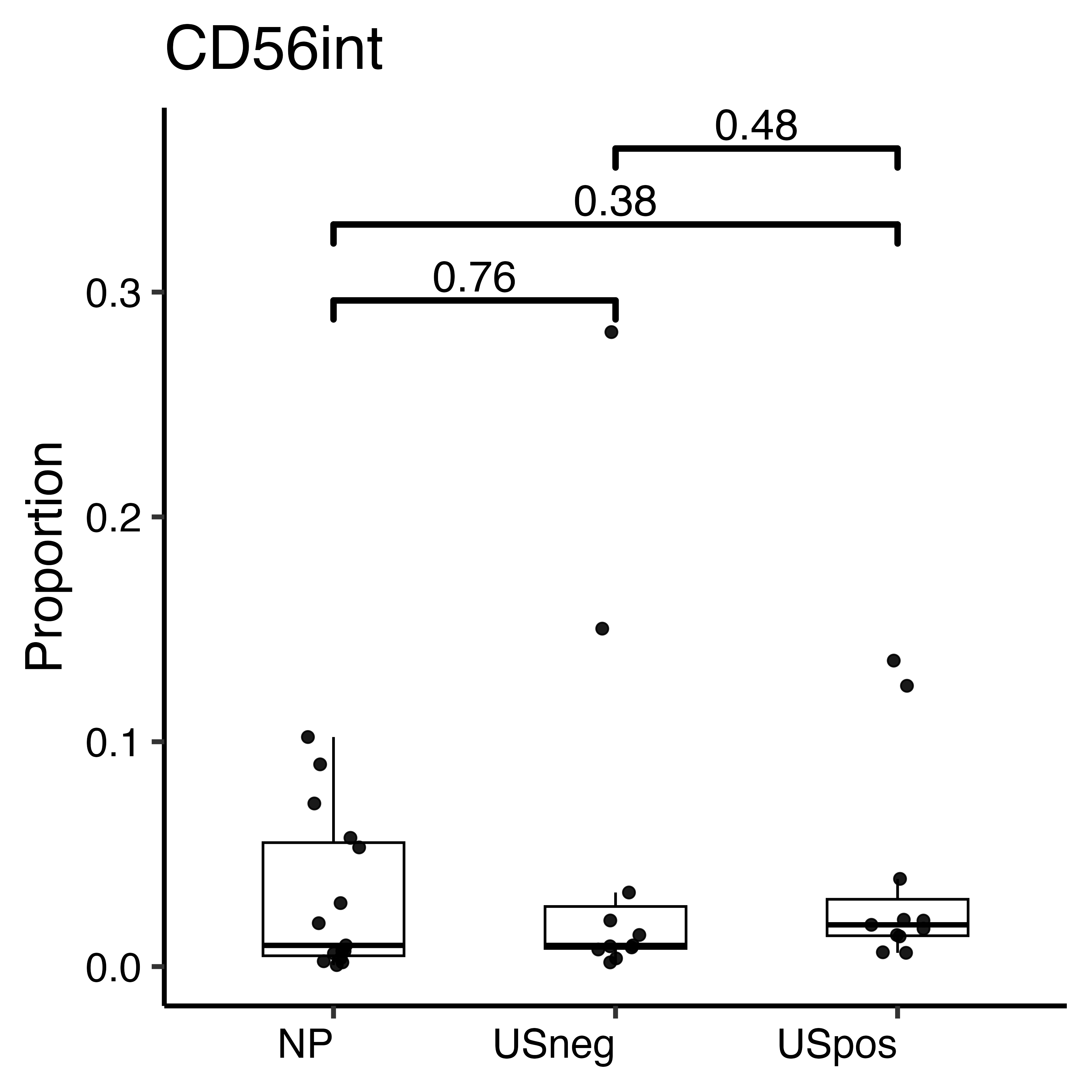

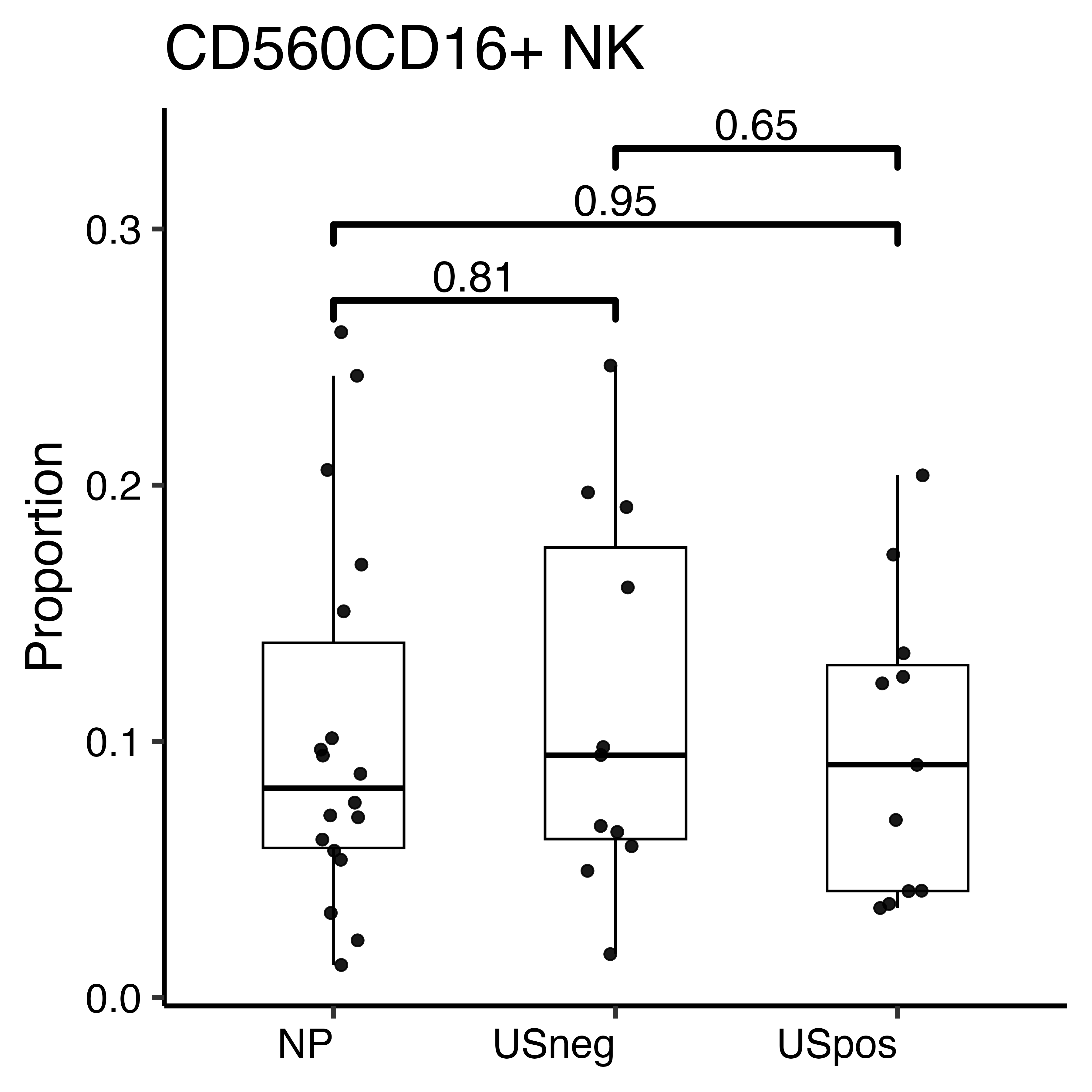

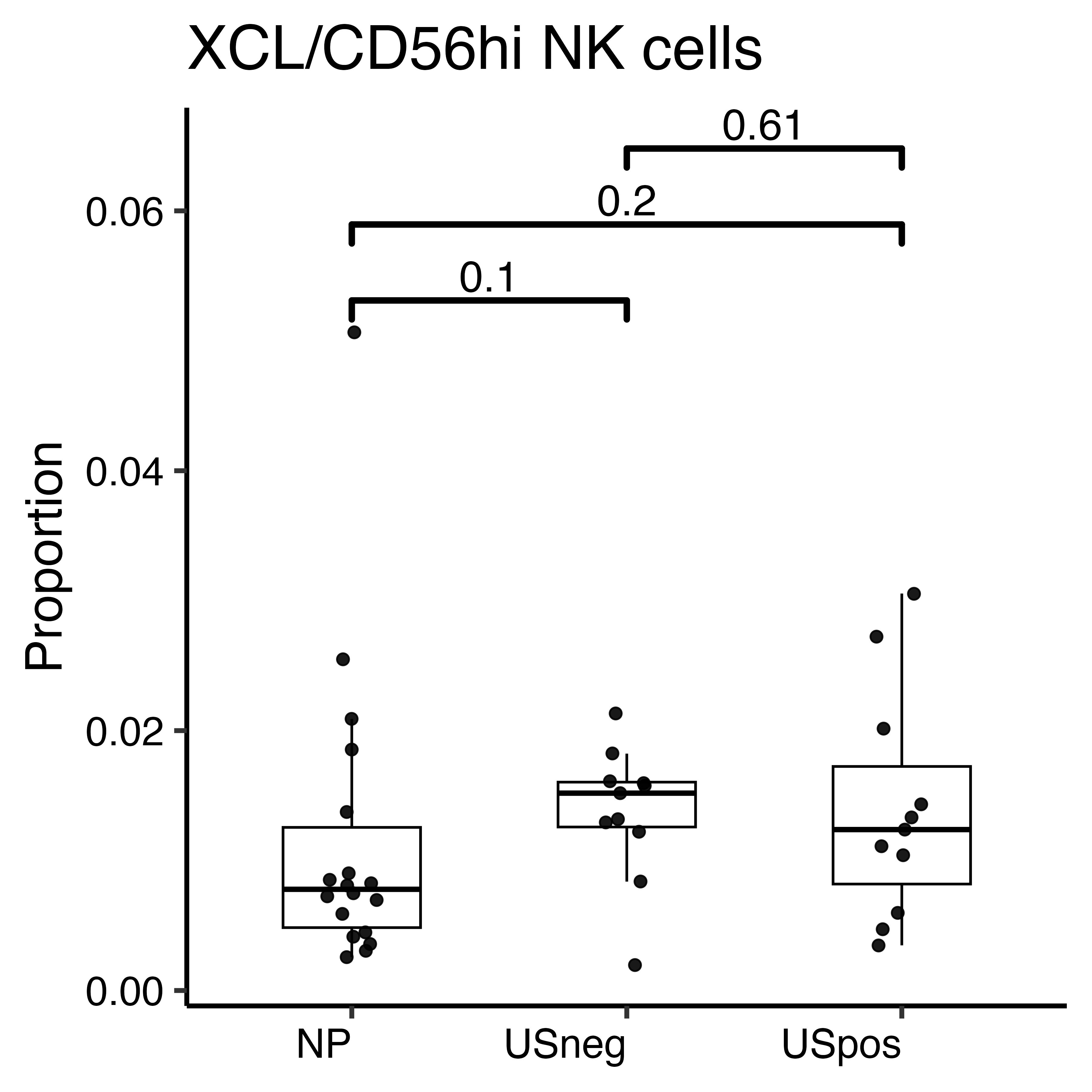

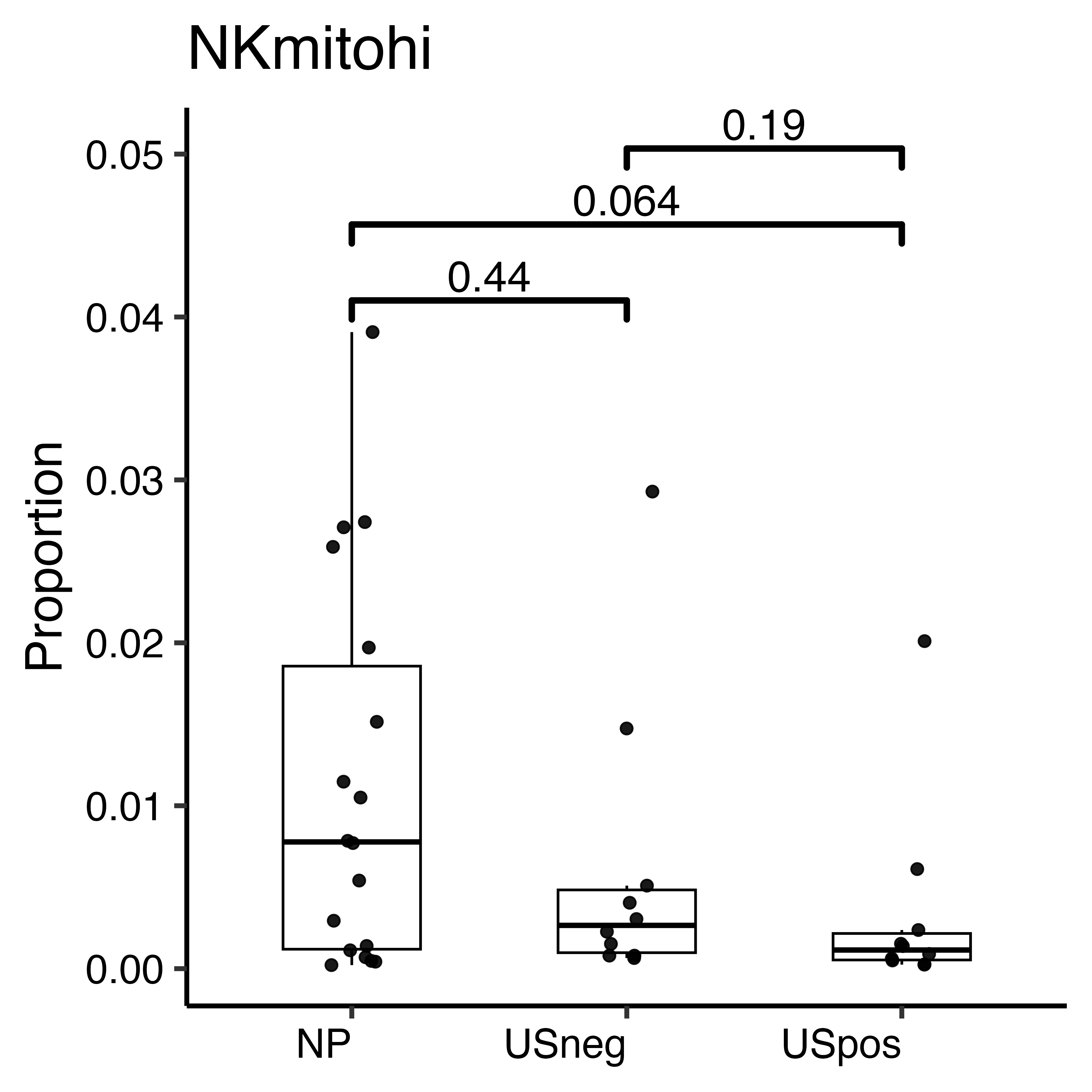

d

Monocytes

IFNresMono

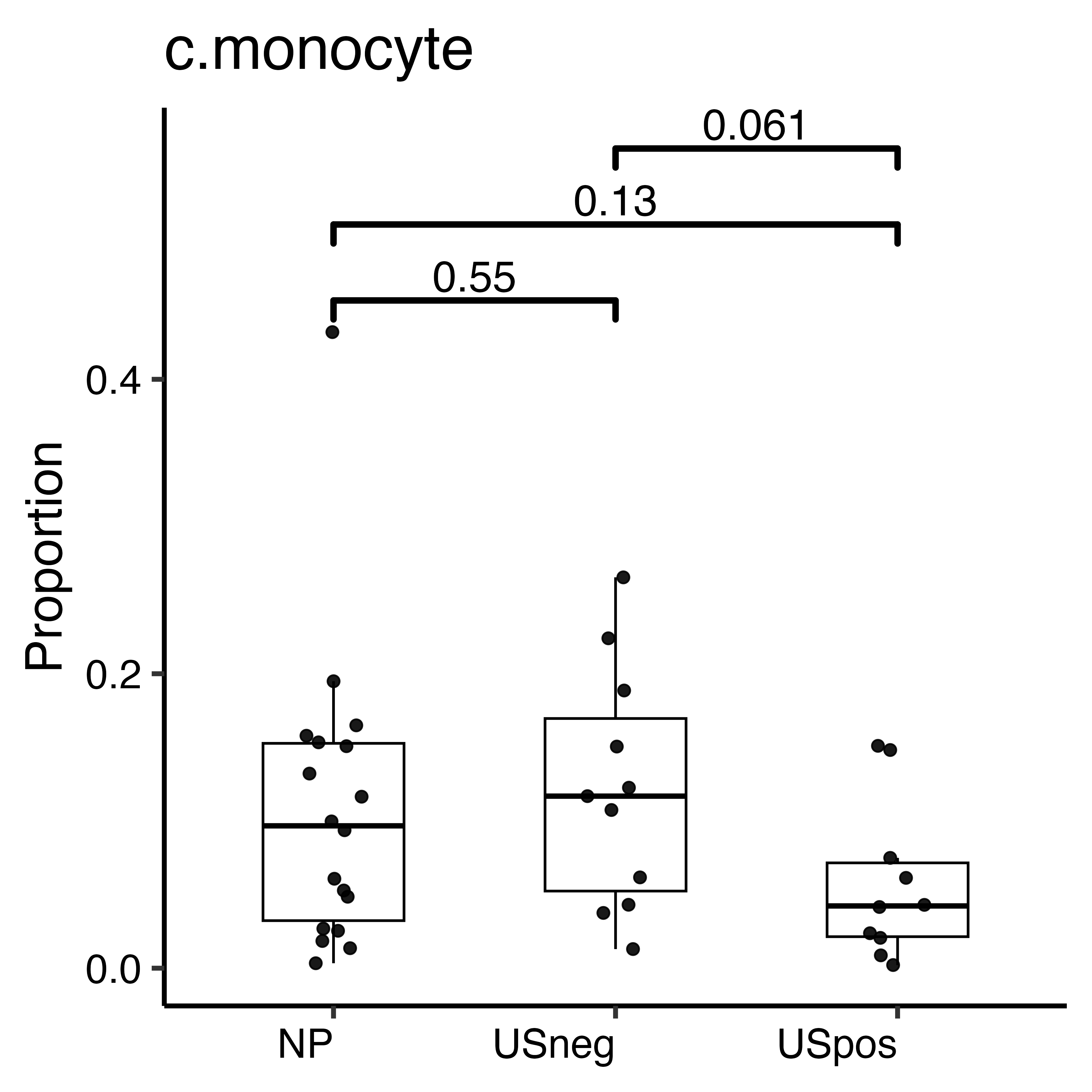

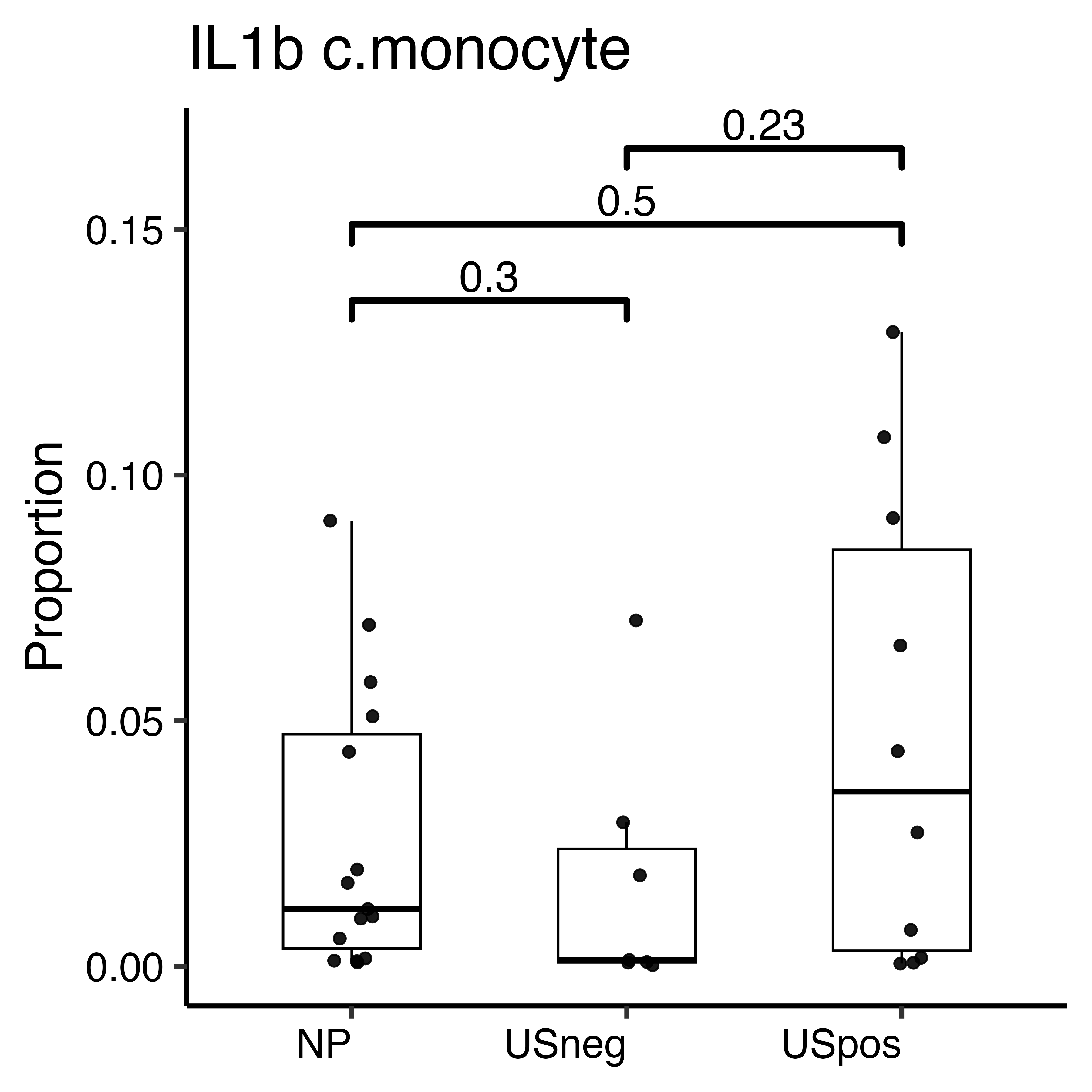

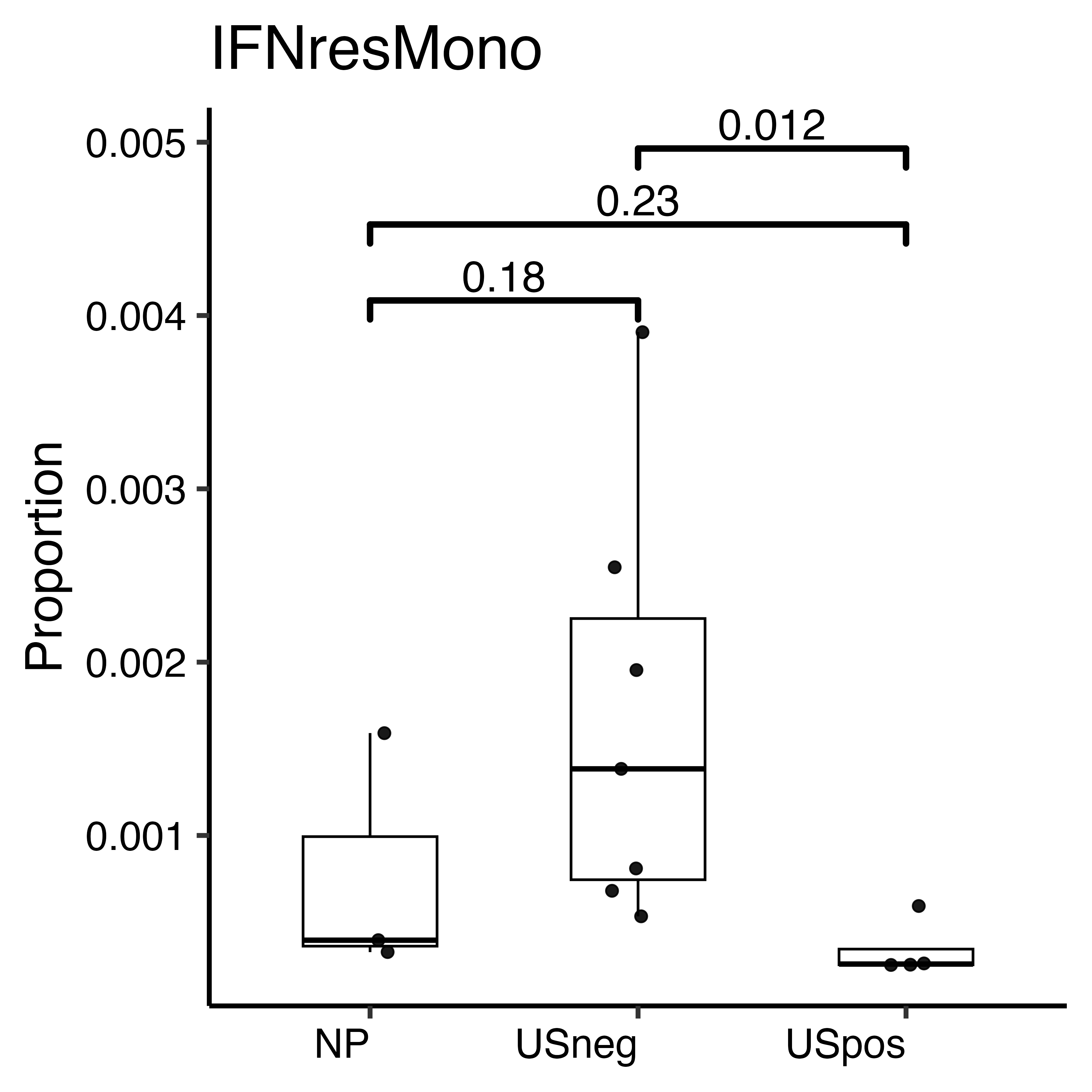

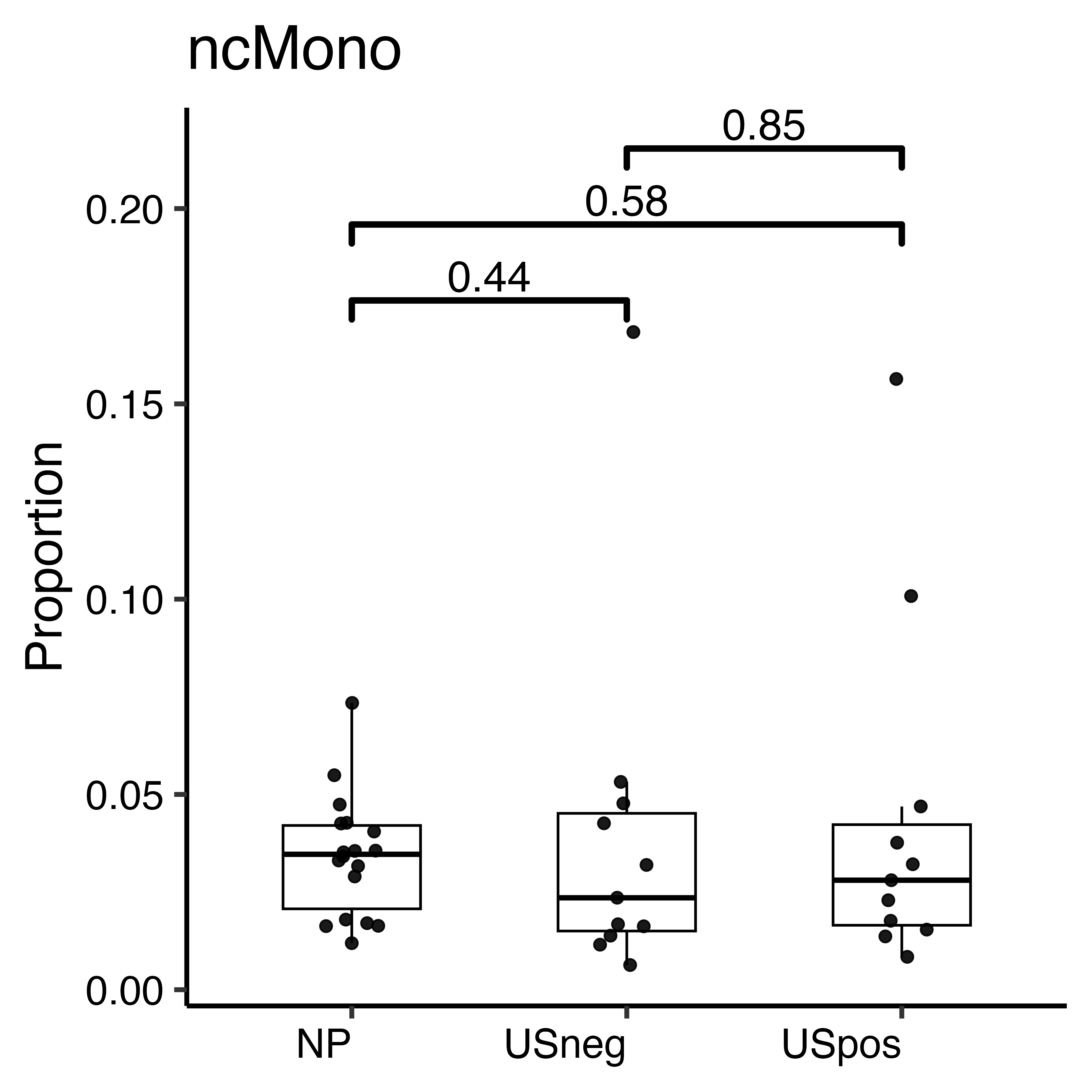

Other subsets

e

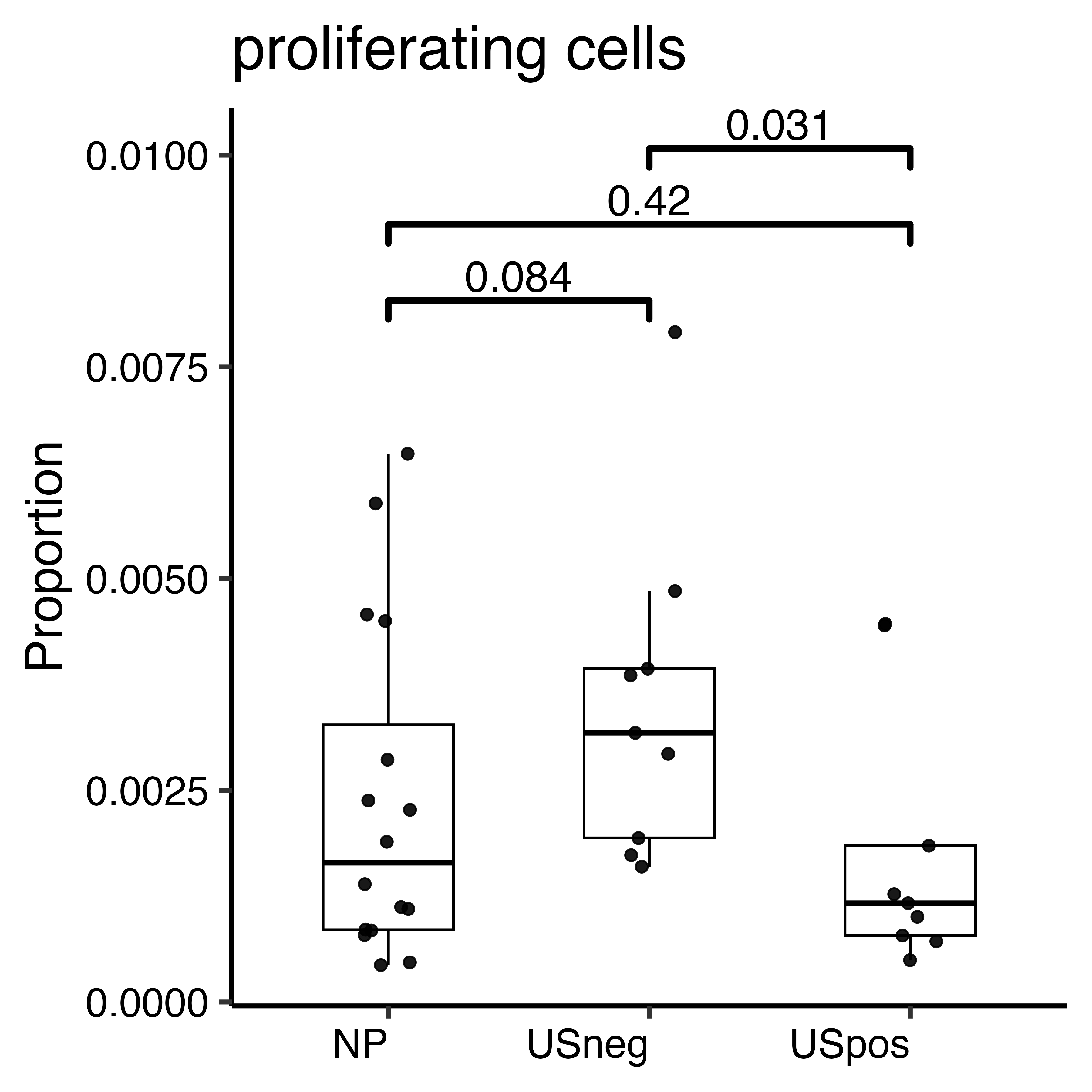

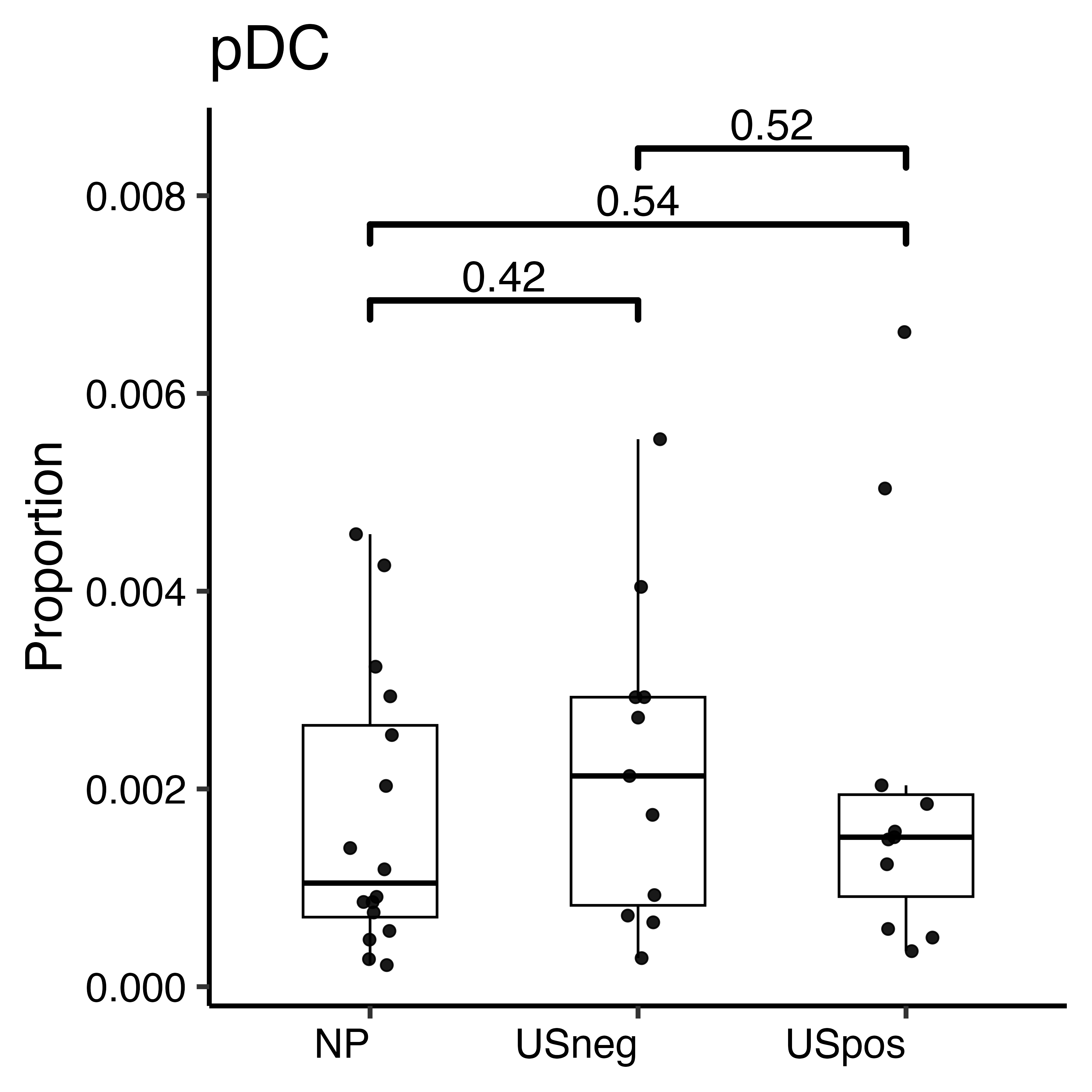

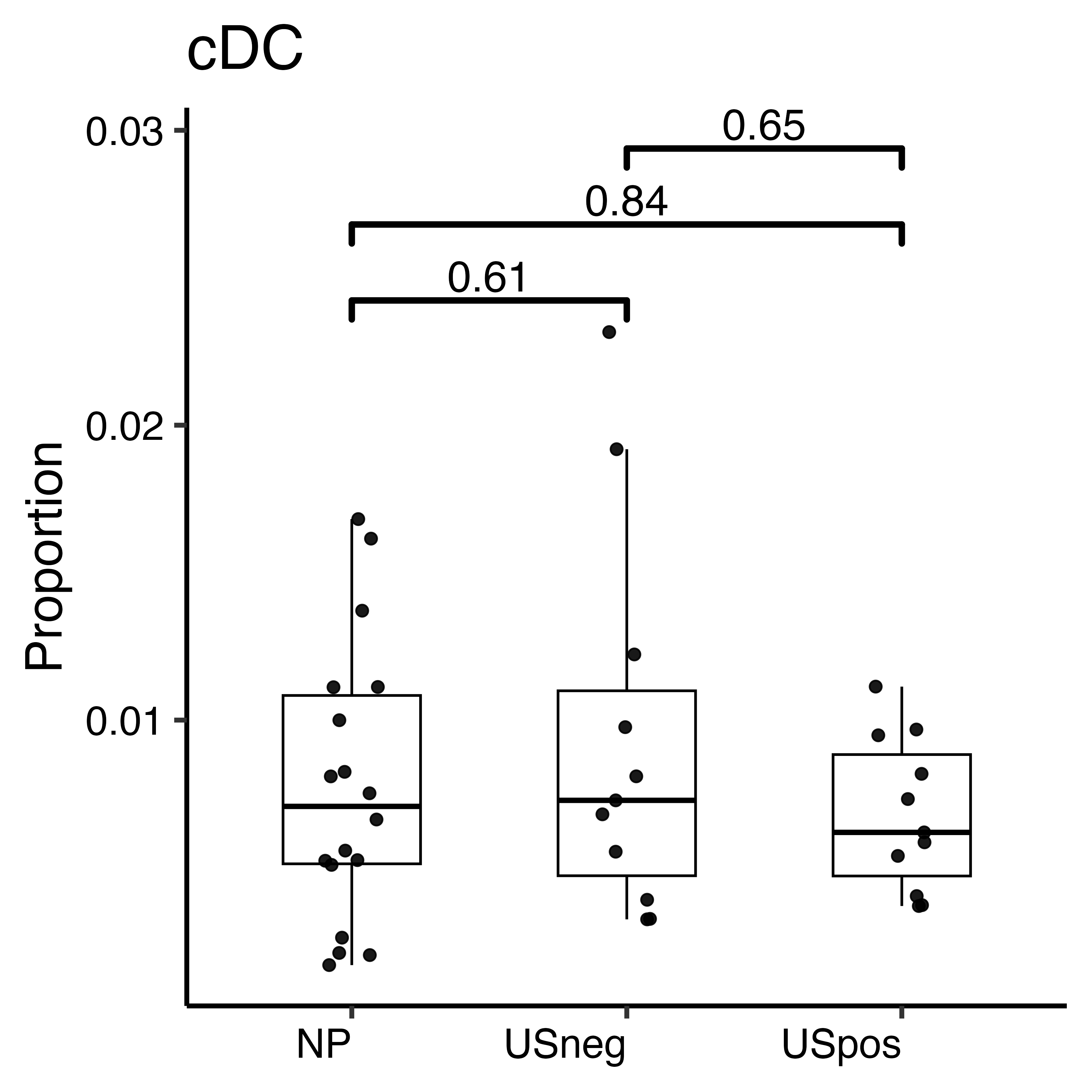

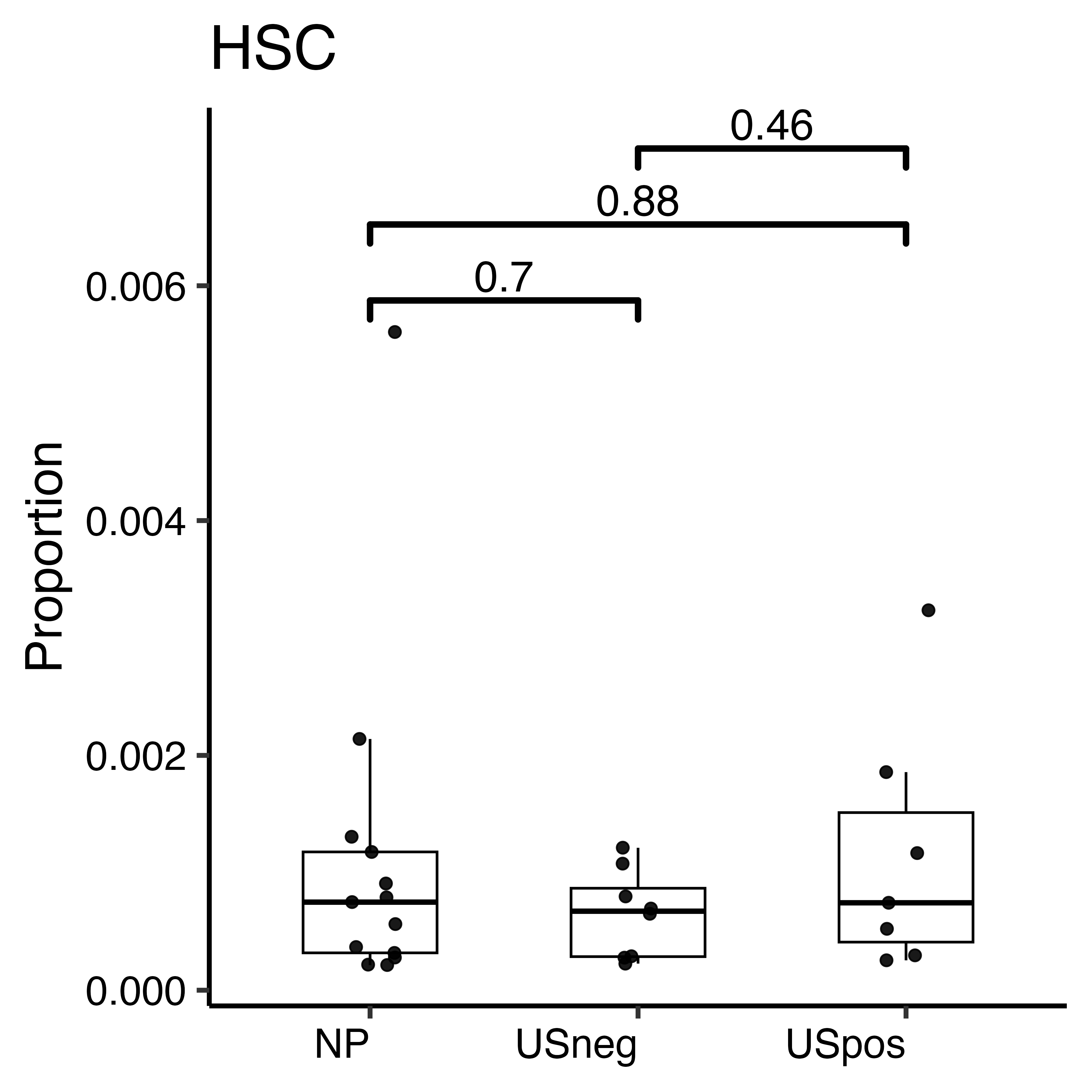

B cells

f

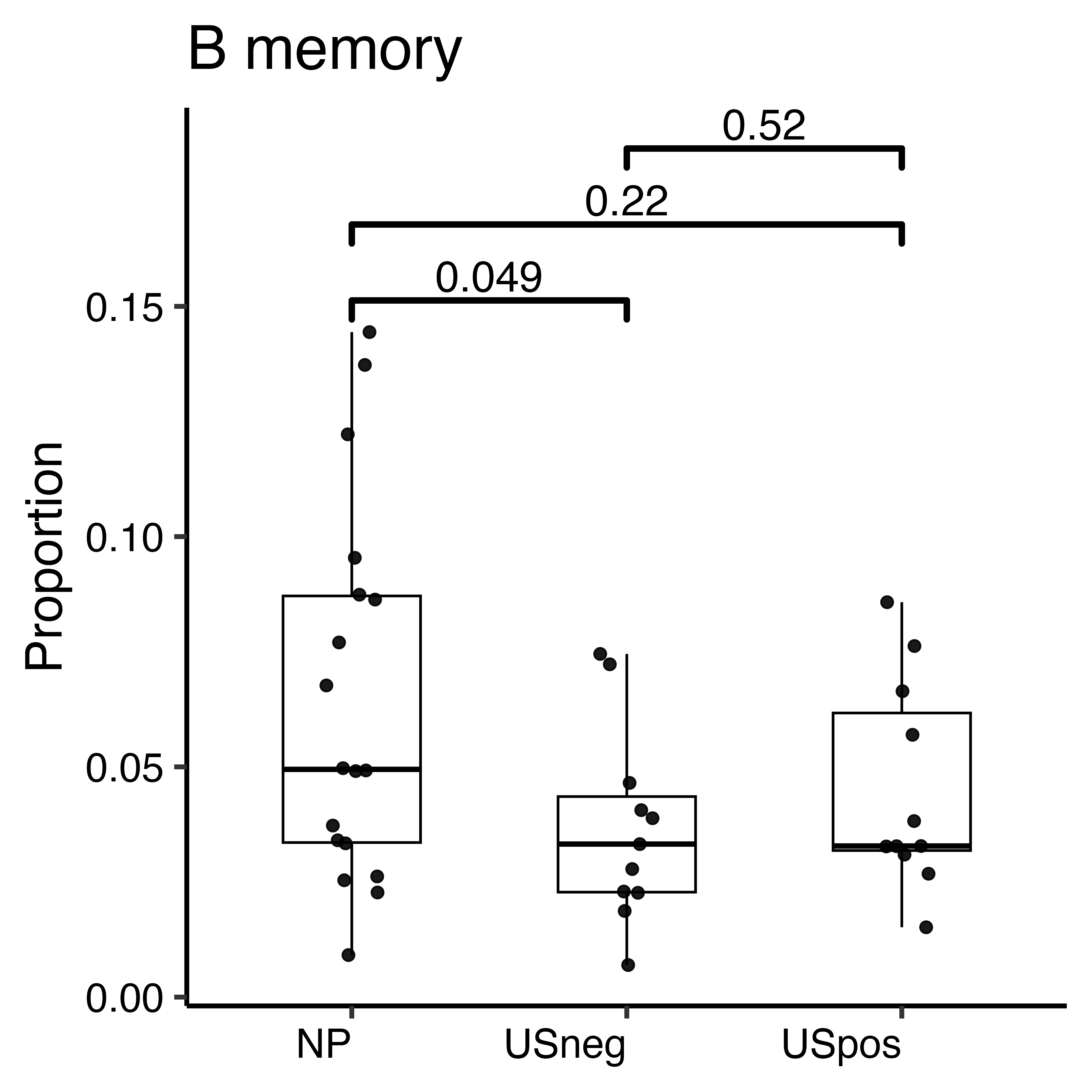

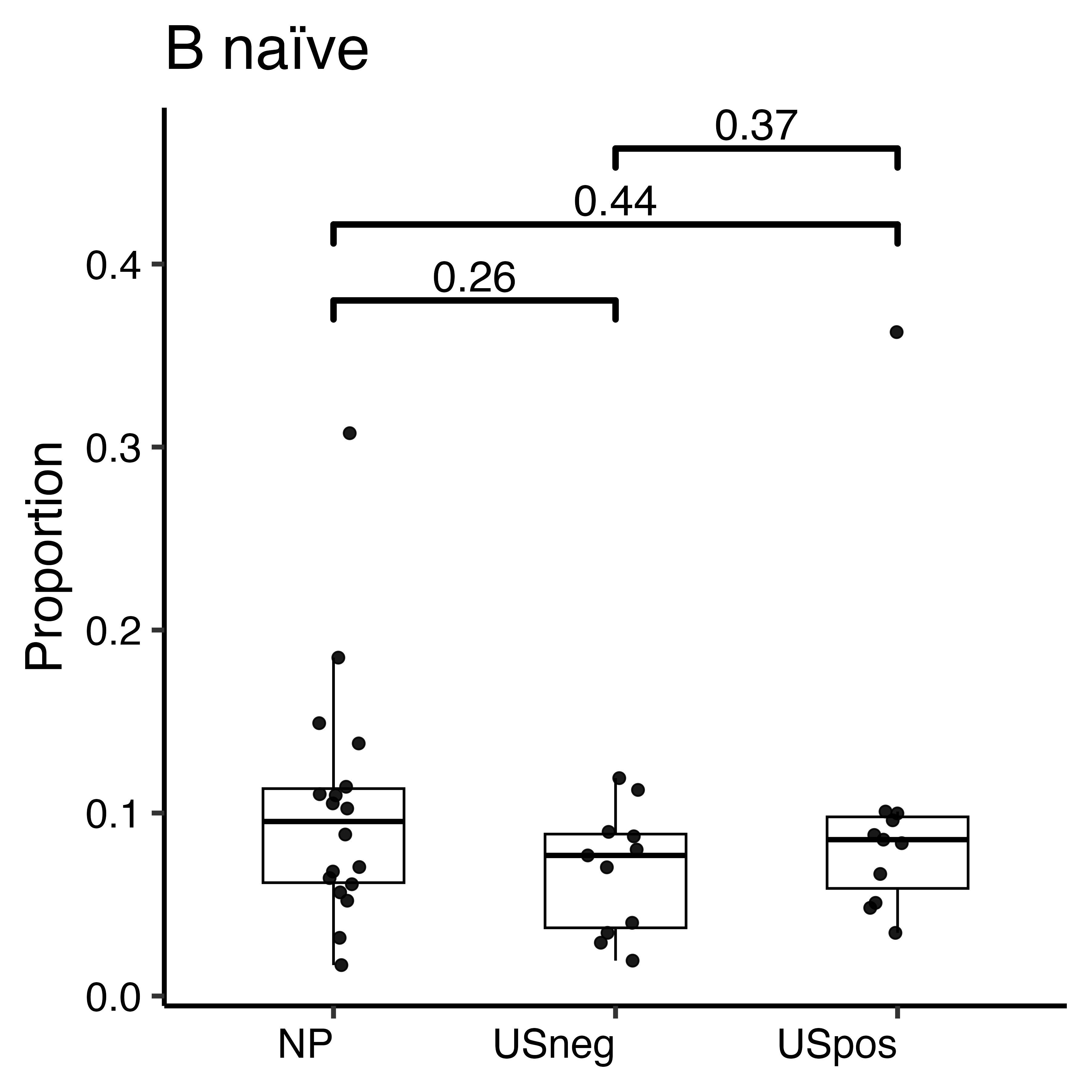

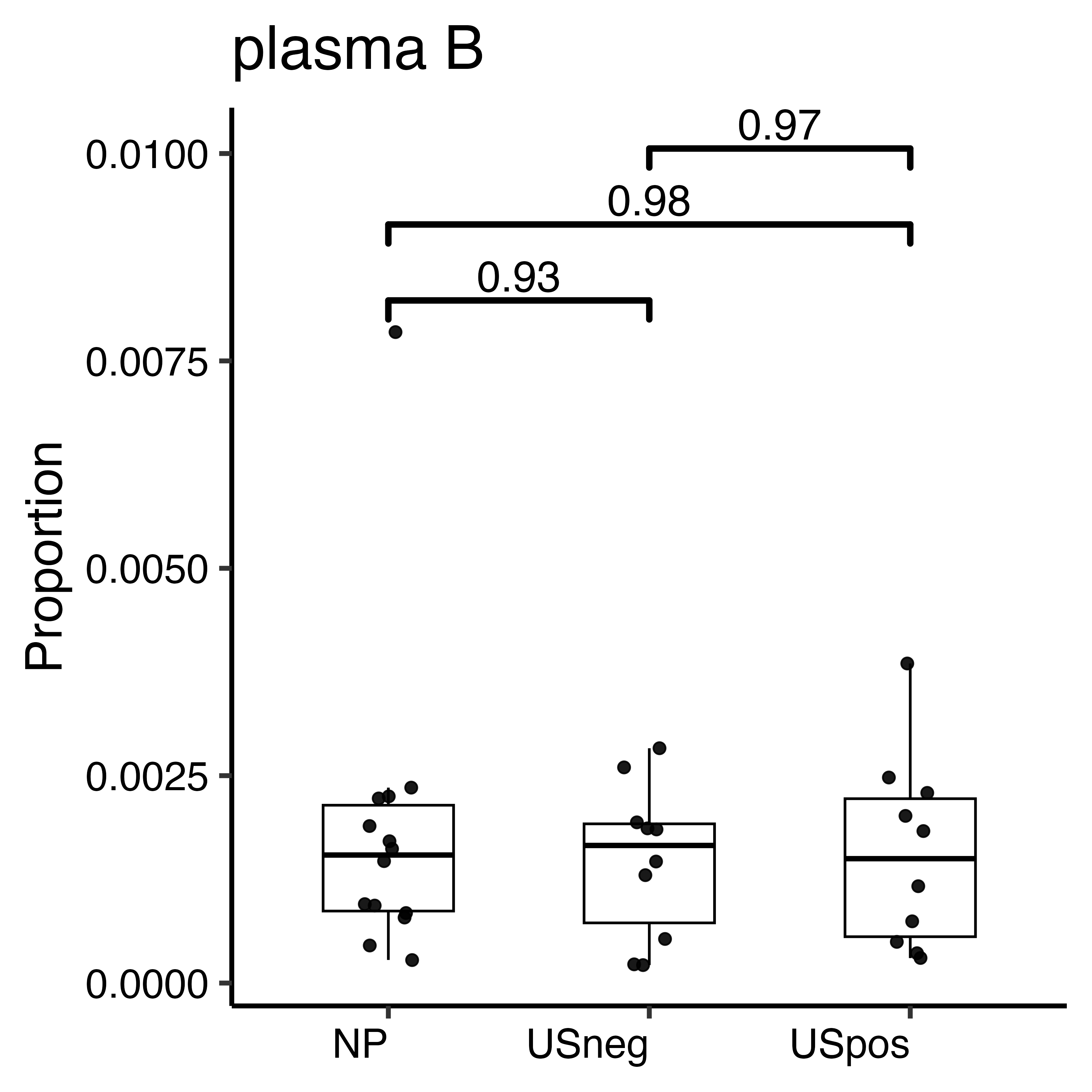

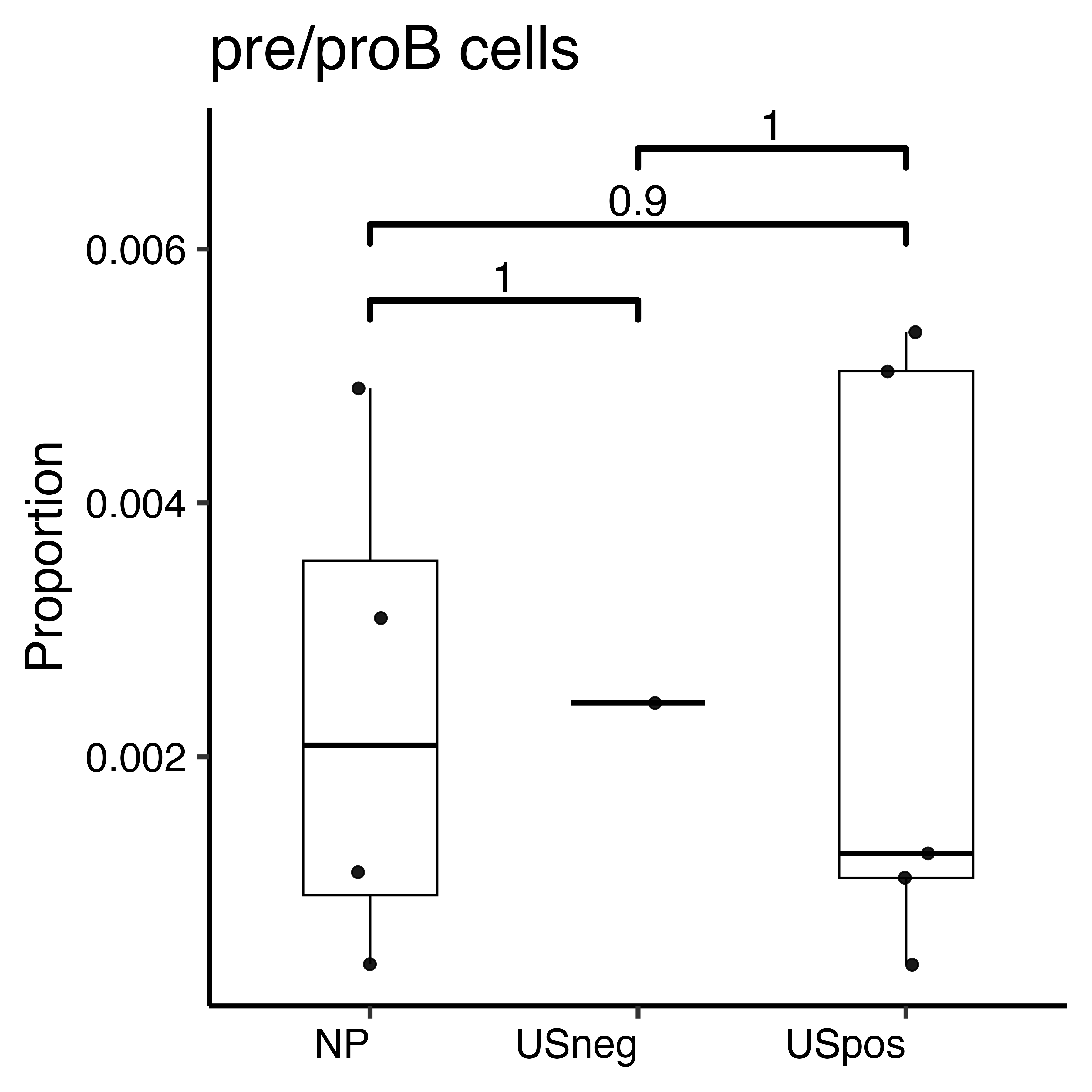

### **Figure S4. Proportional differences in immune cell subsets among ACPA+ at-risk RA groups.** Boxplots showing the relative frequencies of immune cell subsets (a- CD4+ T cells, b- CD8+ T cells, c- NK cells, d- monocytes, e- other subsets including pDCs, cDC, proliferating cells, and HSC, f- B cells) among individuals from three at-risk RA groups: NP, USneg, and USpos progressors. Each boxplot represents interquartile range (IQR) with horizontal line indicating median. Pairwise group comparisons were performed using the two-sided Wilcoxon rank-sum test in R (v4.4.1). Reported p values represent unadjusted significance levels between the indicated groups (p < 0.05, p< 0.01) and denoted above boxplots. Among monocytes (d), significant differences were observed in IFNresMono between USneg future progressors and USpos future progressors (p=0.01). In CD8+ T cells (b) CD8 TEMRA and CMC1 TEMRA were significantly elevated in USpos future progressors (p=0.039, and p=0.04, respectively). Among B cells, memory B cells (f) were significantly enriched in NP compared to USneg future progressors.

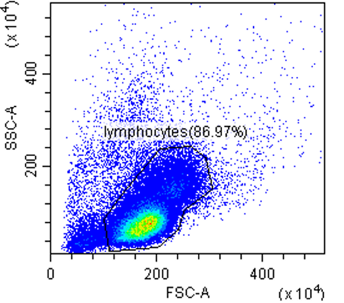

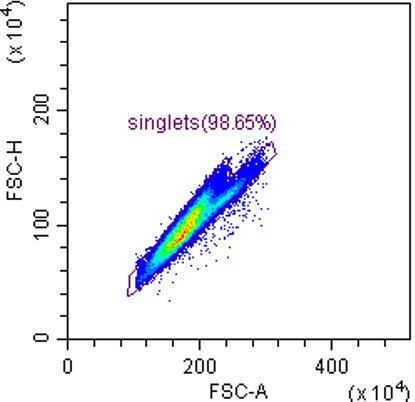

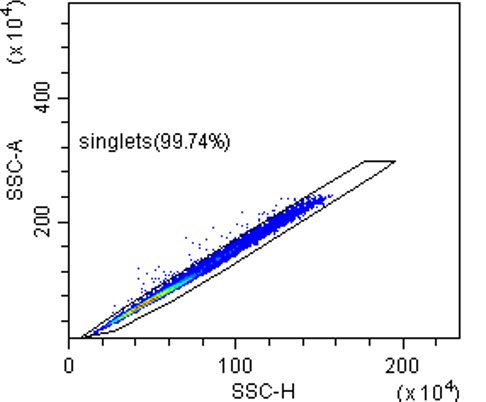

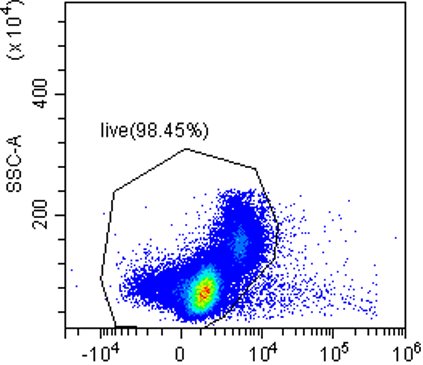

Lymphocytes

Singlets

Live/dead

Singlets

All events

A) **CD14^+^CD16^-^ classical monocytes**

HLA-DR+

CD56-

CD3-

live

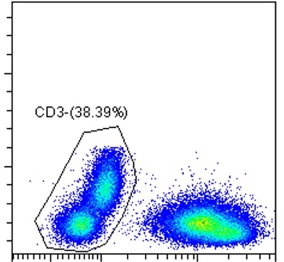

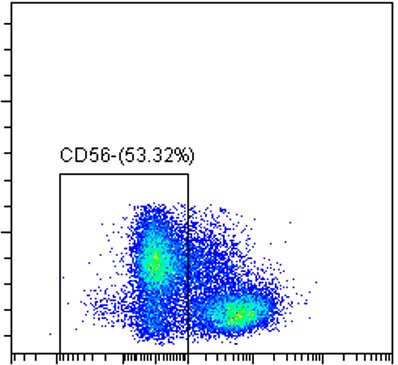

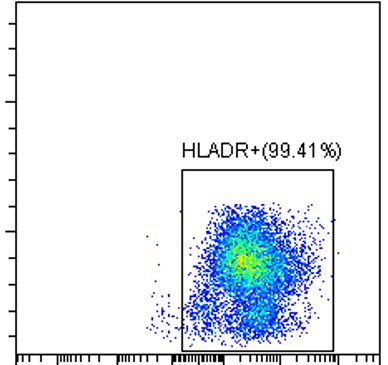

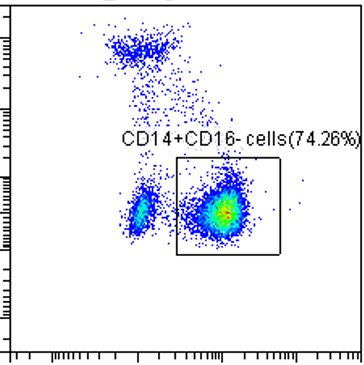

SSC

CD16

CD14

HLA-DR

CD56

CD3

FSC

B) **CD56^dim^CD16^hi^ NK cells**

CD3-

CD14-

Live

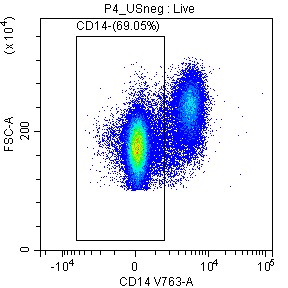

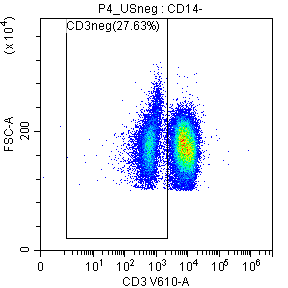

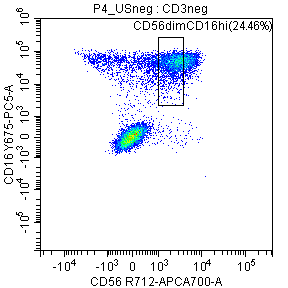

CD56

CD16

CD3

CD14

SSC

FSC

**Figure S5. Flow cytometry gating strategy for identification of classical monocytes and CD56^dim^ NK cell subsets.** Representative flow cytometry plots show sequential gating steps used to identify A) CD14^+^CD16^-^ classical monocytes B) CD56^dim^CD16^hi^ NK cells. Top row) Doublets and debris were excluded based on FSC-A/FSC-H and SSC-A/FSC-A parameters, followed by selection of live singlet cells. (Middle row) Classical monocytes were identified by sequential gating on CD3⁻ cells, followed by exclusion of CD56⁺ NK cells and then selection of HLA-DR⁺ myeloid cells. Within this population, CD14⁺CD16⁻ cells were classified as classical monocytes. (Bottom row) From the CD14⁻ population, CD3⁻ cells were selected and gated on CD56 versus CD16 to identify CD56^dim^CD16^hi^ NK cells.

 
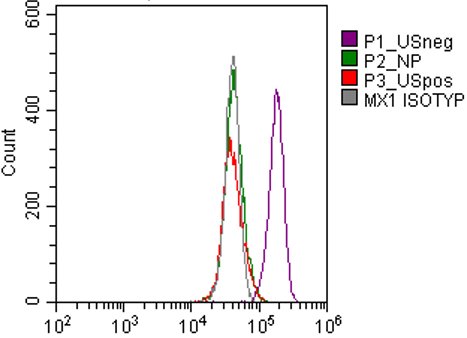

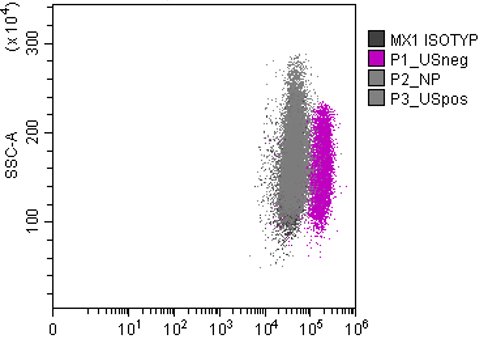

 
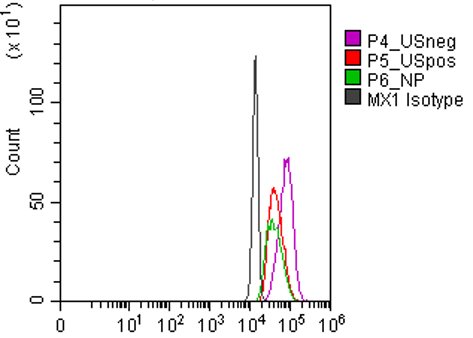

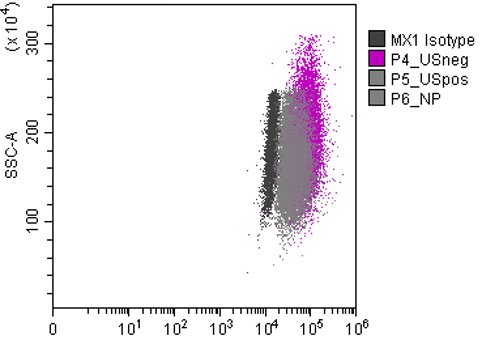

 
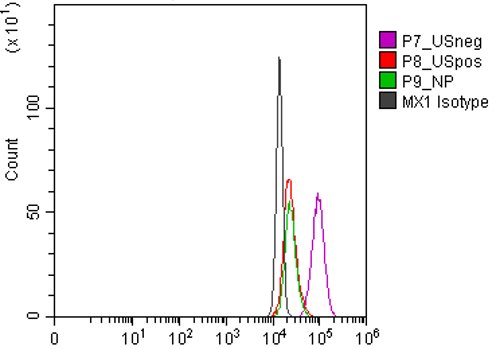

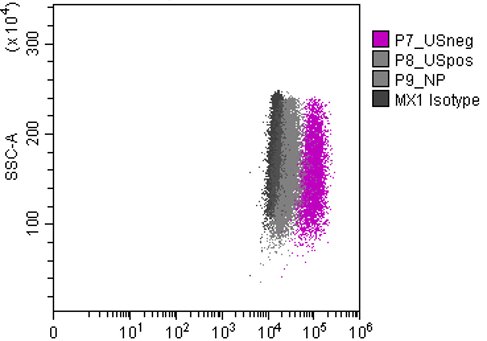

MX1

**Figure S6a. MX1 expression in CD14⁺CD16^-^ classical monocytes.** Representative histograms (left panels) and dot plots (right panels) showing intracellular MX1 protein expression in CD14⁺CD16^-^ monocytes from at-risk RA individuals: NP (green), USneg (purple), and USpos (red) compared with isotype controls (grey). MX1 levels were detected by flow cytometry, demonstrating increased MX1 expression in a subset of USneg participants relative to USpos and NP.

 
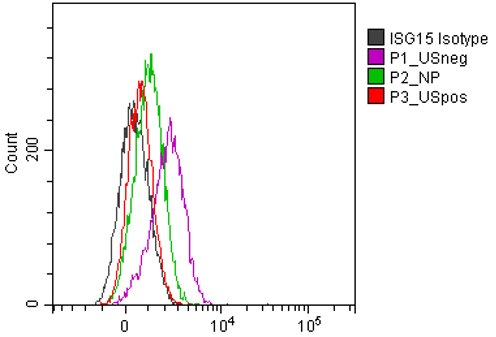
 
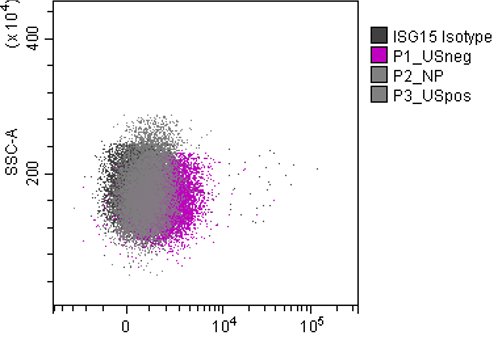

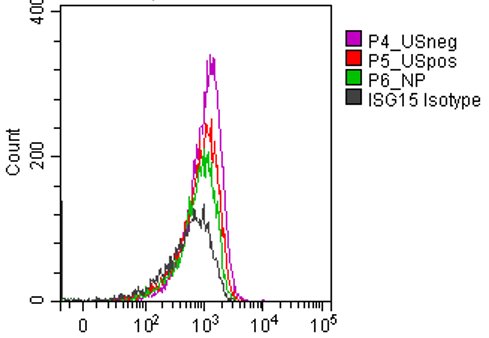

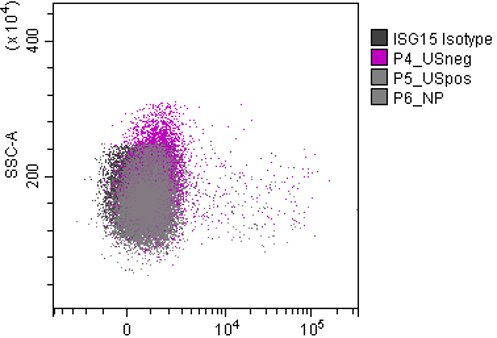

 
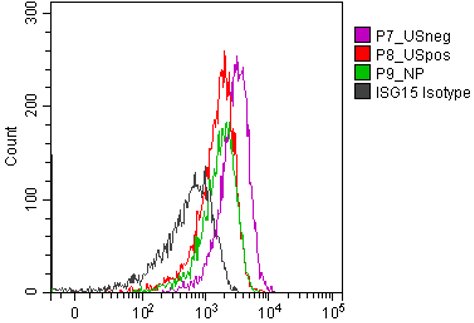

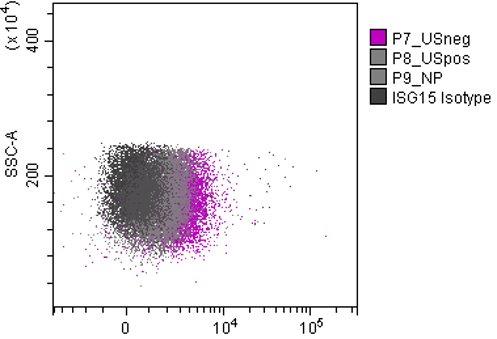

ISG15

**Figure S6b. ISG15 expression in CD14⁺CD16^-^ classical monocytes.** Representative histograms (left panels) and dot plots (right panels) showing intracellular ISG15 protein expression in CD14⁺CD16^-^ monocytes from at-risk RA individuals: NP (green), USneg (purple), and USpos (red) compared with isotype controls (grey). MX1 levels were detected by flow cytometry, demonstrating increased MX1 expression in a subset of US^neg^ participants relative to USpos and NP.

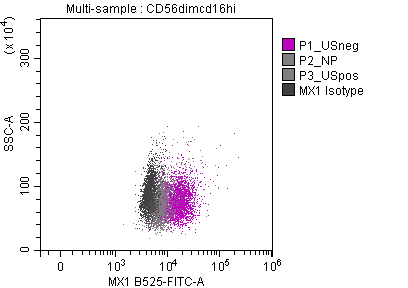

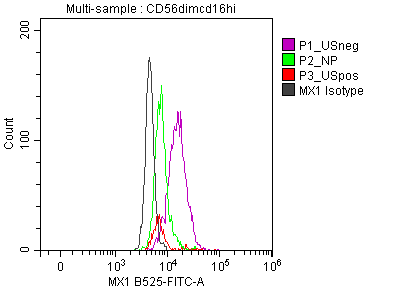

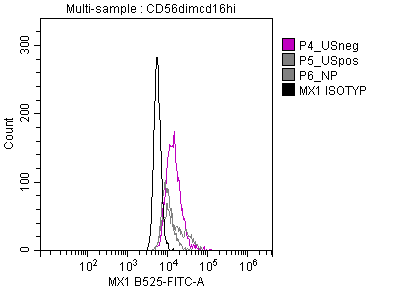

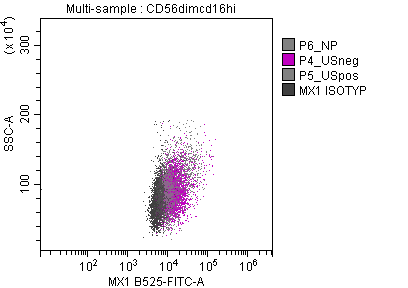

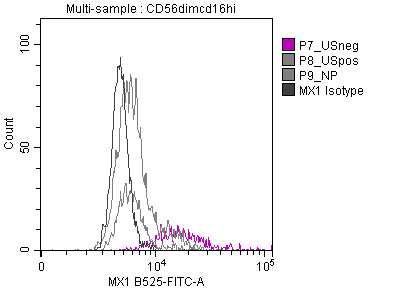

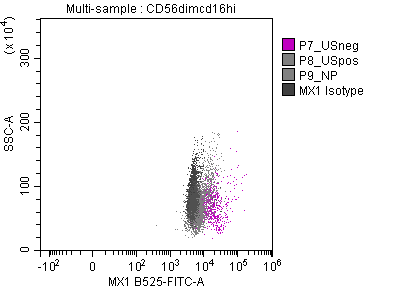

MX1

**Figure S7a. MX1 expression in CD56^dim^CD16^hi^ NK cells.** Histogram overlays (left panel) and dot plot (right panel) showing MX1 expression levels (fluorescence intensity) in CD56^dim^ NK cells from USneg, USpos and NP compared to isotype controls. MX1 expression in increased in USneg (n=3) compared to other groups.

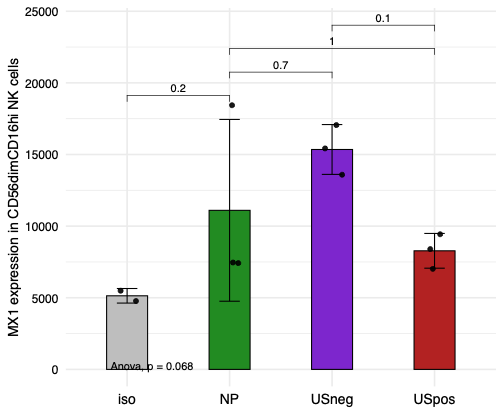

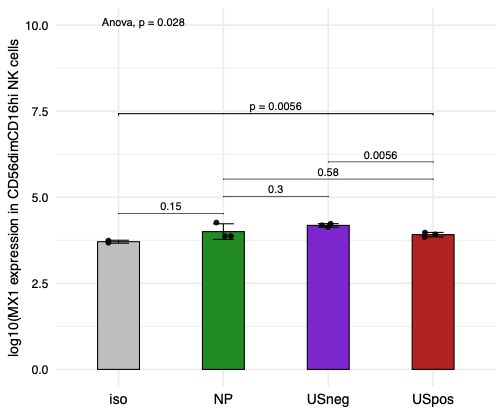

**Figure S7b. Quantification of MX1 expression in CD56^dim^CD16^hi^ NK cells.** (A) Bar plot of raw median MX1 expression values in CD56^dim^CD16^hi^ NK cells across clinical groups. No significant differences were detected using one-way ANOVA followed by pairwise-tests. B) After log-transformation to reduce variance and improve distribution symmetry, the same statistical tests revealed significant differences in MX1 expression between USneg future progressors and the other groups, including NP, USpos, and isotype control.

CD14^+^IL1β^+^ Monocytes

CD14^+^CD16^−^ Monocytes

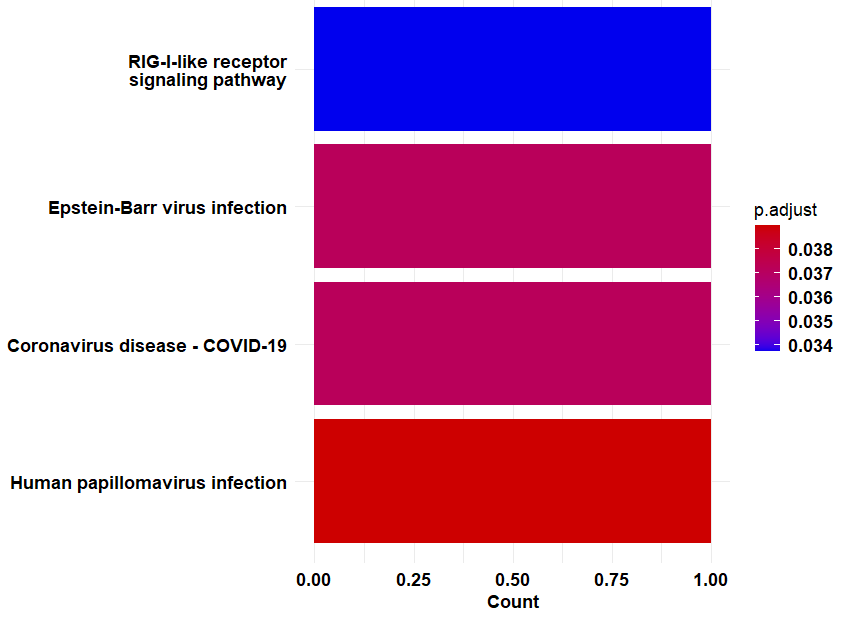

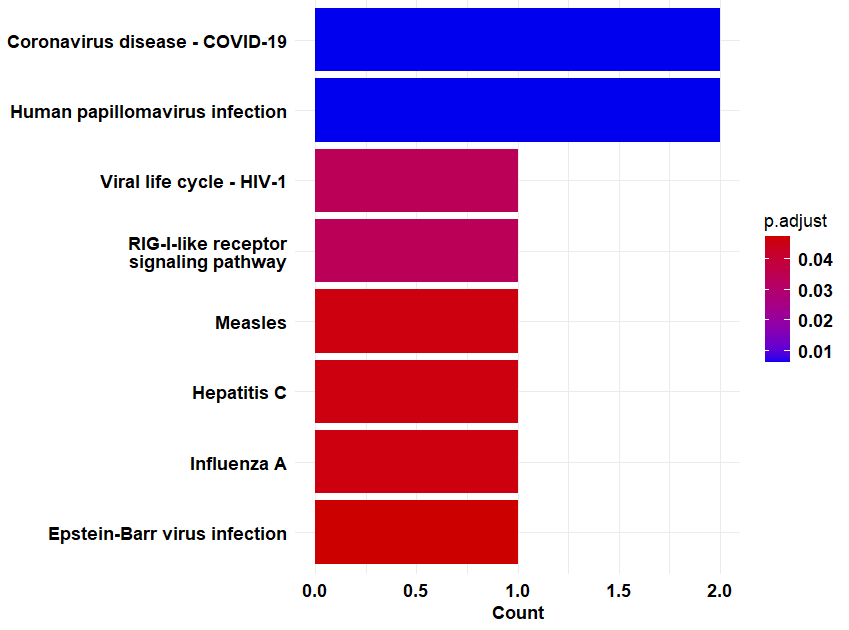

b

a

CD56^dim^CD16^hi^ NK

CD14^−^CD16^+^ Monocytes

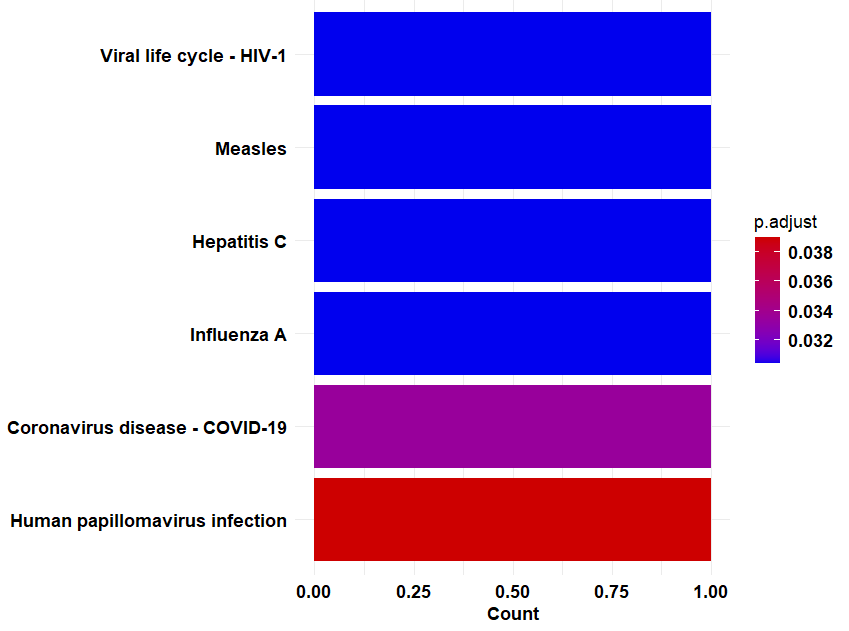

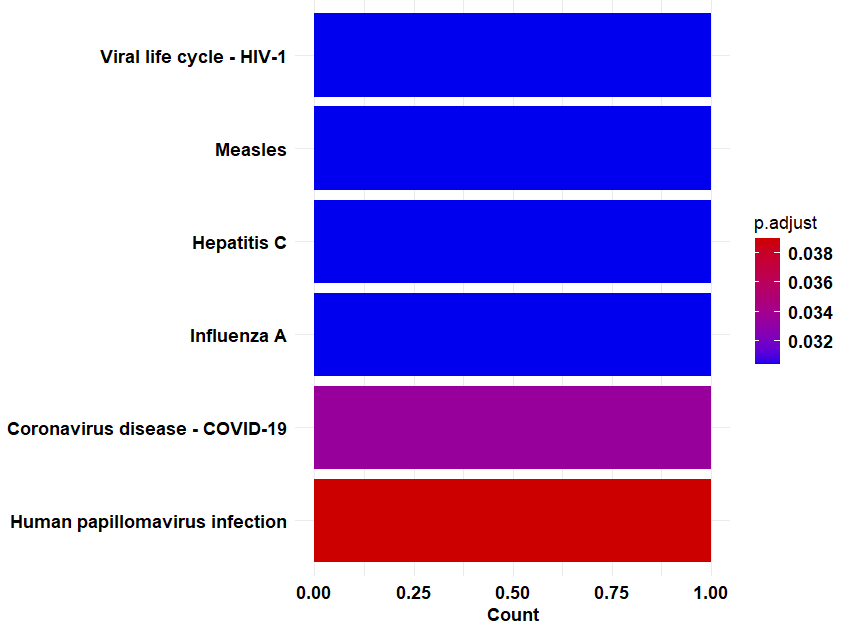

d

c

NK Mitohi

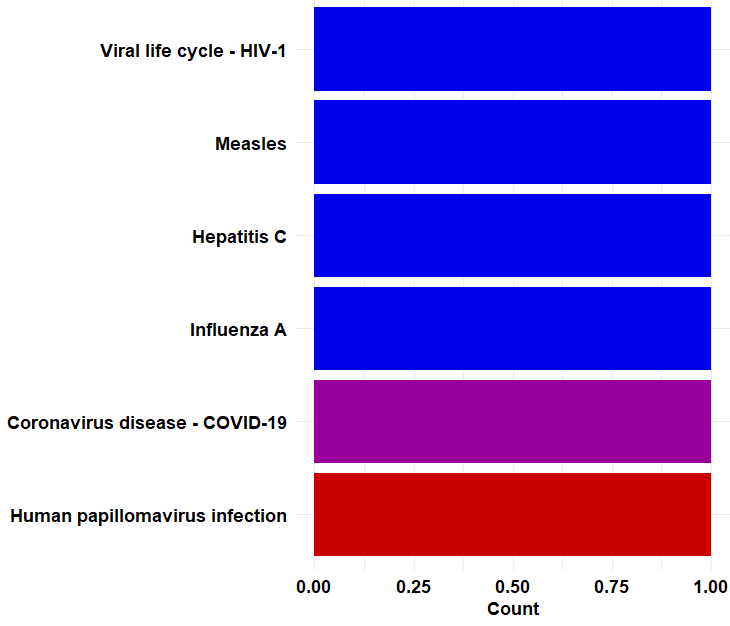

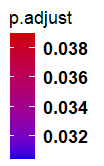

e

**Figure S8. Pathway enrichment across immune cell subsets in at-risk RA groups.** Classical monocytes (**a**) and IL1β⁺ inflammatory classical monocytes (**b**) show enrichment of RIG-I-like receptor signalling and IFN-I-associated pathways. In contrast, non-classical (CD16⁺) monocytes (**c**), CD56^dim^ NK cells (**d**), and NK mitohi with mitochondrial signatures (**e**) are enriched for viral life cycle–related pathways, including those associated with coronavirus, measles, and other viral infections.

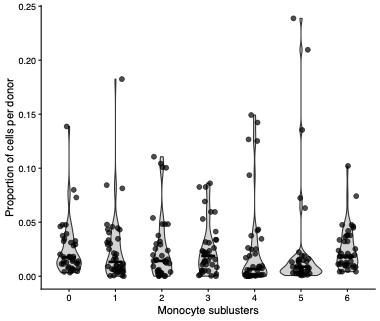

**Figure S9. Proportions of cells per donor in monocyte subclusters.** Violin plots showing the proportion of cells contributed by each donor to individual monocyte subclusters.

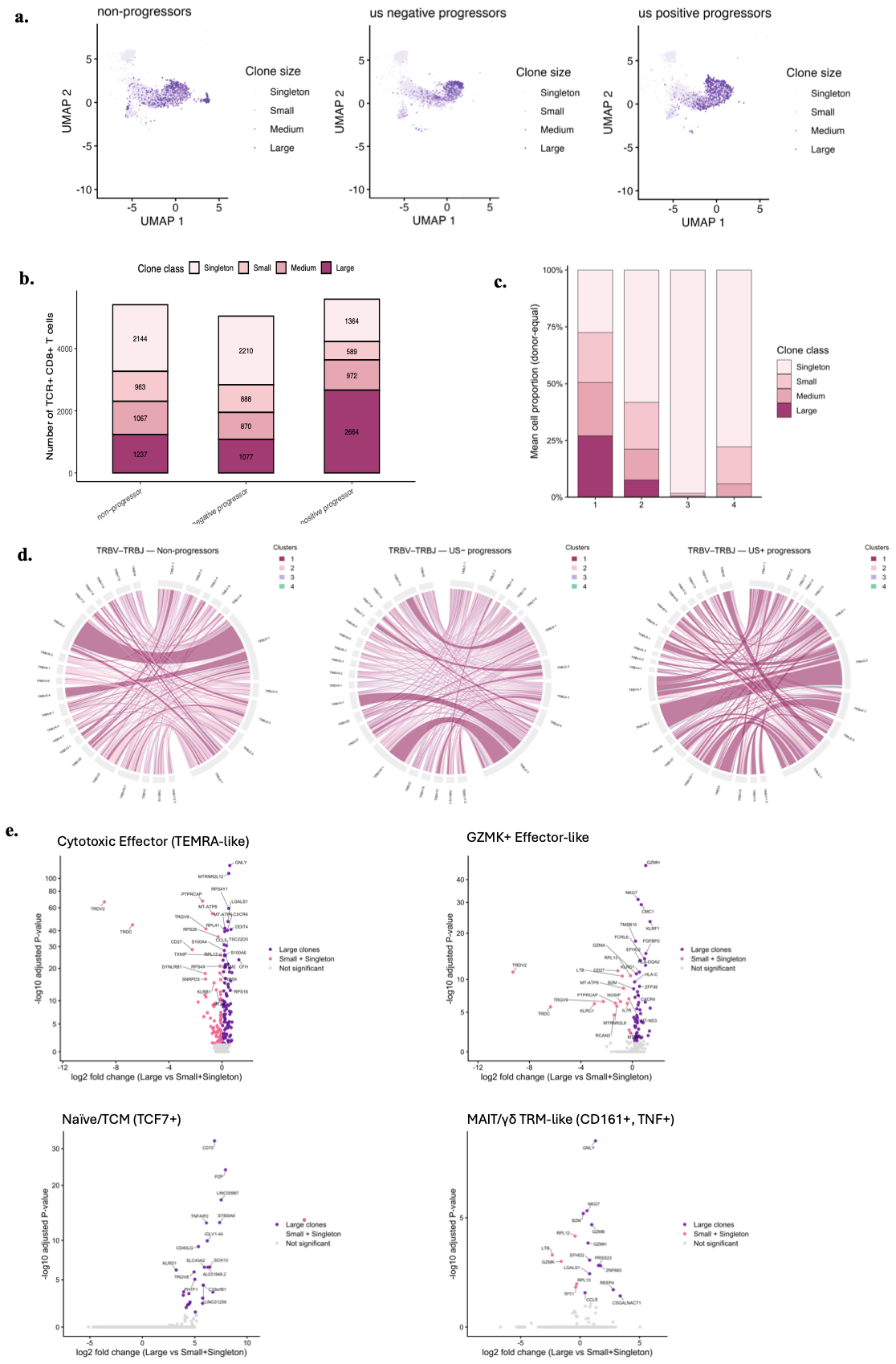
**Figure S10.** **CD8+ T clone class distribution.** UMAP projections showing non-progressors, USneg and USpos future progressors demonstrated that each clinical group displayed distinct distributions of singleton, small, medium and large CD8⁺ T-cell clonotypes across the embedding, with USpos individuals showing the greatest enrichment of expanded clones (a). Group-level quantification confirmed corresponding differences in total CD8⁺ TCR⁺ cell numbers across clone-size classes (b). Mapping clone-size composition onto the transcriptional clusters defined in Figure 4 showed that large clones were concentrated within cytotoxic and TRM-like states, whereas naïve and GZMK-positive effector-like clusters were dominated by smaller clonotypes (c). TRBV–TRBJ pairing networks differed across groups and clone sizes, with expanded clones exhibiting recurrent and skewed V–J combinations (d). Differential expression analyses comparing large versus small or singleton clonotypes within each transcriptional cluster revealed consistent enrichment of cytotoxic and residency-associated genes in expanded clones, while smaller clonotypes retained memory- or regulatory-associated signatures (e).

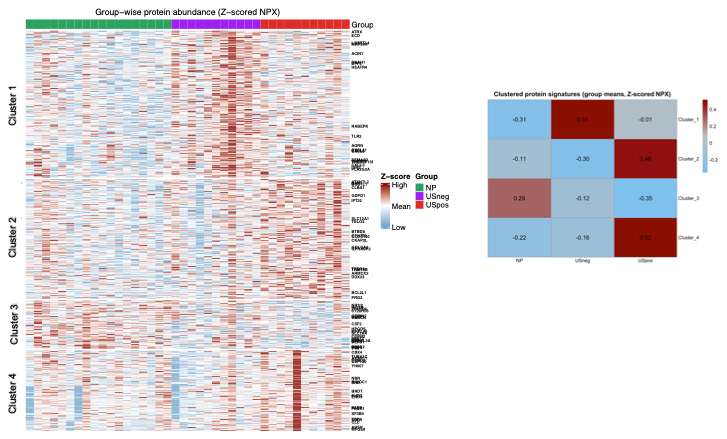

**Figure S11.** **Identification of protein expression clusters associated with ACPA+ at-risk RA groups.** a) Heatmap showing Z-scored NPX values for all measured proteins grouped into four clusters identified by k-means clustering based on shared expression patterns across NP, USneg, and USpos individuals. (b) Cluster-level summary showing the summed Z-scores of proteins within each cluster across the three groups, illustrating differential cluster-level protein abundance between NP, USneg, and USpos.
